# Loss of RUBCN causes autophagy overdrive in a neurodevelopmental disorder with age-dependent neurodegeneration

**DOI:** 10.64898/2026.08.27.26360298

**Authors:** Stephanie Efthymiou, Keisuke Tabata, Hormos Salimi Dafsari, Emil Schober, Christian Latza, Mine Isaoğlu, Anwar Abuelrub, Aboulfazl Rad, Zahra Firoozfar, Valentina Turchetti, Renee Q Lin, Reza Maroofian, Sarah Wiethoff, Erum Afzal, Faisal Zafar, Nuzhat Rana, Annie M. McRae, Rauan Kaiyrzhanov, Ulviyya Guliyeva, Sughra Gulieva, Gia Melikishvili, James Lespinasse, Antonio Vitobello, Anne-Sophie Denommé-Pichon, Ingrid M. Wentzensen, Heather C. Mefford, Lauren C Briere, Melissa A Walker, Frances A High, David A Sweetser, Michael Kendall, Madeleine Franchi, Martha Brown, Donald Latner, Pascal Joset, Ivan Ivanovski, Majid Alfadhel, Iram Alluhaydan, Anne Smedegaard Frederiksen, Vincent Arriens, Britta Hanker, Kshitij Mankad, Julie B Guerin, Serdar Durdağı, Barbara Vona, Heinz Jungbluth, Adam Antebi, Tamotsu Yoshimori, Henry Houlden

## Abstract

Pathogenic variants in *RUBCN*, encoding the Run domain Beclin-1 interacting and cysteine-rich domain-containing protein (Rubicon) have been implicated in autosomal recessive spinocerebellar ataxia 15 (SCAR15). However, the molecular mechanisms underlying disease pathogenesis remain poorly understood. Here, we report 18 individuals from 15 unrelated families harbouring biallelic *RUBCN* variants, who present with an aggressive neurodevelopmental disorder variably characterized by seizures, developmental delay, intellectual disability and movement abnormalities that cause regression, progressive brain atrophy and neurodegenerative features. Through functional characterization, we demonstrate that a subset of disease-associated putative truncating variants disrupt autophagy regulation. In *Caenorhabditis elegans* models, loss-of-function *RUBCN* variants result in an increased autophagic flux and impaired neuronal function, recapitulating key features in humans. Correspondingly, cellular assays reveal that nonsense and frameshift *RUBCN* variants lead to defective autophagy inhibition, underscoring a crucial role for RUBCN as a key negative autophagy regulator. Molecular dynamics simulations rank the eleven missense variants by structural effect, with p.Arg813Trp alone altering the target protein at both the local and the regional level and lying within the RAB7A-binding module that the truncating alleles remove altogether. Our findings establish and expand the *RUBCN*-related disorders as a clinically and molecularly distinct subset of autophagy-related diseases. By delineating both the genetic landscape and cellular consequences of Rubicon dysfunction, this study enhances our understanding of autophagy-related neurodevelopmental disorders and provides a foundation for future therapeutic investigations.

## Introduction

Autophagy (macroautophagy) is an evolutionary conserved degradation pathway that maintains cellular homeostasis by eliminating damaged organelles, misfolded proteins and invading pathogens (1). During autophagy, cytoplasmic cargo is encapsulated by a double-membrane structure called the autophagosome, which subsequently fuses with the lysosome, allowing enzymatic degradation and recycling of dysfunctional cargos (2). This process plays essential roles in various physiological and pathological contexts, including development, immunity, neurodegeneration and cancer (3).

Single-gene disorders affecting the autophagy pathway represent an emerging and heterogeneous group of conditions responsible for a wide range of multisystem diseases in children (4). These disorders primarily impact the central nervous system at various developmental stages, leading to a spectrum of neurological phenotypes, including brain malformations, developmental delay, intellectual disability, epilepsy, movement disorders, and neurodegeneration (5). At the molecular level, pathogenic variants in autophagy-related genes disrupt distinct aspects of this highly conserved process, mostly resulting in defects in autophagosome biogenesis, maturation, or autophagosome-lysosome fusion (6), with variable impacts on autophagic flux, which refers to the overall progression of cellular material through the autophagy pathway to lysosomal degradation.

Rubicon (Run domain Beclin-1 interacting and cysteine-rich domain-containing protein), encoded by *RUBCN* (*KIAA0226*), was initially identified as a Beclin-1-associated regulator of autophagy. Rubicon functions as a negative regulator of autophagosome-lysosome fusion and endocytic trafficking through its interaction with the class III phosphatidylinositol-3 kinase (PI3K) complex (7, 8). Recent studies have identified physiological roles for Rubicon in hepatocyte function (9), adipocyte metabolism (10) and aging (11, 12). Although its role in the brain remains less well characterized, *RUBCN* variants have been associated with autosomal recessive spinocerebellar ataxia (Salih ataxia, SCAR15, OMIM #615705), with a homozygous frameshift variant in *RUBCN* previously identified in two Saudi families (13). However, the precise molecular mechanisms underlying neurological involvement remain elusive, especially regarding the potentially detrimental cellular effects of *RUBCN* loss-of-function (LOF) variants increasing rather than interrupting autophagic flux.

Here, we comprehensively characterize the largest cohort of *RUBCN*-related disorders comprising 18 patients. Our study integrates genetic, clinical, neuroradiological and neuropathological data, complemented by experimental investigations utilizing cellular models and *Caenorhabditis elegans* as a model organism. Our findings elucidate the role of Rubicon as a key negative autophagy regulator and provide novel insights into the clinical and molecular spectrum of *RUBCN*-related disorders.

## Materials & Methods

All reagents, resources, primers and antibodies used in this study are listed in Supplemental Tables 1-2.

### Recruitment of research subjects

Using the GeneMatcher platform (14) and data sharing with collaborators worldwide, 15 families with biallelic *RUBCN* variants were identified. Informed consent for genetic analyses was obtained from all subjects. Clinical details of the cohort were obtained using a standardized proforma. Brain magnetic resonance imaging (MRI) studies were performed as part of the routine diagnostic process and patient care and assessed independently by an expert paediatric (neuro)radiologist. Additionally, we reviewed the literature on all previously published patients with pathogenic *RUBCN* variants and contacted lead authors for up-to-date information.

The study was covered by The Research Ethics Committee Institute of Neurology University College London (IoN UCL) (REC 310045) and the local Ethics Committees of each participating centre. Parents and legal guardians of all affected individuals gave their consent for the publication of clinical and genetic information according to the Declaration of Helsinki. Specific consent was obtained from families for publication of medical photographs and/or video examinations.

### Next generation sequencing

Single-nucleotide variations (SNVs) and indels were identified by exome sequencing (ES) or genome sequencing (GS) in all individuals within this cohort. Genomic DNA was extracted from samples of subjects and their parents, when available, according to standard procedures. Exomes or genomes were captured and sequenced on Illumina sequencers as described elsewhere (15) in Macrogen, Korea for cases identified at UCL or at collaborating centres (16). Briefly, target enrichment was performed with 2 μg genomic DNA using the SureSelectXT Human All Exon Kit version 6 (Agilent) to generate barcoded ES sequencing libraries. Libraries were sequenced with 50x coverage. Quality assessment of the sequence reads was performed by generating QC statistics with FastQC. The bioinformatics filtering strategy included screening for only exonic and donor/acceptor splicing variants. Rare variations present at an allele frequency above 1% in gnomAD v4 (https://gnomad.broadinstitute.org/) or present from exomes or genomes within datasets from UK Biobank and UK 100,000 genome project or from internal research databases (*e.g.*, Queen Square Genomics and UCL SYNaPS Study Group) were excluded. For GeneDx cases, ES protocols have been previously described (17)and variants reported were confirmed by an orthogonal method, as appropriate. Variants were inspected with the Integrative Genomics Viewer and confirmed by Sanger sequencing when appropriate. Sequence variants in *RUBCN* are numbered based on the reference sequence NM_014687.4. Candidate variants were interpreted according to the ACMG Guidelines (18).

### Computational and *in vitro* splice analysis

Computational assessment of splicing effects used SpliceSiteFinder-like, MaxEntScan, NNSplice, and GeneSplicer embedded in Alamut Visual Plus v1.6.1 (Sophia Genetics, Bidart, France), as well as SpliceAI 10K and AbSplice (19) as included in SpliceAI Visual (20). RNA studies of variants were conducted using exon trapping/minigene assays following established protocols with some modifications (21) (22) using two constructs. In brief, the first construct comprised a 1,751 bp region spanning introns 9 to 11, encompassing the c.1474-7C>T variant. The second construct involved a 1,057 bp segment spanning introns 13 to 14 to assay splice effects of the c.1990A>C and c.2051G>A variants. These regions were amplified from genomic DNA obtained from the probands and a healthy control using primers containing specific restriction sites (Supplemental Table 2). The PCR fragments were ligated between exons A and B of a linearized pSPL3-vector following digestion with restriction enzymes XhoI and BamHI. The recombinant vectors were transformed into DH5α competent cells (NEB 5-alpha, New England Biolabs, Frankfurt, Germany), plated and incubated overnight. Following colony PCR with SD6 F (Supplemental Table 2) and the target-specific reverse primer, the wild-type (WT) and variant-containing vector sequences were confirmed by Sanger sequencing and transfected into HEK293T cells (ATCC, Manassas, VA, USA) at a density of 2×10^5^ cells per mL. 2 µg of the respective pSPL3 vectors was transiently transfected using 6 µL of FuGENE 6 Transfection Reagent (Promega, Walldorf, Germany). An empty vector and transfection negative reactions were included as controls. The transfected cells were harvested 24 hours after transfection. Total RNA was prepared using miRNeasy Mini Kit (Qiagen, Hilden, Germany). RNA was reverse transcribed using the High-Capacity cDNA Reverse Transcription Kit (Applied Biosystems, Waltham, MA, USA) following the manufacturer’s protocols. The cDNA was PCR amplified using vector-specific SD6 F and SA2 R primers (Supplemental Table 2). The amplified fragments were visualized on a 1% agarose gel. cDNA amplicons were Sanger sequenced.

### Structural modelling and variant prediction

Due to the limited structural coverage of the protein in the available experimental structures (PDB IDs: 6WCW and 9ZPD), which encompass only a subset of the full-length protein, additional structural models were generated using AlphaFold2 (23) and homology modelling via SWISS-MODEL (24). The impact of missense variants on protein stability was assessed using FoldX (25), which calculates changes in folding free energy (ΔΔG). Variants were classified as stabilizing (ΔΔG < −0.5 kcal/mol), neutral (−0.5 to +0.5 kcal/mol), slightly destabilizing (+0.5 to +1.0 kcal/mol), destabilizing (> +1.0 kcal/mol), or highly destabilizing (> +2.0 kcal/mol). To cross-validate these predictions, mCSM-Stability, a machine learning– based approach, was also applied (26). As the two methods use opposite sign conventions, mCSM predictions were interpreted as stabilizing for positive ΔΔG values and destabilizing for negative ΔΔG values. Protein–protein interaction (PPI) effects were evaluated using mCSM-PPI and, where structural data were available, FoldX-based interface analysis.

### Structural modeling, molecular dynamics (MD) simulations, and trajectory analysis

The available experimental structures of human Rubicon cover only limited regions of the protein. PDB 6WCW resolves the zinc-bound C-terminal RH domain in complex with GTP-loaded RAB7A (27) and PDB 9ZPD contains a restricted Rubicon segment within the PI3KC3-C2 assembly (28). The 972-residue sequence (UniProt Q92622) was therefore modeled with Boltz-2 v2.2.1 (29), with four Zn^2+^ ions supplied as discrete components and chain A of 6WCW provided as a template for the C-terminal region. Twenty-five samples were ranked by aggregate confidence score (30), and the highest-ranked model was selected on the combined basis of that score and its backbone agreement with 6WCW. Prediction confidence was assessed from protein-only predicted Local Distance Difference Test (pLDDT), predicted aligned error (PAE) and predicted distance error (PDE) values (31), both globally and within an 11-residue window centered on each variant position. The model was prepared in Maestro (Schrödinger, LLC) at pH 7.4 (32), minimized under OPLS3e (33) force field (ff), and assessed with ERRAT (34), Verify3D (35) and PROCHECK (36).

Simulations were conducted in Desmond (37) under OPLS3e ff, in orthorhombic TIP3P boxes (38) at 0.15 M NaCl, and run in the *NPT* ensemble at 310 K and 1.01325 bar with particle-mesh Ewald (PME) electrostatics (39). The 14 zinc-coordinating cysteines were modeled as thiolates and His877 and His920 as neutral singly protonated histidines, with metal-donor interactions represented through non-bonded terms. Refinement proceeded in three stages, from restrained simulation with GROMOS-type clustering (40) to a continuous 1 μs unrestrained run, from which the WT reference conformation was drawn as the frame closest to the coordinate average of its final 500 ns (41). Each of the 11 missense variants was then introduced independently into that conformation, giving 12 systems that entered production from a common basin, and each underwent three replicate 200 ns simulations for 7.2 μs of aggregate sampling.

All observables were computed over the final 100 ns of each trajectory on a protein-only selection. Backbone RMSD, Cα RMSF, radius of gyration (R_g_), total and apolar solvent-accessible surface area (SASA) and three-state secondary structure were determined at full length and within the RUN domain (residues 48-189), central region (190–720) and RH domain (721–972) (Figure 1). The micro-environment of each substituted position was characterized from the occupancies of hydrogen bonds, salt bridges and hydrophobic contacts, together with side-chain rotamer populations, and zinc-site integrity was verified in every trajectory. Full analysis parameters are given in Supplemental Methods.

**Figure 1.**
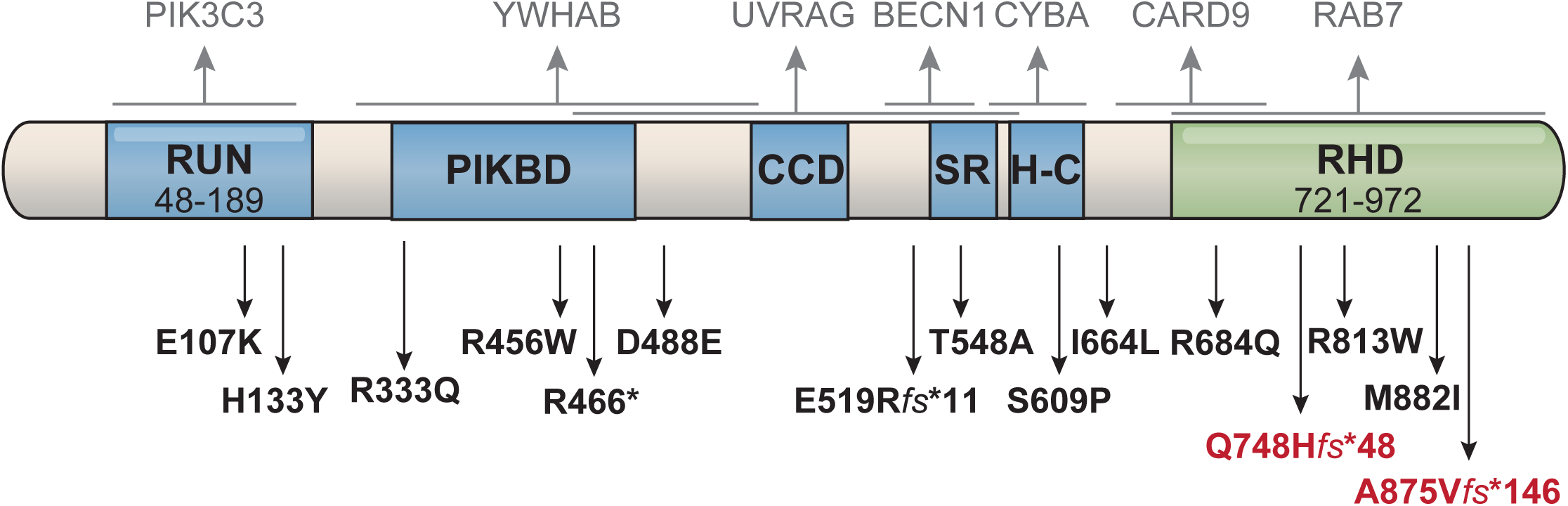
RUN domain organisation and interactions of Rubicon (RUBCN) in pathways implicated in congenital disorders of autophagy. Schematic representation of the human Rubicon protein (972 amino acids), highlighting its major functional domains and known interaction partners involved in canonical and non-canonical autophagy, LC3-associated phagocytosis (LAP) and endocytic trafficking. RUBCN contains a RUN domain (aa 48-189) at the N-terminus which mediates interactions with VPS34 and regulates class III PI3-kinase complex activity. The central region includes the PIKBD (PI3-kinase–binding domain), CCD (coiled-coil domain), serine-rich (SR) region and helix-coil-rich domain (H-C), facilitating binding to Beclin1 and UVRAG and modulating autophagosome maturation. The RHD (RUBCN-homology) domain (aa 721-972) at the C-terminus which is critical for interactions with Rab7 and components of the NADPH oxidase complex (NOX2) and CARD9 during LAP. Binding partners are shown above and RUBCN variants identified in patients are shown below (red: previously reported, black: newly reported).

### Plasmids

All plasmids used in this study are listed in Supplemental Table 1. The full-length human *RUBCN* cDNA was amplified from HeLa cells and cloned into pcDNA3.1+_3 x FLAG vector. Site-direct mutagenesis was used to generate *RUBCN* variants.

### Cell culture

Cells were maintained in Dulbecco’s modified Eagle medium (DMEM), supplemented with 2 mM L-glutamine, nonessential amino acids, 100 U/ml penicillin, 100 μg/ml streptomycin, and 10% fetal bovine serum. For DNA transfection, TransIT-LT1 Transfection Reagent or polyethylenimine (PEI MAX) was used according to the manufacturer’s protocol. For transduction of HEK293 cells stably expressing Halo-LC3, retrovirus was prepared and infected as described (42). In brief, Plat-E cells were co-transfected with the envelope plasmid pLP-VSVG and the pMRX_Halo-LC3 plasmid using PEI MAX. Culture supernatants were collected 48 h post-transfection, filtered, and supplemented with 4 μg/ml polybrene before applied to target cells. After overnight inoculation, the medium was replaced with fresh medium. Two days later, the cells were cultured in medium containing 3 µg/ml puromycin for at least three additional days.

### Immunoprecipitation and immunoblotting

Cells were lysed with lysis buffer (50 mM Tris-HCl [pH 7.5], 150 mM NaCl, 1% TritonX-100 and protease inhibitor cocktail). After centrifugation at 20,000 x g for 10 min at 4°C, the supernatants were incubated with 30 μl of anti-FLAG-M2 agarose beads for 2 h at 4°C. The beads were washed four times with lysis buffer, resuspended with 30 μl of 2x sample buffer (100 mM Tris-HCl [pH 6.8], 4% SDS, 12% β-mercaptoethanol, 20% glycerol, 0.001% bromophenol blue) and boiled at 95°C for 5 min. Supernatants were collected and subjected to immunoblotting. Proteins were separated by SDS-PAGE, transferred to PVDF membranes, and blocked with 5% nonfat milk. Membranes were incubated overnight at 4°C with primary antibodies, washed with 0.5% Tween 20 in PBS, and incubated with HRP-conjugated secondary antibodies for 1 h at room temperature. Signals were detected using Immobilon Forte Western HRP substrate (Merck) and visualized with a ChemiDoc Touch imaging system (Bio-Rad).

### Pulse-chase reporter processing assay and in-gel fluorescence imaging

The pulse-chase reporter processing assay was performed as described previously (43). To assess starvation-induced autophagy flux, HeLa cells stably expressing Halo-LC3 were incubated with 100 nM tetramethylrhodamine (TMR)-conjugated ligand for 20 min at 37°C. After washing, the cells were incubated in starvation medium (EBSS) for 6 h at 37°C. The cells were then lysed in 2x sample buffer and subjected to SDS-PAGE, followed by visualization with ChemiDoc imaging system (BioRad).

### Immunofluorescence microscopy

Cells grown on glass coverslips were fixed with 4% paraformaldehyde in PBS for 30 min, permeabilized with PBS containing 0.1% Triton X-100, and blocked with 5% FBS. The samples were then incubated with primary antibodies for 2 h at room temperature. After three washes with PBS, cells were incubated with Alexa-dye conjugated secondary antibodies in PBS containing 5% FBS for 1 h. Coverslips were mounted in VECTASHIELD mounting medium, and images were acquired using an FV3000 confocal microscope (Olympus).

### Statistical analysis

Unless otherwise specified, values represent the mean of the indicated number of replicates, and error bars denote the SEM as described in the figure legends. Statistical analyses were performed using one-way ANOVA in Prism 9 (GraphPad software). A *P* value < 0.05 was considered statistically significant. All experiments were independently repeated at least twice, as noted in the figure legends. Representative images are shown for immunoblotting, in-gel fluorescence, or microscopy. For the MD simulations, each replicate was reduced to a single value per system, region and observable, giving *n* = 3 (44, 45). Variants were compared with WT by two-sided Welch’s *t* tests (46) with Hedges’ *g* (47) and Holm-Bonferroni adjustment (48). An adjusted *P* below 0.05 was considered significant.

### Generation and characterization of a *C. elegans* model

Using CRISPR/Cas9, the pathogenic patient variant ENST00000296343.10.2624delC, p.Ala875Val*fs*\*146 (major isoform), corresponding to the nematode c.1535delC variant (strain PHX6163, Y56A3A.16), was introduced as the rub-1(syb6163) allele, which carries a viable homozygous deletion. Similarly, the human ENST00000273582.9.1261C>T, ENSP00000273582.5.Arg421* variant (minor isoform), corresponding to the nematode p.Pro91* variant (strain PHX5396), was introduced as rub-1(syb5396); this truncation was viable only in the heterozygous state with the balancer allele hT2[bli-4(e937)let-?(q782)qIs48]. Nematodes were cultured as previously published (49). To assess autophagic flux, we crossed the mutant strains into the reporter strain MAH215 (50) with a lgg-1p::mCherry::GFP::lgg-1 vector for measurement of the autophagosome marker Atg8/LC3/LGG-1 that indicates pre-fusion autophagosomes as GFP punctae and post-fusion autophagosomes as mCherry-only punctae. Three independent biological replicates of 10 hand-picked adult day 1 nematodes each were used for microscopy. Images of nematodes were taken with Andor Dragonfly at ×60 magnification at day 1 adulthood. Punctae counting was performed manually while analyzing researchers were blinded to image titles.

To assess neuronal dendrite morphology, we crossed the mutant strains into the PVD neuronal reporter strain wdIs51(F49H12.4∷GFP) (51). We counted hyperbranching of primary, secondary, tertiary and quaternary dendrites within the first five menorah-like PVD neuronal structures from neuronal soma towards the nematode pharynx. In addition, we counted neuronal specification, contact of menorahs with each other, as well as neuronal bubble-and-bead-like structures (“beading“). Images of nematodes were taken with Andor Dragonfly at ×60 magnification at day 1 adulthood. Counting was performed manually while analyzing researchers were blinded to image titles.

## Results

### Clinical findings

We obtained data of 18 patients with pathogenic *RUBCN* variants from 15 families, of which three are previously published families (13, 52, 53) (Figures 1-2; Supplemental Tables 3-5). We identified 12 previously unreported individuals with SCAR15 of European (n = 5), American (n = 3), Arabic (n = 3) and African (n = 1) origin. Parental consanguinity was reported in 8 families, of whom 6 carried homozygous variants in *RUBCN* (Figure 2A). Reported findings about the extended family history of families included recurrent miscarriages with early neonatal death in one family, occurrence of epilepsy or febrile seizures, psychiatric disorders or ADHD in direct first-degree relatives. The clinical features of the affected individuals are summarized in Supplemental Table 4.

**Figure 2.**
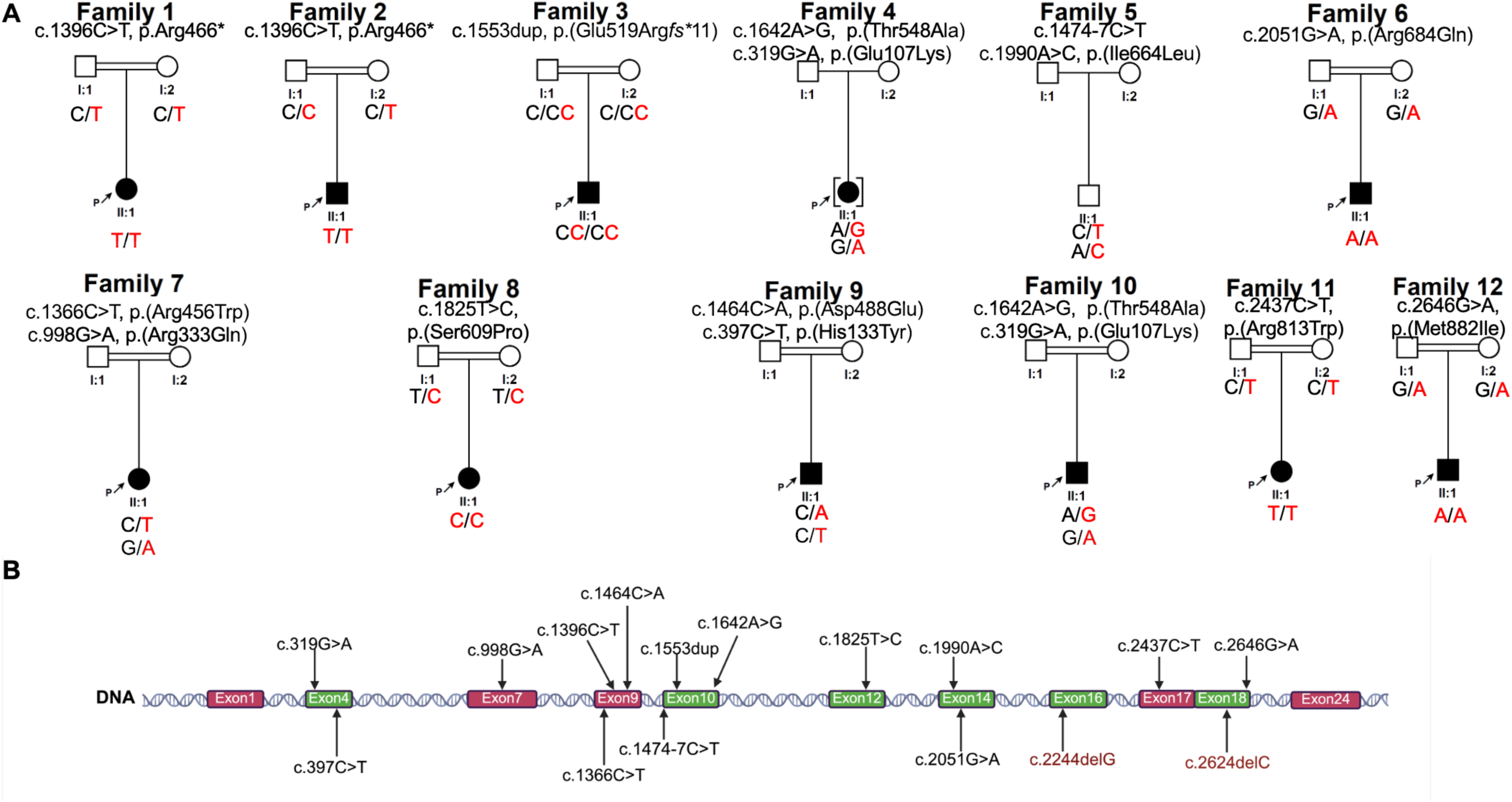
Genetic and clinical presentation of individuals harbouring RUBCN variants. A) Pedigrees of affected families showing segregation of the biallelic RUBCN variants identified. B). Schematic representation of the *RUBCN* gene, with the 24 exons depicted as coloured horizontal boxes. The exon positions of disease-associated *RUBCN* variants are indicated by arrows, with the corresponding nucleotide changes shown adjacent to each arrow. Variants are annotated according to HGVS nomenclature based on transcript NM_014687.4 (red: previously reported, black: newly reported).

### Neurological features

Among the key clinical diagnostic criteria for SCAR15, global developmental delay was universal, with delay in motor, speech and/or cognitive development reported in all assessed individuals. The degree of intellectual disability was variable, ranging from mild to moderate, whereas speech impairment was generally more pronounced, ranging from moderate to severe, with three individuals being nonverbal. Epilepsy was the second most common neurological feature, present in nine individuals and associated with an early onset, except in families 10 and 11 where affected individuals responded to steroid treatment and have remained seizure-free. Three patients were reported with generalized seizures, two with motor status epilepticus and/or non-convulsive status. Most individuals with epilepsy also exhibited a progressive neurodegenerative disorder with prominent movement abnormalities on the background of a non-specific neurodevelopmental disorder. The most common non-neurological finding included failure-to-thrive. Additional variable clinical features include spasticity, choreoathetoid movements, brain atrophy and abnormalities in vision and/or hearing, further expanding the phenotypic spectrum and refining a disorder with both neurodevelopmental and neurodegenerative features. Of note, affected individuals exhibited a syndromic phenotype with variable craniofacial, skeletal, and dermatological features. Microcephaly was observed in five individuals with available follow-up measurements; it was congenital in two, acquired postnatally in one, and could not be classified in two because birth head circumference data were unavailable. Craniofacial features included triangular face, prominent forehead or mild frontal bossing, brachycephaly, deep-set eyes, thick eyebrows, long eyelashes, a depressed nasal bridge with a flat nasal tip and anteverted nostrils, deep philtrum, high-arched palate, and micrognathia. Skeletal manifestations included joint contractures, scoliosis, kyphosis, and mild fifth-finger clinodactyly, while dermatological findings included hypertrichosis and naevi.

### Neuroradiological findings

We obtained information from MRI scans from nine patients with *RUBCN* variants (age ranged between 17 months to 18 years of age) of which six revealed abnormal findings. Brain MRI features in this cohort were suggestive of a structural neurodevelopmental disorder with hypoplasia of the corpus callosum, the basal ganglia and thalamus as well as evidence of delayed brain myelination, alongside cortical malformations and diffuse cerebral and hemispheric white matter hypoplasia. Longitudinal imaging comparison in one individual done during infant age did not demonstrate obvious radiological progression.

Less frequent clinical findings included episodes of tachycardia and congenital heart anomalies (persistent left superior vena cava, patent ductus arteriosus, ventricular septal defect, and hypospadias) in two unrelated individuals.

### Genetic findings

*RUBCN* consists of 24 exons, encodes a protein of 972 amino acids and is expressed across multiple adult human tissues including the central nervous system, skeletal and cardiac muscle, thymus, immune cells, lung and kidneys (52). In the previously reported families, biallelic *RUBCN* variants were identified as homozygous loss-of-function variants as well as compound heterozygous combinations of missense, truncating, and splice-site variants (52–54). Analysis of all *RUBCN* coding exons in 15 families with SCAR15 syndrome revealed 13 affected individuals with homozygous variants and five cases *with* compound heterozygous variants. In total, six individuals had truncating variants on both alleles, one had a putative splicing variant (Family 6), three had missense variants on both alleles, and eight had a combination of missense or splice site variants. Most of the missense variants affected highly conserved amino acids (Supplemental Figure 1). Parental segregation studies supported a recessive mode of inheritance with unaffected heterozygous parents showing no clinical manifestations. An exception was observed in Family 2, in which the affected individual had a homozygous *RUBCN* variant in the context of maternal uniparental disomy (UPD); thus, although the variant is homozygous, it was not inherited from both parents and did not represent the typical biparental transmission of a recessive allele. Type and distribution of *RUBCN* variants identified and the RUBCN protein are illustrated in Figures 1 and 2. Three affected individuals had truncating variants. In two, *individuals* were homozygous for the *RUBCN* Arg466* variant, a likely recurring variant (of no specific ethnic background) with a relatively milder disease progression. Supplemental Table 3 shows *in silico* splice prediction scores that were all unremarkable while pathogenicity assessments are summarized in Supplemental Table 5. RT-PCR of the first construct spanning exons 10-11 to test the c.1474-7C>T variant showed a splice profile identical to WT (Supplemental Figure 2). Together with the second construct spanning exon 14 testing the missense variant c. 1990A>C, identified in trans, that showed a WT construct with leaky splicing, they are unlikely to be pathogenic and do not explain the clinical phenotype (Supplemental Figure 3). The c.2051G>T variant showed correct inclusion of exon 14, thus aberrant splicing was not observed in this assay (Supplemental Figure 4).

### *In silico* structural and molecular dynamics analyses

*In silico* structural analyses were performed to evaluate the potential effects of missense variants on protein stability using FoldX and mCSM-Stability (Supplemental Table 6). Across the dataset, most variants were predicted to have neutral or modest effects on global stability, although a subset showed more pronounced destabilizing effects. The p.(Arg813Trp) variant, located in the C-terminal cysteine-rich region, showed the strongest predicted effect, with ΔΔG values of up to +10.96 kcal/mol by FoldX and concordant destabilizing predictions by mCSM. Similarly, p.(Arg456Trp) showed consistent moderate destabilization across both approaches. In contrast, several variants showed discordant predictions between methods. For example, p.(His133Tyr), located within the RUN domain, was predicted to be strongly destabilizing by FoldX but stabilizing by mCSM, suggesting that its effects may not be adequately captured by global stability predictions. Variants including p.(Asp488Glu) and p.(Thr548Ala) showed near-neutral FoldX predictions but modest destabilizing effects by mCSM, while p.(Arg333Gln), p.(Ser609Pro), p.(Ile664Leu), and p.(Arg684Gln) showed minimal or inconsistent effects across models.

Analysis of protein–protein interaction interfaces using mCSM-PPI revealed a broader tendency toward reduced predicted interaction stability for several variants (Supplemental Table 7). Notably, p.(Arg813Trp) and p.(Arg456Trp) showed consistent predicted destabilization of both protein stability and PPI interfaces. In contrast, p.(Glu107Lys) and p.(Ser609Pro) showed neutral or mildly stabilizing PPI predictions. Overall, these analyses suggest that while only a subset of variants are predicted to substantially affect global protein stability, others may exert more subtle effects on local structure or protein interactions.

### Missense variants remodelled local contacts, and one compacted the RH domain

Model confidence was strongly regional, and seven of the eleven substitutions fall within the least confident interval, so the analyses below are weighted accordingly (Supplemental Figure 5; Supplemental Tables 8 and 9). Refinement improved the ERRAT and Verify3D scores while the Ramachandran distribution became less favourable, and all validation metrics are given in Supplemental Results (Supplemental Figure 6; Supplemental Table 10). The zinc scaffold was preserved throughout. Four-donor coordination was retained in 100% of analysed frames and coordinating-residue mobility did not differ from WT in any variant, the only difference being a 0.028 Å increase in mean donor distance in p.Arg813Trp that left coordination number unchanged (Supplemental Figure 7; Supplemental Tables 11 and 12).

Backbone root-mean-square deviation (RMSD) remained within a narrow band in every system (Figure 3A) and the Cα RMSF profiles were superimposable along the sequence, with the same peaks in the same positions in WT and in every variant (Figure 3B). Of 176 replicate-level comparisons, 174 of which yielded a test statistic, one reached significance after correction. The RH domain was more compact in p.Arg813Trp, at 26.86 ± 0.20 against 27.67 ± 0.11 Å (Hedges’ *g* = -4.07, Holm-adjusted *P* = 0.014), consistently across all three replicates, though both series drifted within the analysis window, so the effect is reported as large and reproducible rather than as a converged estimate. p.Met882Ile was indistinguishable from WT on the same measurement (*P* = 0.92; Figure 3C). Apolar surface area of the same region increased by 110 Å^2^ in the same direction in every replicate without reaching significance (Figure 3D). Effect sizes for every variant, observable and interval are summarized in Figure 3E, and four further effects were replicate-consistent without surviving correction (Supplemental Tables 13 and 14).

**Figure 3.**
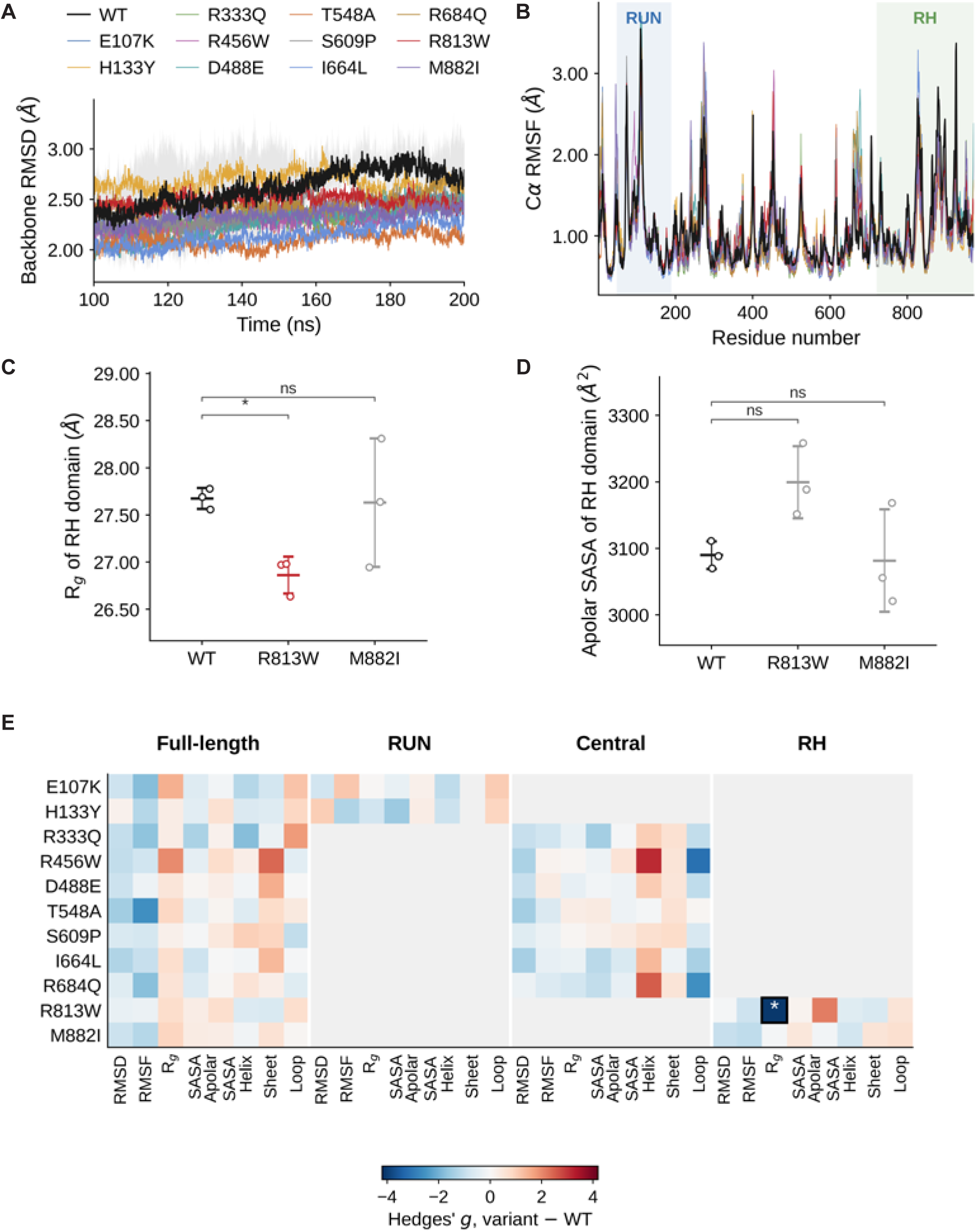
Conformational behaviour of Rubicon across the WT and eleven variant systems. All values are replicate-level, computed over the final 100 ns of three independent 200 ns simulations per system (*n* = 3). (A) Backbone RMSD of the full-length protein, averaged across replicates, the SD shaded for WT. (B) Cα RMSF profile, averaged across replicates, with the RUN (48–189) and RH (721–972) domains shaded. (C) R_g_ and (D) apolar SASA of the RH domain for WT and the two variants located there, each open circle one replicate with the mean and SD marked. (E) Hedges’ *g* for every variant-versus-WT comparison, by sequence interval and observable. Grey cells denote comparisons not made, since only variants located in a given interval were compared there, and the boxed cell marks the single comparison attaining Holm-adjusted *P* < 0.05. Asterisks in C and D denote Holm-adjusted *P*: \**P* < 0.05. ns, not significant.

Local contacts rearranged at eight of the eleven positions, five changes surviving correction. At position 813 the introduced tryptophan formed hydrophobic contacts absent in WT, engaging Leu867 in 97.5% of frames,

Leu847 in 100% and Val809 in 78.5%, while the polar network of the native arginine was lost, including a hydrogen bond to His842 occupied in 70.1% of WT frames (Supplemental Figure 8). Burial of the indole ring against the adjacent leucine face accounts for both the compaction and the increased apolar exposure. Elsewhere, p.Arg684Gln abolished the Arg684-Asp666 salt bridge and Arg333Tyr the Arg333-Asp331 pair, p.Thr548Ala and p.Asp488Glu each formed a new highly occupied contact, and p.His133Tyr, p.Arg456Trp and p.Ser609Pro showed intermediate rearrangements (Supplemental Figure 9; Supplemental Table 15). p.Glu107Lys, p.Ile664Leu and p.Met882Ile showed no change exceeding 15% points.

At nine position-and-dihedral combinations the replicates did not converge on a common dominant rotamer, which bounds the resolution of the local analysis (Supplemental Table 16), and stationarity diagnostics for all 576 analysed series are reported in Supplemental Table 17. Across the eleven substitutions the simulations resolved local contact rearrangements at eight positions and a single regional effect, which arose where local and global changes coincided.

### Truncating variants lacking the C-terminal domain failed to interact with RAB7

Rubicon is a known negative regulator of the autophagosome-lysosome fusion machinery (55) and interacts with RAB7, a small GTPase crucially involved in this machinery (56). To investigate the impact of patient-derived variants on the subcellular localization of RUBCN and its interaction with RAB7, HeLa cells were transiently transfected with FLAG-tagged RUBCN constructs (WT and mutant) (Figure 4). The previously characterized CGHL mutant, which is unable to bind RAB7 and therefore has impaired RUBCN function (56), was included as a control. Expression of the transfected RUBCN variants was confirmed by immunoblotting. All variants were detected at the expected molecular weight, and expression levels were comparable, except for the truncated variants (Figure 4A). As expected, WT RUBCN localized to both cytosolic and punctate structures, where it colocalized with endogenous RAB7. In contrast, the CGHL mutant was diffusely distributed in the cytosol and failed to form puncta, as reported previously (56). Similarly, the patient-derived variants Arg466*, Ala875Valfs*146, and Glu519Arg*fs*\*11 showed cytosolic localization comparable to the CGHL mutant and markedly reduced RUBCN–RAB7 colocalization (Figure 4B). We next assessed the interaction of these variants with RAB7 by immunoprecipitation. Consistent with previous findings, WT RUBCN bound endogenous RAB7, whereas the Arg466*, Ala875Val*fs*\*146, and Glu519Arg*fs*\*11 variants failed to interact with RAB7 (Figure 4C). Indeed, these three variants lack the C-terminal domain that is essential for RAB7 binding. Accordingly, we conclude that under the experimental conditions, the Arg466*, Ala875Val*fs*\*146, and Glu519Arg*fs*\*11 variants are unable to interact with RAB7, as their truncating variants result in the production of putative fragmented RUBCN proteins that no longer retain the RAB7-binding region.

**Figure 4.**
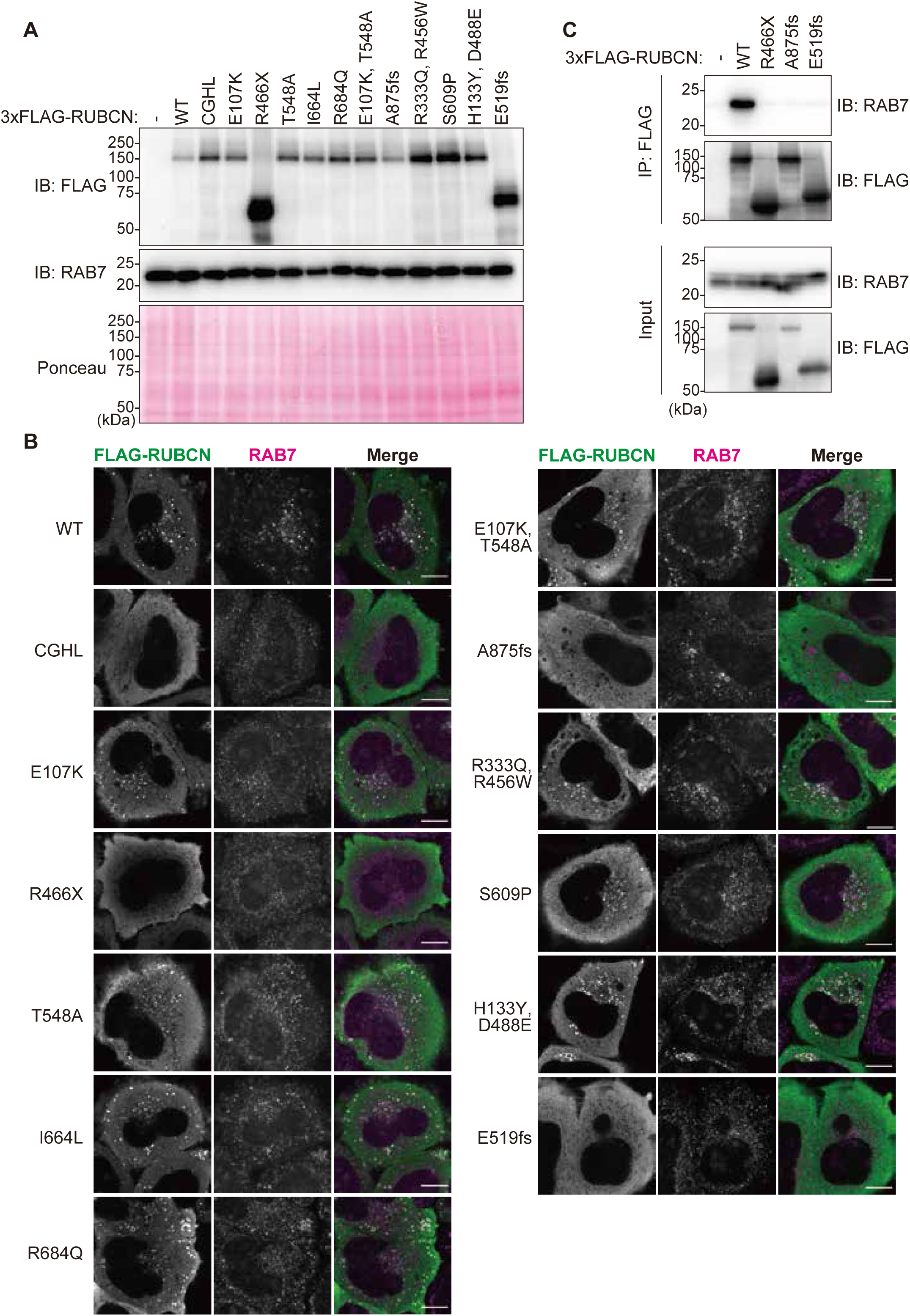
Arg466*, Ala875Val*fs*146* and Glu519Arg*fs*\*11 variants lack RAB7 binding. (A) Expression of RUBCN variants. HeLa cells transiently expressing each 3xFLAG-tagged RUBCN variant were lysed and subjected to immunoblotting. Arg466*, Ala875Valfs*146 and Glu519Arg*fs*\*11 were expressed at reduced levels. CGHL mutant was included as a RAB7-binding-deficient control. (B) Localization of RUBCN variants. HeLa cells transiently expressing each 3xFLAG-tagged RUBCN variant were immunostained with an anti-RAB7 antibody. Scale bars indicate 10 μm. (C) Interaction of RUBCN variants with RAB7. HeLa cells transiently expressing each 3xFLAG-tagged RUBCN variant were subjected to immunoprecipitation using anti-FLAG agarose beads.

### Truncating variants lacking the C-terminal domain show increased autophagic flux

Rubicon has been reported to act as a negative regulator of autophagy, and its binding to RAB7 is essential for this inhibitory function (27) (Figure 5). We next examined whether each RUBCN variant retained the ability to inhibit autophagy. To monitor autophagic flux, we performed a pulse-chase reporter processing assay as described previously (43). This assay exploits the lysosomal protease resistance of ligand-bound Halo tag: pulse labelling with a fluorescent ligand enables tracking of Halo-tagged reporters, and the accumulation of free Halo band upon lysosomal delivery provides a robust and quantitative readout of autophagic flux in mammalian cells. As expected, overexpression of WT RUBCN reduced the accumulation of free Halo band, whereas the CGHL control mutant had no effect. These results confirmed that WT RUBCN inhibits autophagy, whereas the CGHL mutant does not. Importantly, overexpression of the patient-derived variants Arg466*, Ala875Valfs*146, or Glu519Arg*fs*\*11 failed to suppress autophagic flux, similar to the CGHL mutant (Figure 4). Thus, the Arg466*, Ala875Val*fs*\*146, and Glu519Arg*fs*\*11 variants lack the autophagy-inhibitory function of RUBCN.

**Figure 5.**
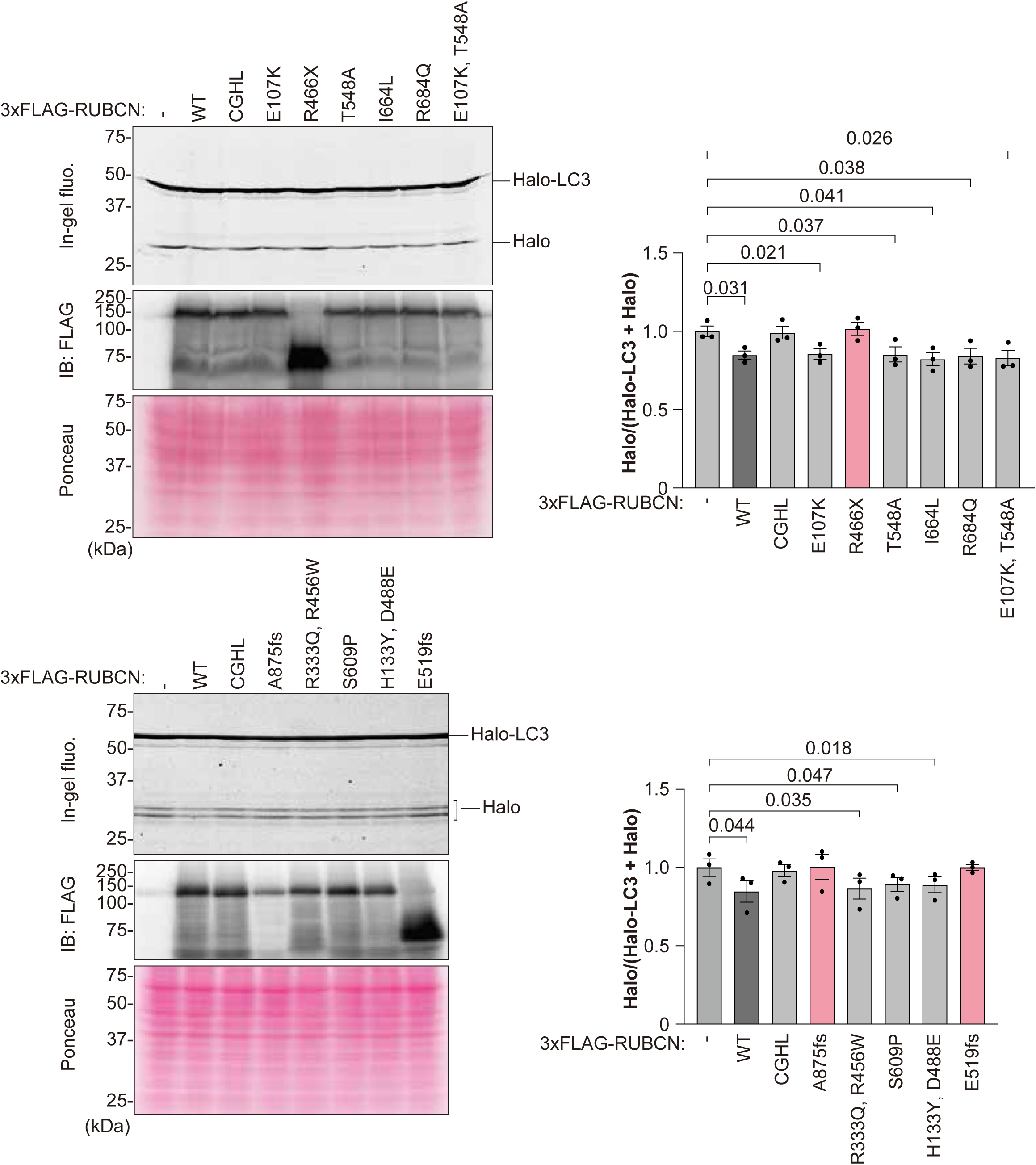
Expression of Arg466*, Ala875Val*fs*146* and Glu519Arg*fs*\*11 variants does not inhibit autophagy. In-gel fluorescence and immunoblotting were performed using total cell lysates from cells overexpressing RUBCN variants. HEK293T cells stably expressing Halo-LC3 were transiently transfected with each RUBCN variant. After 48 h of transfection, cells were subjected to pulse-chase reporter processing assay. Representative images are show on the left. Graphs show the mean ± SEM from three independent experiments. Statistical significance was determined by one way-ANOVA.

**Figure 6.**
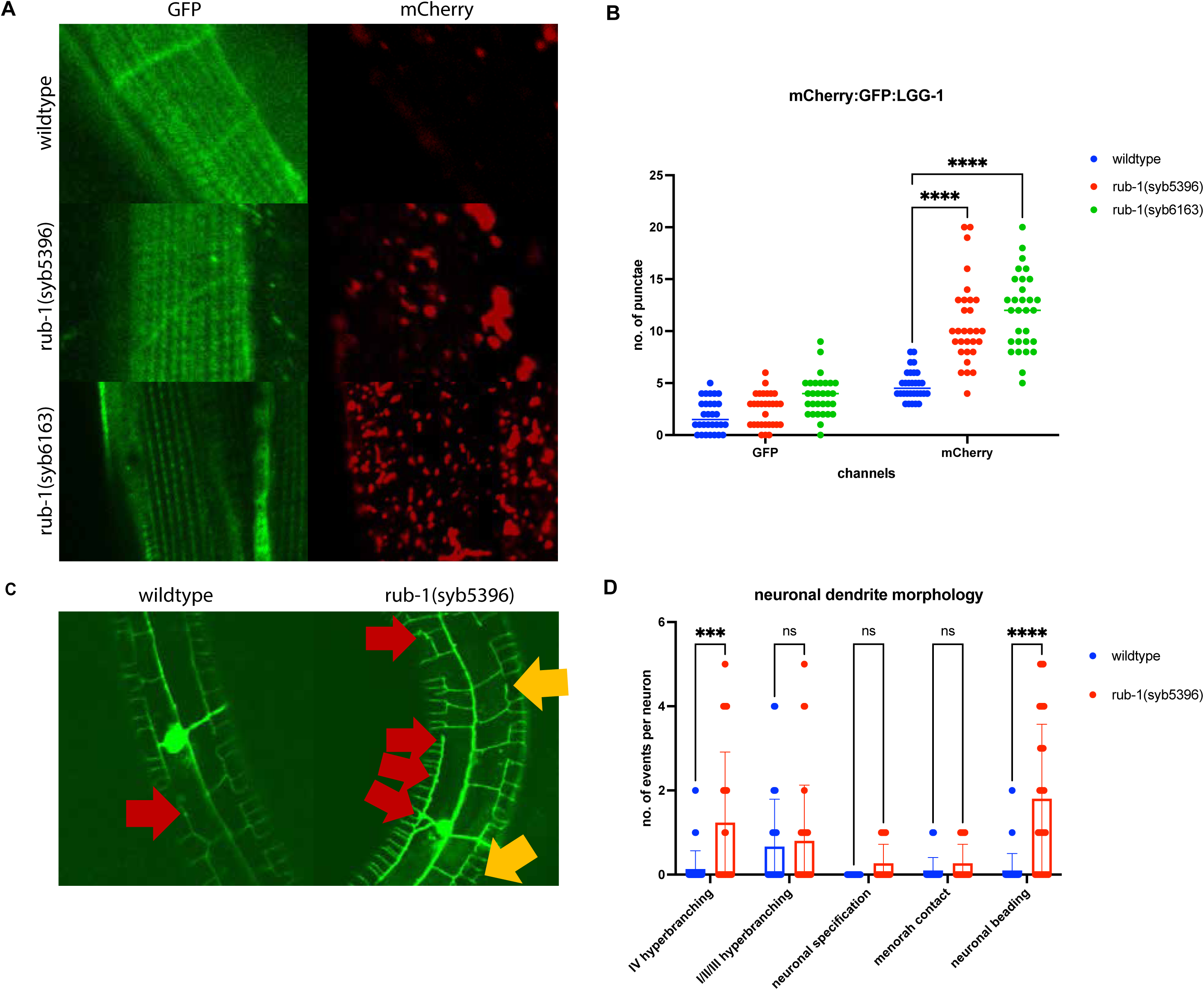
Patient variant modelling in *C. elegans* indicates increased autophagic flux and aberrant neuronal dendrite morphology. (A) Representative fluorescent images of body wall muscle in WT and mutant worms, rub-1(syb5396) and rub-1(syb6163), in a tandem-tagged Atg8/LC3/LGG-1 reporter strain for monitoring autophagic flux, showing similar GFP-positive pre-fusion autophagosomes in all conditions and increased mCherry-positive post-fusion autophagosomes in mutant RUBCN/RUB-1 strains. (B) Quantitative assessment shows similar pre-fusion autophagosome counts in the heterozygous rub-1(syb5396) and an increasing trend in the homozygous rub-1(syb6163) strain and significantly increased post-fusion autophagosome counts in both RUBCN/RUB-1 mutant strains compared to WT. (C) Microscopy pictures of PVD neuronal morphology showing increased neuronal beading (red arrows) and hyperbranching (yellow arrows) in mutant strain. (D) Quantitative assessment shows a significantly increased number of quaternary hyperbranching and neuronal beading in mutant strain.

### Functional studies in *C. elegans* demonstrate variant pathogenicity

Given the conserved role of Rubicon in autophagy and the high degree of conservation between human *RUBCN* and its *C. elegans* ortholog rub-1 (46% similarity), we used *C. elegans* as an *in vivo* model to assess the functional consequences and pathogenicity of selected patient-derived variants (11). First, we inserted patient variants into the genome of the model organism *C. elegans* utilizing a Crispr/Cas9 approach to investigate nematode survival with pathogenic variants. We observed that the human variant ENST00000296343.10:c.2624delC, p.Ala875Val*fs*\*146 (major isoform) that corresponds to nematode c.1535delC (strain PHX6163 Y56A3A.16) showed a viable deletion in homozygous state as rub-1(syb6163). However, the human variant ENST00000273582.9:c.1261C>T, ENSP00000273582.5:p.Arg421* (minor isoform) that corresponds to the nematode p.Pro91* (strain PHX5396) was a truncation that was only viable in a heterozygous state with a balancer allele as rub-1(syb5396)/hT2[bli-4(e937)let-?(q782)qIs48]. The human p.Arg421* variant is localized much earlier in the protein, likely leading to absence of the expression of this essential protein for autophagic flux, which would be in line with our observation that homozygosity of these variants in *C. elegans* would not be viable. However, the other more C-terminal human variant in p.Ala875Val*fs*\*146 was indeed viable in homozygosity in *C. elegans*.

To assess the effect of patient variants on autophagic flux in an *in vivo* model organism, we investigated the body wall muscle in CRISPR/Cas9-engineered mutant nematodes crossed into the reporter strain MAH215 which indicates GFP-positive pre-fusion autophagosomes and mCherry-positive post-fusion autophagosomes due to the pH drop and GFP-quenching with autophagosome-lysosome fusion (Figure 5A). We observed that mCherry-positive post-fusion autophagosomes were significantly upregulated in both mutant strains, while GFP-positive pre-fusion autophagosomes were similar to WT (Figure 5B). This observation of an increased autophagic flux is in line with the findings in cellular assays of *RUBCN*-related disorders.

To assess the effect of patient variants on neuronal morphology *in vivo*, we investigated the heterozygous mutant strain rub-1(syb5396) after crossing with the PVD (multimodal sensory/proprioceptive neurons named anatomically by their cell body position and lineage as a Posterior Ventral-class neuron) neuronal reporter strain (Figure 5C). Attempts of crossing the homozygous mutant strain rub-1(syb6163) did not produce viable offsprings, likely because the PVD neuron is involved in egg-laying (57) and autophago-lysosomal genes are crucial for molting in *C. elegans* (58). In the heterozygous rub-1(syb5396) strain, we observed significant increases in quaternary hyperbranching (Figure 5C) as well as neuronal beading (Figure 5D). This observation of aberrant neuronal dendrite morphology in nematodes *in vivo* may be in line with the finding of brain malformations in patients with *RUBCN*-related disorders.

## Discussion

In this study, we expand the clinical, genetic, and mechanistic landscape of autosomal-recessive RUBCN-related disorder. We provide evidence for an expanding phenotypic range associated with RUBCN defects, suggesting a relatively more severe end of the clinical spectrum that is more neurodevelopmental than motor delay. Our findings suggest that RUBCN-related disorder may include dysmorphic craniofacial features, neurodevelopmental delay, epilepsy and spasticity. By combining deep phenotyping of a multi-ethnic cohort with cellular and in vivo functional analyses, we demonstrate that loss of RUBCN function leads to overly increased autophagic flux and aberrant neuronal morphology, providing a unifying disease mechanism that links genotype to neurodevelopmental and neurodegenerative phenotypes.

Our patient cohort confirms RUBCN-related disease as a neurodegenerative condition evolving on the background of a neurodevelopmental disorder with prominent movement abnormalities, epilepsy, and cognitive impairment. While SCAR15 has historically been conceptualized as a relatively “pure” form of cerebellar ataxia, our findings broaden the clinical spectrum to include individuals in whom developmental delay, epilepsy, spasticity, and syndromic features may be prominent. Epilepsy was frequently of early onset and severe, including motor and non-convulsive status epilepticus. In two individuals, seizures responded to corticosteroid treatment and remained in remission, suggesting that epilepsy may be amenable to therapeutic intervention, although whether this alters the overall disease course remains unknown. Beyond the neurological involvement, several patients displayed distinctive craniofacial, skeletal and dermatological features, that included characteristic facial dysmorphology, joint contractures, scoliosis, hypertrichosis and naevi. These findings indicate that RUBCN-associated disease extends beyond the nervous system and should be considered a multisystem neurodevelopmental syndrome rather than a narrowly defined ataxia.

To provide a structural basis for that characterisation, we modeled full-length Rubicon and simulated each missense substitution within a common conformational framework, on a zinc scaffold whose four-donor coordination remained intact throughout. Only p.Arg813Trp produced a significant effect at both the regional and the local level. Substitution of the solvent-facing arginine by tryptophan exchanges a polar contact network for persistent packing against an adjacent leucine face, compacting the domain while exposing a greater apolar surface. Similar rearrangements have been observed in other target proteins, where burial of a bulky aromatic side chain at an exposed position reorganises packing without unfolding the surrounding fold (61, 62). Since the RH domain is recruited to late endosomal membranes through GTP-loaded RAB7A (52), reorientation of this surface may provide a structural counterpart to the loss of RAB7A binding seen with the truncating alleles, arising through reorganisation rather than deletion. Position 813 was also the best-resolved of the eleven, placing this inference on firmer ground than any drawn elsewhere. The remaining substitutions acted locally and in both directions, abolishing charge pairs at p.Arg333Gln, p.Arg456Trp and p.Arg684Gln, creating new contacts at p.Thr548Ala, p.Asp488Glu and p.His133Tyr, combining both at p.Ser609Pro as expected of a proline, and leaving no trace at p.Glu107Lys, p.Ile664Leu and p.Met882Ile. Loss of a near-permanent salt bridge has likewise been shown to destabilise a protein locally without gross unfolding (63). Importantly, these modelling approaches provide a framework for prioritising variants for experimental validation, while predictions should be interpreted cautiously given variable confidence across the full-length model and the limited timescale accessible to molecular dynamics simulations.

At the molecular level, functional assays investigating RUBCN-RAB7 interaction, demonstrate that patient-derived truncating variants p.Arg466*, p.Ala875Val*fs*\*146 and p.Glu519Arg*fs***\***11 abolish the interaction between RUBCN and RAB7. This interaction is required for RUBCN-mediated regulation of autophagy, and loss of the C-terminal RAB7-binding domain consistently resulted in cytosolic mislocalisation, failure to form punctate structures and inability to bind endogenous RAB7. Consistent with these observations, overexpression of these patient-derived variants failed to suppress autophagic flux in mammalian cells, phenocopying a previously characterized RAB7-binding-deficient mutant. These findings support impaired RUBCN-mediated regulation of autophagy as a shared molecular consequence of pathogenic *RUBCN* variants and are consistent with a loss-of-function disease mechanism. However, recent studies indicate that RUBCN function is more complex than that of a constitutive negative regulator of autophagy. RUBCN isoforms can exert opposing effects on autophagy in a cell-type-dependent manner, with the shorter RUBCN isoform promoting autophagy in B cells, while RAB7A phosphorylation can selectively relieve RUBCN-mediated inhibition during Parkin-dependent mitophagy without substantially affecting bulk autophagy. Thus, loss of RUBCN function may have context-dependent effects on distinct autophagic pathways rather than resulting in a uniform increase in autophagy (59, 60).

An important limitation is that the experiments were performed using truncated ORF constructs and therefore do not establish whether the corresponding endogenous transcripts undergo nonsense-mediated decay or the extent to which this contributes to the disease mechanism. Moreover, the functional consequences of the missense variants remain less well understood. Although our structural modelling identified variant-specific changes in local residue interactions and regional conformational behaviour, these predictions do not establish how such changes affect RUBCN function or contribute to disease. Of note, as our functional studies were performed under basal conditions, it remains to be determined whether the loss of RUBCN-mediated autophagy inhibition is preserved under autophagy-inducing stress conditions.

Modelling of RUBCN variants in the model organism *C. elegans* gives insights into the effects of *in vivo* increases in autophagic flux. This is in line with previous reports on RUBCN knockout or knockdown in mice, flies and worms (11). While several congenital disorders of autophagy are associated with a downregulation in the activity of the autophagosome-lysosome fusion complex machinery, only loss-of-function RUBCN variants were reported with actual upregulation of autophagic flux (55). While RUBCN loss-of-function was reported with lifespan extension in basic science publications (11), it remains unclear whether these patient variants exert similar effects in human despite the neurological phenotype reported. Moreover, whether broad upregulation of macroautophagy is beneficial in the context of human ageing remains an open question, as the effects of autophagy modulation may depend on the tissue, cellular context, and extent of pathway activation (61).

In addition, our *C. elegans* model showed aberrant neuronal dendrite morphology. Previous studies showed downregulation of neuronal beading (axons or dendrites develop swelling or blebbing along their length) with knockdown of the autophagy tethering factor *epg-5* (51), a congenital disorder of autophagy known to cause autophagy blockade (62, 63). The observation of an increased neuronal beading in the setting of this RUBCN-deficient upregulation of autophagic flux may further suggest that, for different reasons, an autophagy overdrive may also be detrimental to neuronal longevity in patients with *RUBCN*-related disorders showing age-dependent movement disorders. Of interest, the clinical overlap between RUBCN-related disease and disorders caused by impaired autophagic degradation suggests that convergent downstream mechanisms, or compensatory activation of autophagy upstream of the block, may contribute to neurodevelopmental and neurodegenerative phenotypes, although this hypothesis requires experimental validation.

Notably, we also observed hyperbranching of the quaternary dendrite in a *rub-1* mutant strain (Figure 5D). Ectopic branching was previously reported in *dhc-1* mutant strain (64), an ortholog of the human *DYNC1H1* that if mutated is associated with neuropathies and neurodevelopmental disorders in human (65). While the hyperbranching in nematode RUB-1 deficiency does not have a clear correlate in human neuroradiological data, it may be consistent with the abnormal neuronal morphogenesis underlying structural brain abnormalities observed in some individuals, including callosal abnormalities. We propose that tightly-regulated autophagy is required in the quality control of post-mitotic neurons even during their formation and any dysregulation in autophagic flux leads to aberrant dendrite morphology, as evident by ectopic dendrites in several nematode genes responsible in autophagosome trafficking, *unc-116*, *dhc-1* and *bicd-1* (64). The lack of changes in neuronal soma size is evident in *rub-1* mutant strains and was previously also reported in worms with downregulated mTOR, upregulated autophagy, and dysregulated ER exit sites (66). In summary, patient variant modelling in *C. elegans* recapitulates the overdrive in autophagic flux and neurodevelopmental disorder from human *RUBCN*-related disorders.

The impact of pathogenic variants in genes such as *RUBCN* extends beyond canonical autophagy, influencing other mechanisms such as exosomal secretion through distinct mechanisms (12, 67). This suggests that pathogenic variants may exert pleiotropic effects on intracellular and intercellular communication pathways. The phenotypic consequences of these variants appear to be modulated by both variant type (missense vs. nonsense/frameshift) and allelic configuration (homozygous vs. compound heterozygous). Here, we provide evidence that homozygous LOF variants, particularly nonsense and frameshift variants, are predicted to result in near-complete loss of protein function, severely impairing the ability of RUBCN to negatively regulate autophagy. Whether these variants similarly affect exosomal secretion due to impaired endosomal-lysosomal degradation remains to be determined. In contrast, compound heterozygous individuals carrying a combination of missense and truncating or splice-site variants may retain partial protein function, resulting in intermediate phenotypes. Similarly, individuals harbouring two missense variants, likely representing hypomorphic alleles, may preserve sufficient residual RUBCN activity to partially compensate for autophagy dysregulation and associated trafficking defects, potentially explaining the milder disease progression observed for example in family 7. These observations underscore the importance of functional characterization of individual variants to accurately interpret genotype– phenotype relationships in *RUBCN*-related disease (Table 1).

Collectively, our data support a disease model in which Rubicon LOF disrupts autophagy homeostasis during neurodevelopment, likely leading to abnormal neuronal maturation and long-term vulnerability of post-mitotic neurons. Pathogenic variants that impair RUBCN-RAB7 interaction release a critical inhibitory checkpoint on late-stage autophagy, resulting in sustained upregulation of autophagic flux. This mechanism distinguishes RUBCN deficiency from other congenital disorders of autophagy and underscores the importance of precise autophagy regulation, rather than simple activation or suppression, in maintaining neuronal integrity. While increased autophagy may initially be tolerated or even beneficial in some cellular contexts, chronic overactivation in neurons appears to compromise dendritic architecture, synaptic integrity and neuronal resilience. This is supported by converging evidence from patient neuroimaging and the complementary cellular and *in vivo C. elegans* models demonstrating excessive autophagic flux, aberrant dendritic hyperbranching and neuronal beading. We propose that this early disruption of neuronal structural homeostasis establishes a neurodevelopmental substrate upon which progressive motor and epileptic manifestations emerge, thereby unifying the developmental and degenerative components of RUBCN-related disease.

Together, these findings define RUBCN-related disease as a disorder of autophagy homeostasis that disrupts neuronal development and long-term maintenance. By integrating human genetics with mechanistic studies across model systems, this work provides a framework for improved diagnosis of unsolved neurodevelopmental disorders and establishes a foundation for future therapeutic strategies aimed at restoring autophagy balance in RUBCN-associated disease.

## Data Availability

All data produced in the present study are available upon reasonable request to the authors. The full-length structural model of human Rubicon generated with Boltz-2 has been deposited in ModelArchive and is publicly accessible at DOI 10.5452/ma-o1x4b.

## Acknowledgements

The authors would like to thank the affected individuals and their families for their support of this study. S.E. expresses her sincere gratitude to Prof E. Davis for her valuable feedback and support in improving this article.

## Funding

S.E. is supported through a fellowship from the Jack Bear Foundation (USA). The Houlden lab is grateful for the important support from patients and families, our UK and international collaborators, brainbank and biobanks, and grateful for essential funding from The Wellcome Trust (221951/Z/20/Z), The MRC, The MSA Trust, UK Dementia Research Institute, The National Institute for Health Research University College London Hospitals Biomedical Research Centre NIHR-BRC), The Michael J Fox Foundation (MJFF), The Fidelity Foundation, Rosetrees Trust, EAN, ERDERA: European Union’s Horizon Europe research and innovation programme, The Dolby Family Fund, Alzheimer’s Research UK (ARUK), Mission MSA, Defeat MSA, Parkinson’s disease UK, Parkinson’s Foundation, Muscular Dystrophy UK, Ataxia UK, CureDRPLA, ALS Association, National Ataxia Foundation (NAF), Target ALS Foundation, Medical Research Foundation and The National Brain Appeal. This research was conducted as part of the Queen Square Genomics group at University College London, supported by the National Institute for Health Research University College London Hospitals Biomedical Research Centre. K.T. is funded by JSPS KAKENHI Grant Number JP21K06169 and J25K09567; UCL-OU seed fund; RDMM-Europe. B.V. is funded by the Deutsche Forschungsgemeinschaft (DFG, German Research Foundation), via the DFG Heisenberg program VO 2138/8-1 grant 543719215, and the DFG Collaborative Research Center 1690 (Project A03). H.S.D was funded by the Jack Bear Foundation, the Koeln Fortune Program/Faculty of Medicine, University of Cologne (371/2021 and 243/2022), as well as the Cologne Clinician Scientist Program/Medical Faculty/University of Cologne and German Research Foundation (CCSP, DFG project No. 413543196). A.A. was supported by the Max Planck Gesellschaft. E,S. was funded by the Koeln Fortune Program/Faculty of Medicine, University of Cologne as well as the Friedrich-Ebert-Stiftung Studiumsstipendium. D.R.L. was funded by the U.S. National Institutes of Health - Eunice Kennedy Shriver National Institute of Child Health and Human Development (NICHD; 1R01HD112437), and by the US National Human Genome Research Institute (NHGRI; UM1HG007301).

## Conflict of interest statement

IMW is an employee of and may own stock in GeneDx, Inc. All other authors declare no conflict of interest. We confirm that we have read the Journal’s position on issues involved in ethical publication and affirm that this report is consistent with those guidelines.

## Data availability

The full-length structural model of human Rubicon generated with Boltz-2 has been deposited in ModelArchive and is publicly accessible at DOI 10.5452/ma-o1x4b. All other data supporting the findings of this study are provided in the manuscript and its Supplemental Information.

## Notes

### Competing Interest Statement

IMW is an employee of and may hold stocks at GeneDx, Inc. All other authors declare no conflict of interest. We confirm that we have read the Journal's position on issues involved in ethical publication and affirm that this report is consistent with those guidelines.

### Author Declarations

The study was covered by The Research Ethics Committee Institute of Neurology University College London (IoN UCL) (REC 310045)

