## Supplementary Data for "Loss of RUBCN causes autophagy overdrive in a neurodevelopmental disorder with age-dependent neurodegeneration"

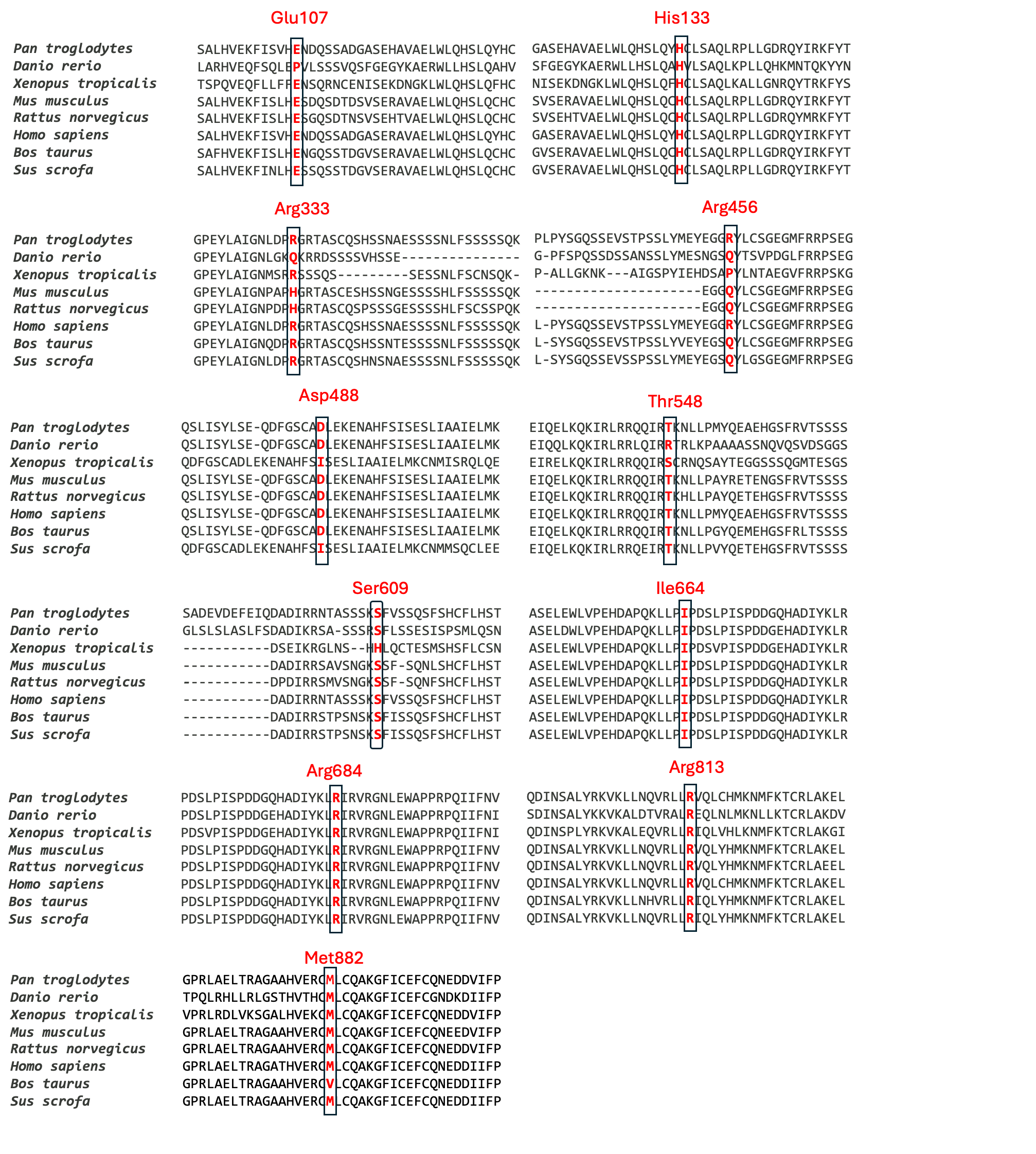
**Supplementary Figure 1**. Analysis of the conservation level of the missense *RUBCN* mutations identified in this study. For each missense variant, a panel of orthologs from different organisms is shown to demonstrate the conservation of the involved residue across species, which suggests that a change of that residue may adversely affect the protein function.

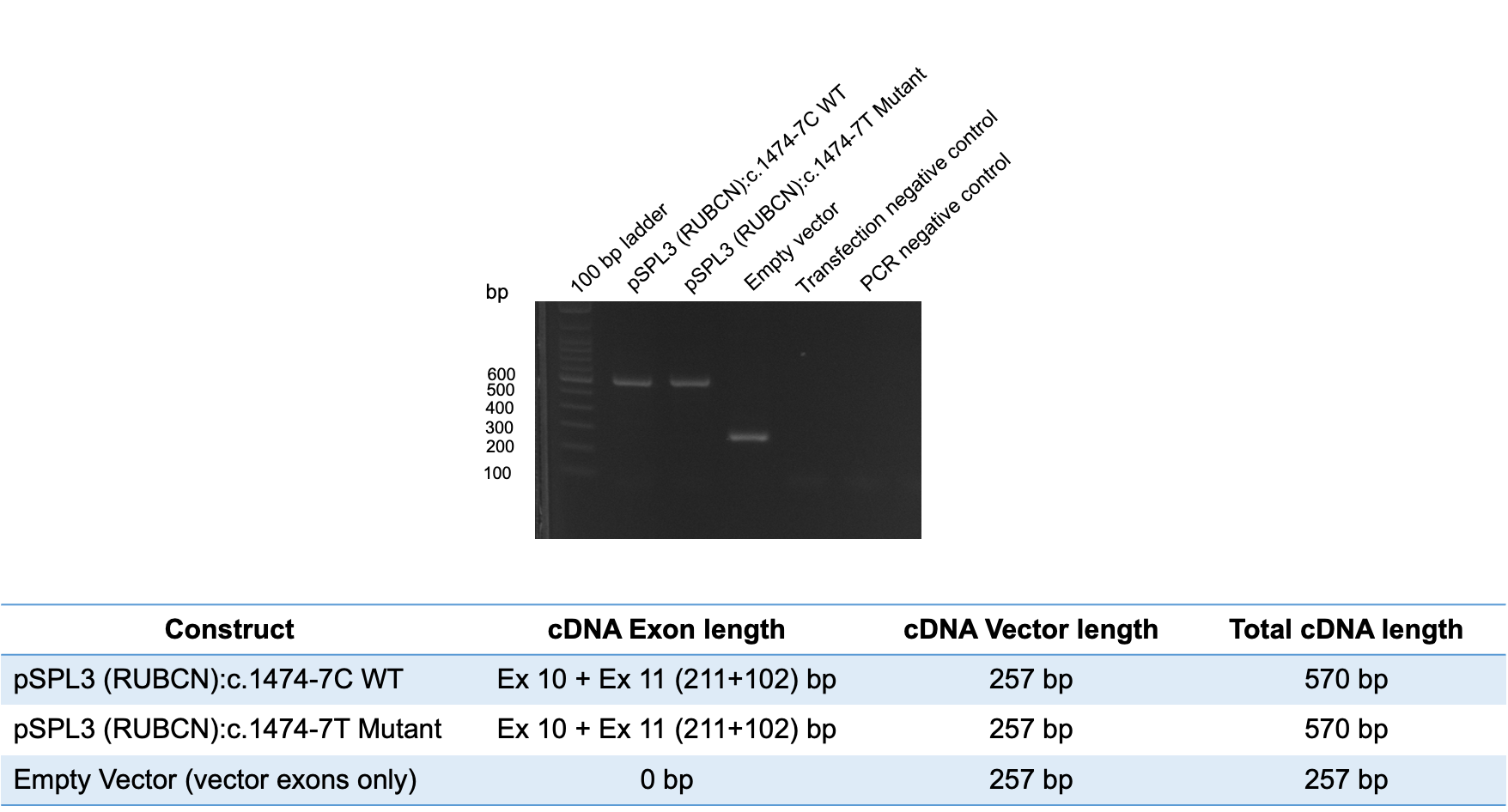

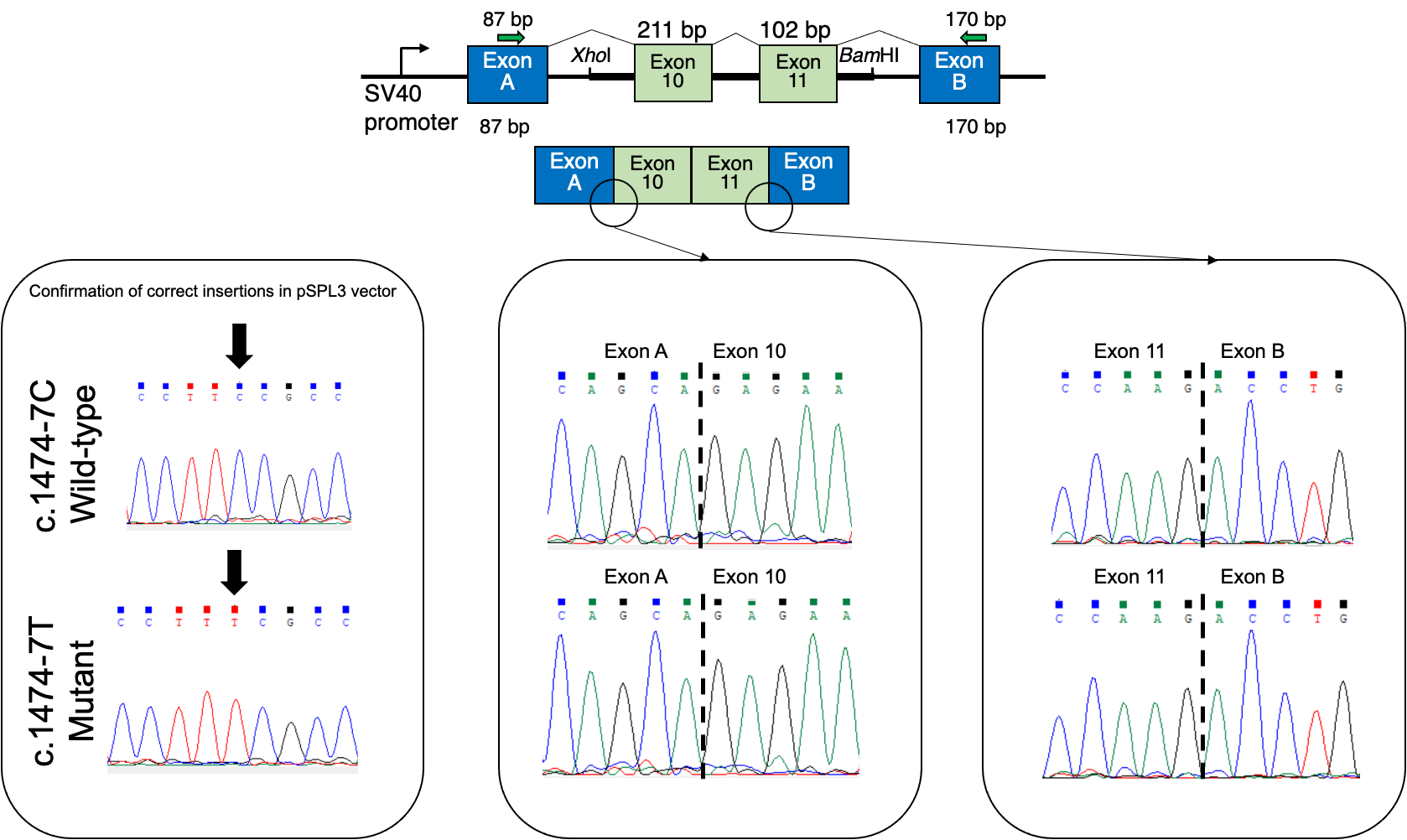

**Supplementary Figure 2. Minigene splicing assay of the *RUBCN* construct (c.1474-7C>T) using the pSPL3 exon-trapping vector.** Agarose gel electrophoresis of RT-PCR products generated from HEK293T cells transfected with the pSPL3-*RUBCN* WT, mutant construct and the empty vector control. The WT construct produced the expected 570-bp transcript, whereas the empty vector generated the 257-bp vector-only product. Schematic representation of the splicing pattern observed in the WT construct, demonstrating normal inclusion of *RUBCN* exons 10 and 11 between the pSPL3 vector exons (SD and SA), resulting in a 570-bp RT-PCR product. Sanger sequencing chromatograms confirming the expected exon–exon junctions and correct inclusion of *RUBCN* exons 10 and 11 in the WT and mutant transcripts.

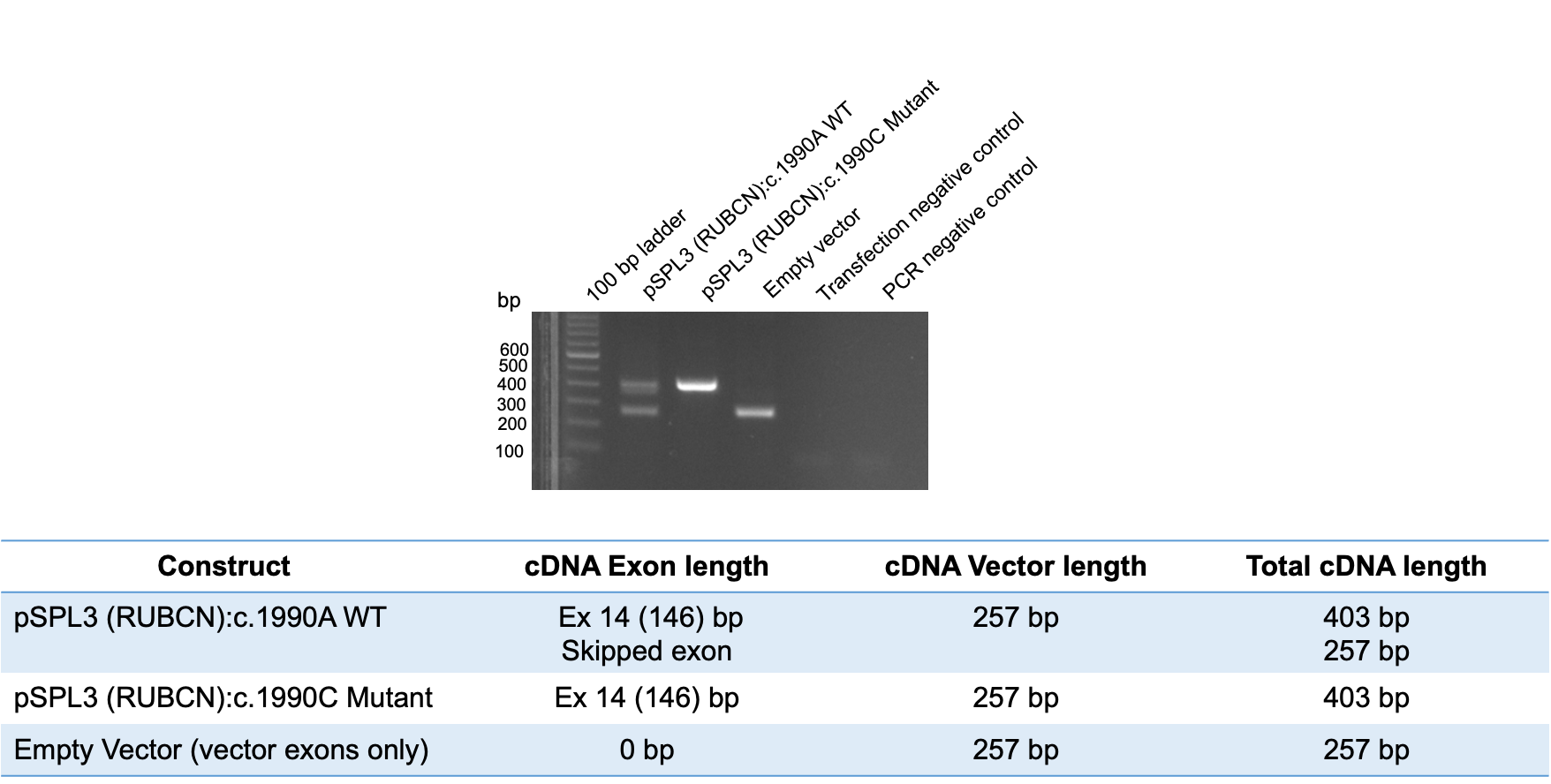

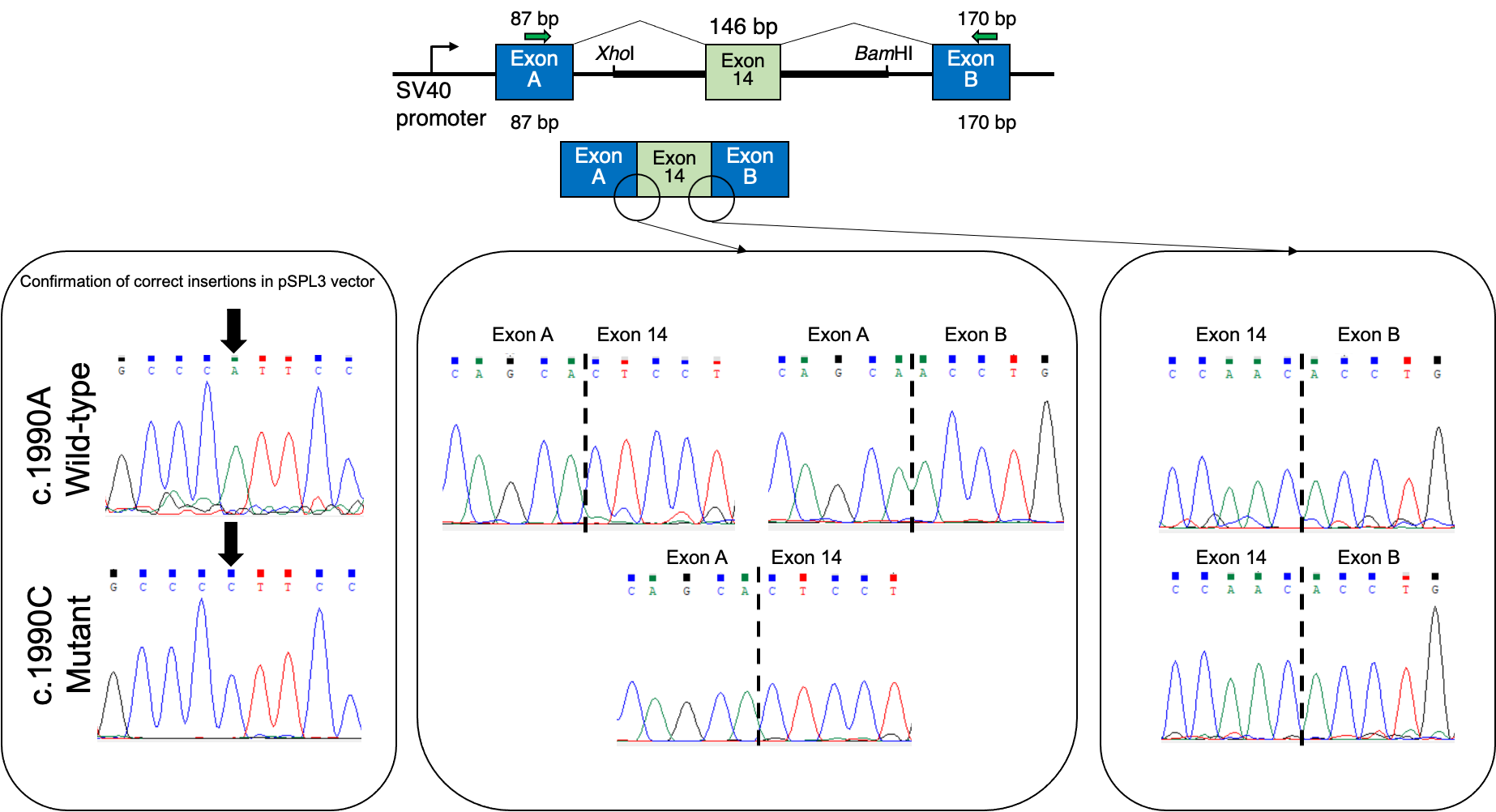

**Supplementary Figure 3. Minigene splicing assay of the *RUBCN* construct (c.1990A>C) using the pSPL3 exon-trapping vector.** Agarose gel electrophoresis of RT-PCR products generated from HEK293T cells transfected with the pSPL3-*RUBCN* WT construct and the empty vector control. The WT construct produced two RT-PCR products: a 403-bp transcript corresponding to normal inclusion of exon 14 and a 257-bp transcript resulting from complete skipping of exon 14. The empty vector generated the expected 257-bp vector-only product. Schematic representation of the two splicing isoforms detected in the WT construct. The upper transcript includes *RUBCN* exon 14 between the pSPL3 vector exons, whereas the lower transcript represents complete skipping of exon 14. Sanger sequencing chromatograms confirming the exon–exon junctions of both splice products. Sequencing verified normal inclusion of exon 14 in the 403-bp transcript and complete exon 14 skipping in the 257-bp transcript.

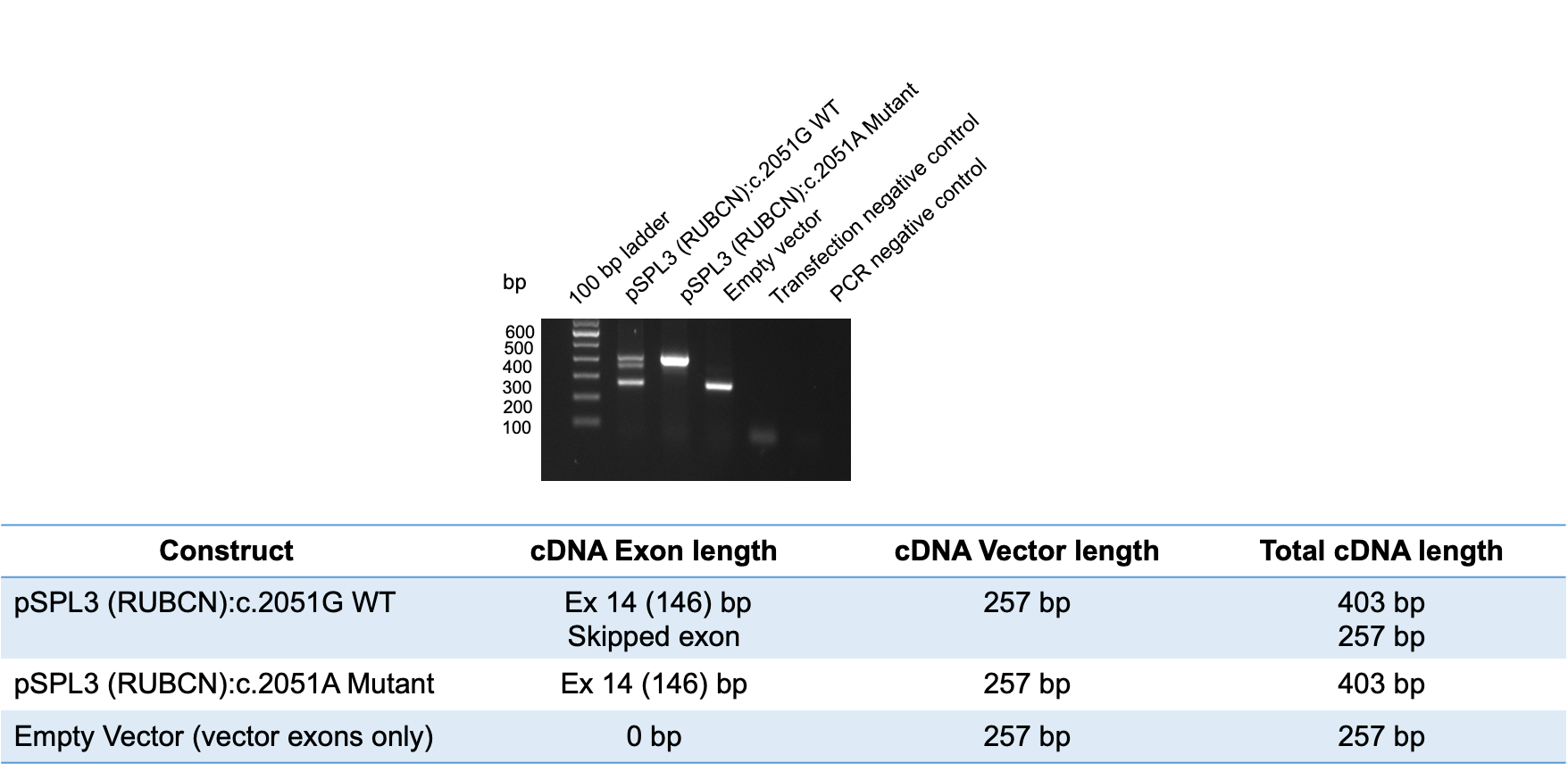

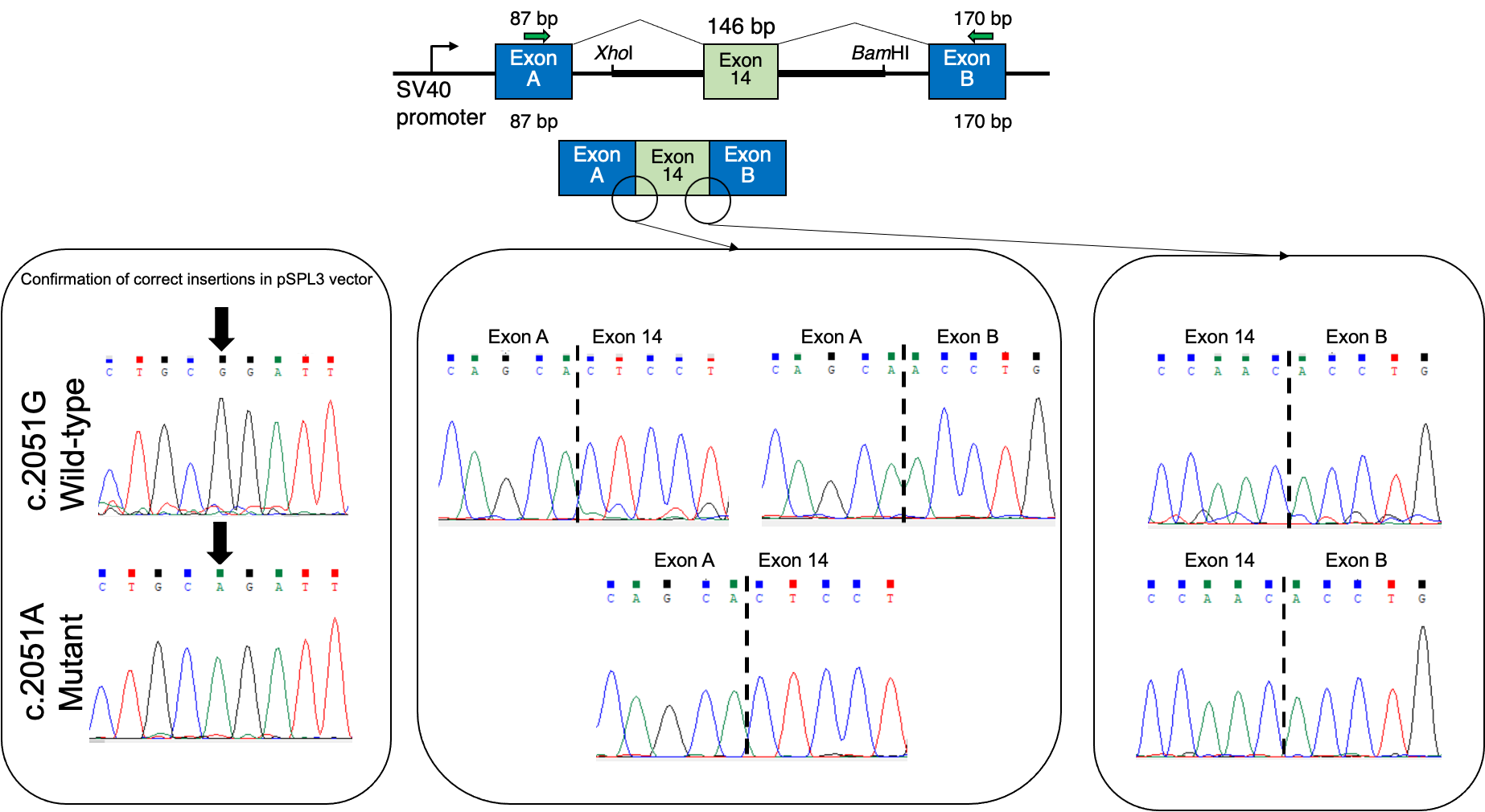

**Supplementary Figure 4. Minigene splicing assay of the *RUBCN* construct (c.2051G>A) using the pSPL3 exon-trapping vector.** Agarose gel electrophoresis of RT-PCR products generated from HEK293T cells transfected with the pSPL3-*RUBCN* WT construct and the empty vector control. The WT construct produced two RT-PCR products: a 403-bp transcript corresponding to normal inclusion of exon 14 and a 257-bp transcript resulting from complete skipping of exon 14. The empty vector generated the expected 257-bp vector-only product. Schematic representation of the two splicing isoforms detected in the WT construct. The upper transcript includes *RUBCN* exon 14 between the pSPL3 vector exons, whereas the lower transcript represents complete skipping of exon 14. Sanger sequencing chromatograms confirming the exon–exon junctions of both splice products. Sequencing verified normal inclusion of exon 14 in the 403-bp transcript and complete exon 14 skipping in the 257-bp transcript.

**Table S1.**

| **REAGENT or RESOURCE** | **SOURCE** | **IDENTIFIER** |
| --- | --- | --- |
| Primary Antibodies |  |  |
| Mouse monoclonal anti-FLAG (M2) | Sigma-Aldrich | F3165 |
| Rabbit monoclonal anti-Rab7 | Cell Signaling Technology | 9367 |
| Secondary Antibodies |  |  |
| Goat-anti-Rabbit(H+L)-HRP | Jackson Immuno Research Laboratories, Inc. | 111-035-003 |
| Goat-anti-Mouse(H+L)-HRP | Jackson Immuno Research Laboratories, Inc. | 115-035-003 |
| Alexa Fluor 488 donkey anti-mouse IgG | Thermo Fisher Scientific | A-21202 |
| Alexa Fluor 568 donkey anti-rabbit IgG | Thermo Fisher Scientific | A-10042 |
| Reagents and Chemicals |  |  |
| Dulbecco's modified Eagle's medium (DMEM) | Sigma-Aldrich | D6429 |
| Fetal bovine serum (FBS) | Sigma-Aldrich | F7524 |
| Penicillin-Streptomycin | Sigma-Aldrich | P4333 |
| L-Glutamine Solution | Sigma-Aldrich | G7513 |
| Trypsin/EDTA | Sigma-Aldrich | T4174 |
| Earle’s Balanced Salts (EBSS) | Sigma-Aldrich | E2888 |
| 4%-Paraformaldehyde Phosphate Buffer Solution | Nakarai Tesque | 09154-85 |
| TransIT-LT1 reagent | Mirus | MIR2300 |
| polyethylenimine (PEI MAX) | Polysciences Inc. | 24765 |
| Opti-MEM reduced serum Medium | Thermo Fisher Scientific | 31985070 |
| polybrene | Sigma Aldrich | H9268 |
| Mounting Medium | VECTASHIELD | H-1000 |
| cOmplete, EDTA-free Protease Inhibitor Cocktail | Roche | 4693132001 |
| Immobilon Forte Western HRP substrate | Merck | WBLUF0500 |
| anti-FLAG M2 agarose beads | Sigma-Aldrich | A2220 |
| In-Fusion HD Cloning Plus | TAKARA Bio | 638909 |
| GEL/PCR purification mini kit | FAVORGEN | FAGCK 001 |
| NucleoSpin Plasmid EasyPure | TAKARA Bio | U0727C |
| NucleoBond　Xtra Midi | TAKARA Bio | U0410B |
| tetramethylrhodamine (TMR)-conjugated ligands | Promega | G8252 |
| Cell lines |  |  |
| HeLa Kyoto | N/A | N/A |
| Plat-E | Dr. Kitamura, Gene Ther. 7, 1063–1066 (2000). |  |
| HEK293T_Halo-LC3 | This study | N/A |
| Plasmids |  |  |
| pLP-VSVG | Tabata K. et al., 2024 | N/A |
| pMRX-ires-puro-HaloTag7-LC3 | Addgene | 184899 |
| pcDNA3.1+ | Thermo Fisher Scientific | V79020 |
| pcDNA3.1+_3xFLAG-RUBCN wt | Bhargava HK et al., 2020 | N/A |
| pcDNA3.1+_3xFLAG-RUBCN CGHL | This study | N/A |
| pcDNA3.1+_3xFLAG-RUBCN E107K | This study | N/A |
| pcDNA3.1+_3xFLAG-RUBCN R466Ter | This study | N/A |
| pcDNA3.1+_3xFLAG-RUBCN T548A | This study | N/A |
| pcDNA3.1+_3xFLAG-RUBCN I664L | This study | N/A |
| pcDNA3.1+_3xFLAG-RUBCN R684Q | This study | N/A |
| pcDNA3.1+_3xFLAG-RUBCN E107K, T548A | This study | N/A |
| pcDNA3.1+_3xFLAG-RUBCN A875fs | This study | N/A |
| pcDNA3.1+_3xFLAG-RUBCN R33Q, R456W | This study | N/A |
| pcDNA3.1+_3xFLAG-RUBCN S609P | This study | N/A |
| pcDNA3.1+_3xFLAG-RUBCN H133Y, D488E | This study | N/A |
| pcDNA3.1+_3xFLAG-RUBCN E519Rfs | This study | N/A |
| Cloning primers |  |  |
| RUBCN: 5’-gcttggatccgaattcATGCGGCCGGAGGGCGCGGG-3’ and 5’-gttcgggcccctcgaggatTCAGGTGGCCTCCAGGACGGC-3’ | Bhargava HK et al., 2020 | N/A |
| Software and Algorithms |  |  |
| Fiji (ImageJ version: 2.0.0-rc-69/1.52n) | N/A | https://fiji.sc |
| Excel version: 16.58 | Microsoft | N/A |
| Prism9 (version: 9.3.1) | GraphPad | https://www.graphpad.com |

**Table S2.** Minigene assay primers

| **Region of Interest** | **Primer Name** | **Primer Sequence 5´- 3´** | **Product Size** |
| --- | --- | --- | --- |
| Construct 1  Exons 3-5 | RUBCN Ex11-12 XhoI F | aattctcgagGAATTCAATCGGGACCTGAC | 1751 bp |
|  | RUBCN Ex11-12 BamHI R | attggatccGGGCTCAAAGAGGCTAAGTC |  |
| Construct 2  Exons 9-10 | RUBCN Ex14 XhoI F | aattctcgagCCAGTCTAAGGGAGGAGTTGG | 1057 bp |
|  | RUBCN Ex14 BamHI R | attggatccGATGGGCTCTGCCCACAT |  |
| pSPL3 Exons A and B | SD6 F | TCTGAGTCACCTGGACAACC | -- |
|  | SA2 R | ATCTCAGTGGTATTTGTGAGC |  |

**Table S3.** Splice Predictions of *RUBCN* variants

| Prediction Tool | c.1474-7C>T | c.1990A>C | c.2051G>A |
| --- | --- | --- | --- |
| SpliceSiteFinder-like | +1.7% | No change | No change |
| MaxEntScan | -11.1% | No change | No change |
| NNSPLICE | +14.8% | -2.2% | No change |
| GeneSplicer | -6.7% | -0.4% | -2.2% |
| SpliceAI 10k  [≥0.2\|0.5\|0.8] | 0.03 AL (-10) | No change | No change |
| AbSplice  [≥0.01\|0.05\|0.2] | No change | No change | No change |

Abbreviation: AL, acceptor loss; predictions show changes in the strengths relative to the native splice sites

**Table S8.** Boltz-2 confidence metrics for the 25 full-length RUBICON-Zn^2+^ diffusion samples, ranked by aggregate confidence score.

| **Rank** | **Model ID^a^** | **Confidence score^b^** | **pTM^c^** | **ipTM**  **(ligand ipTM)ᵈ** | **Complex pLDDT^e^**  **(0-1)** | **Complex PDE^f^ (Å)** |
| --- | --- | --- | --- | --- | --- | --- |
| 1 | 20 | 0.5517 | 0.3581 | 0.7299 | 0.5072 | 4.3592 |
| 2 | 0 | 0.5515 | 0.3641 | 0.7146 | 0.5108 | 4.3180 |
| 3 | 5 | 0.5514 | 0.3610 | 0.7079 | 0.5123 | 4.2992 |
| 4 | 1 | 0.5503 | 0.3617 | 0.7144 | 0.5093 | 4.3314 |
| 5 | 6 | 0.5494 | 0.3642 | 0.7195 | 0.5069 | 4.2943 |
| 6 | 2 | 0.5494 | 0.3725 | 0.7242 | 0.5057 | 4.2928 |
| 7 | 15 | 0.5493 | 0.3632 | 0.7352 | 0.5028 | 4.3146 |
| 8 | 10 | 0.5486 | 0.3700 | 0.7296 | 0.5034 | 4.3001 |
| 9 | 21 | 0.5477 | 0.3647 | 0.7214 | 0.5042 | 4.3066 |
| 10 | 16 | 0.5472 | 0.3519 | 0.6987 | 0.5094 | 4.2929 |
| 11 | 7 | 0.5467 | 0.3720 | 0.7047 | 0.5072 | 4.3009 |
| 12 | 11 | 0.5463 | 0.3703 | 0.7265 | 0.5012 | 4.3143 |
| 13 | 17 | 0.5454 | 0.3621 | 0.7194 | 0.5018 | 4.3777 |
| 14 | 3 | 0.5448 | 0.3619 | 0.7224 | 0.5003 | 4.3022 |
| 15 | 22 | 0.5444 | 0.3614 | 0.7139 | 0.5020 | 4.3389 |
| 16 | 12 | 0.5436 | 0.3676 | 0.6922 | 0.5065 | 4.3171 |
| 17 | 13 | 0.5436 | 0.3546 | 0.6933 | 0.5062 | 4.3650 |
| 18 | 8 | 0.5415 | 0.3625 | 0.7085 | 0.4998 | 4.3781 |
| 19 | 23 | 0.5388 | 0.3658 | 0.6850 | 0.5023 | 4.3465 |
| 20 | 24 | 0.5348 | 0.3637 | 0.6759 | 0.4996 | 4.3204 |
| 21 | 18 | 0.5326 | 0.3619 | 0.6604 | 0.5007 | 4.3515 |
| 22 | 14 | 0.5303 | 0.3583 | 0.6235 | 0.5070 | 4.3667 |
| 23 | 9 | 0.5285 | 0.3498 | 0.6215 | 0.5052 | 4.4094 |
| 24 | 19 | 0.5277 | 0.3552 | 0.6408 | 0.4994 | 4.3708 |
| 25 | 4 | 0.5251 | 0.3594 | 0.6217 | 0.5009 | 4.3782 |

^a^Diffusion-sample identifier assigned by Boltz-2. ^b^Aggregate confidence as reported by Boltz-2, calculated as 0.8 × complex pLDDT + 0.2 × ipTM. ^c^Global predicted template-modelling (pTM) score. ^d^Interface predicted template-modelling (ipTM) score. In all 25 samples ipTM was identical to ligand ipTM and protein ipTM was zero, so this metric reflects the RUBICON-Zn^2+^ interfaces alone. ^e^Mean predicted local accuracy across the complete protein-metal model. ^f^Predicted distance error (PDE) for the complete assembly; smaller values denote greater confidence in the predicted interatomic distances.

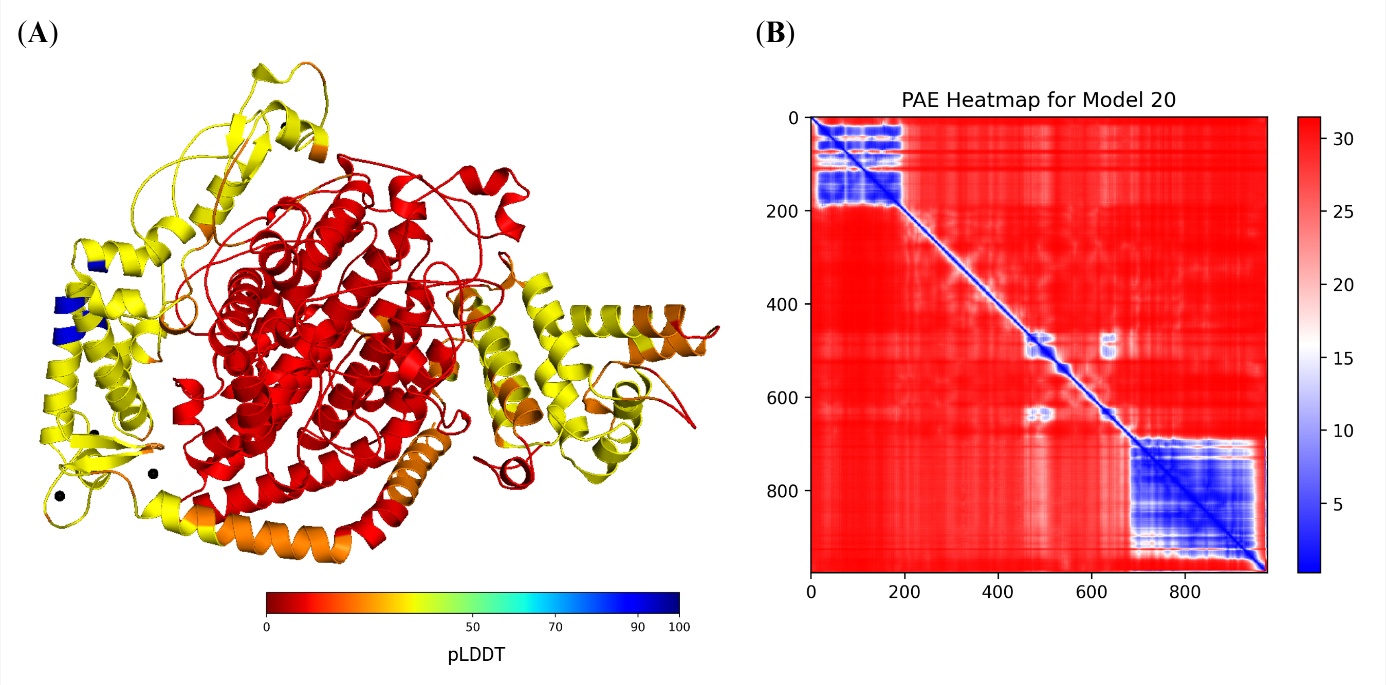

**Figure S5. Confidence profile of Model_20.** (A) The structure coloured by residue-level pLDDT rescaled to the conventional 0-100 range, red marking the least and blue the most confidently predicted positions, with the four Zn^2+^ components drawn as dark-grey spheres. (B) Predicted aligned error (PAE) matrix for the 972 RUBICON residues, in ångströms, low error in blue and high error in red. The matrix is the protein-only submatrix obtained after removal of the rows and columns corresponding to the Zn^2+^ components.

**Table S9.** Local confidence of Model_20 at the 11 missense-variant positions, given as the mean PAE, PDE and pLDDT of the 11-residue window centred on each position. The values describe confidence in the local wild-type (WT) structure surrounding each variant position.

| **Variant position** | **Mean PAE^a^ (Å)** | **Mean PDE^a^ (Å)** | **Mean pLDDT^b^ (0-1)** |
| --- | --- | --- | --- |
| 107 | 5.356 | 1.785 | 0.569 |
| 133 | 1.201 | 0.498 | 0.802 |
| 333 | 7.602 | 2.519 | 0.267 |
| 456 | 8.783 | 2.476 | 0.254 |
| 488 | 4.946 | 1.566 | 0.335 |
| 548 | 3.821 | 1.194 | 0.376 |
| 609 | 7.581 | 2.162 | 0.273 |
| 664 | 7.890 | 2.137 | 0.275 |
| 684 | 7.260 | 1.930 | 0.299 |
| 813 | 0.544 | 0.291 | 0.859 |
| 882 | 1.034 | 0.405 | 0.792 |
| Mean across all variant-centred windows^c^ | 5.093 | 1.542 | 0.464 |

^a^Mean of all 121 elements of the corresponding 11 × 11 PAE or PDE block, including the main diagonal. ^b^Mean residue-level pLDDT across the window, reported on the native 0-1 scale. ^c^Mean of the 11 variant-centred window values.

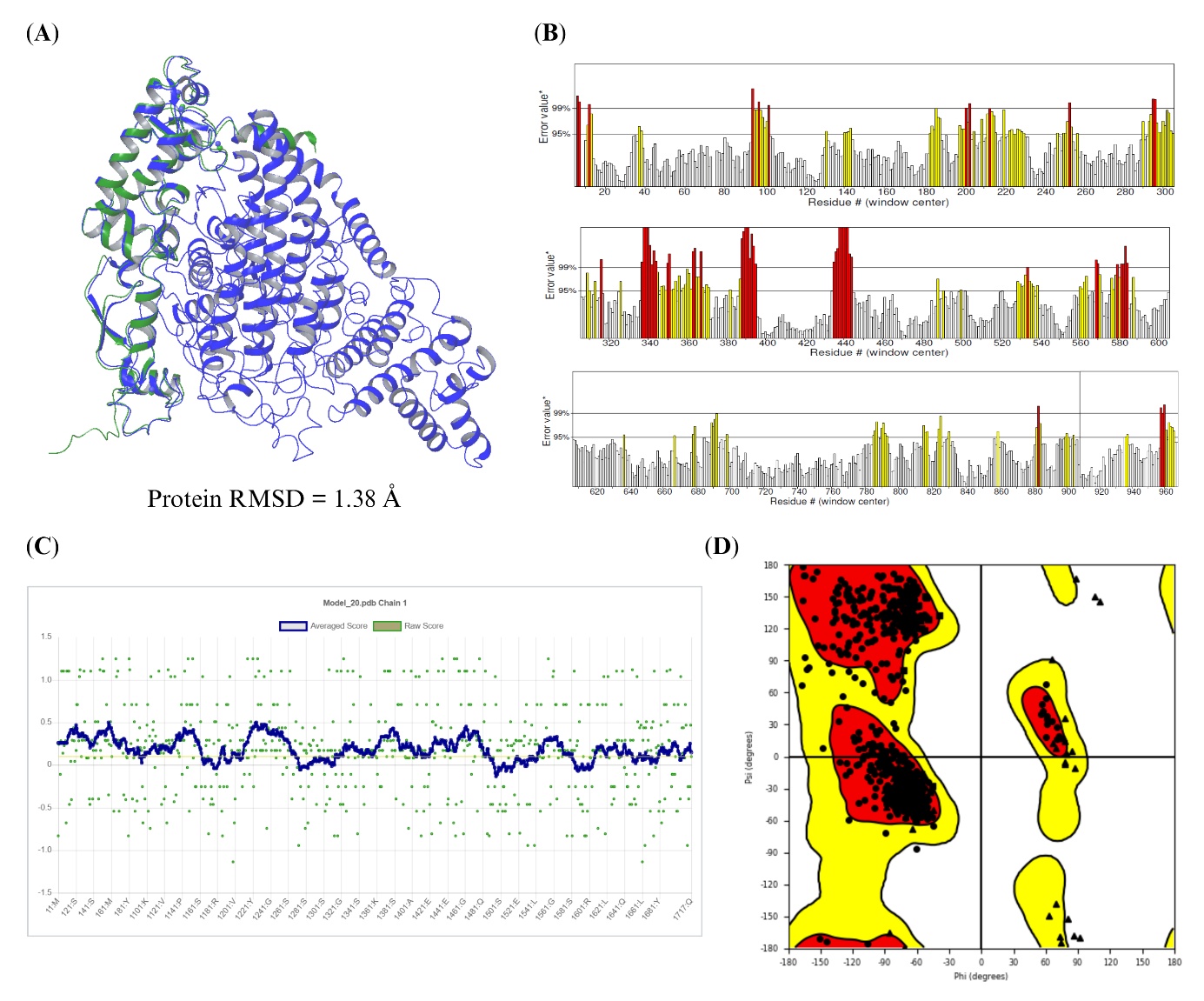

**Figure S6. Model_20 compared with the experimental RUBICON homology (RH) domain and assessed for stereochemical quality.** (A) Backbone superposition on the RUBICON chain of PDB 6WCW, Model_20 in blue and the experimental structure in green, with RAB7AA and the Zn^2+^ ions excluded from both the superposition and the RMSD calculation; the resulting RMSD is given in the panel. (B) Residue-wise ERRAT profile, in which yellow and red bars mark residues above the 95% and 99% rejection thresholds. (C) Verify3D profile, with raw residue 3D-1D scores in green and the window-averaged trace in blue, and the 0.1 acceptance level indicated. (D) PROCHECK Ramachandran plot of backbone φ and ψ angles, with the most favoured, additionally allowed and generously allowed regions shaded.

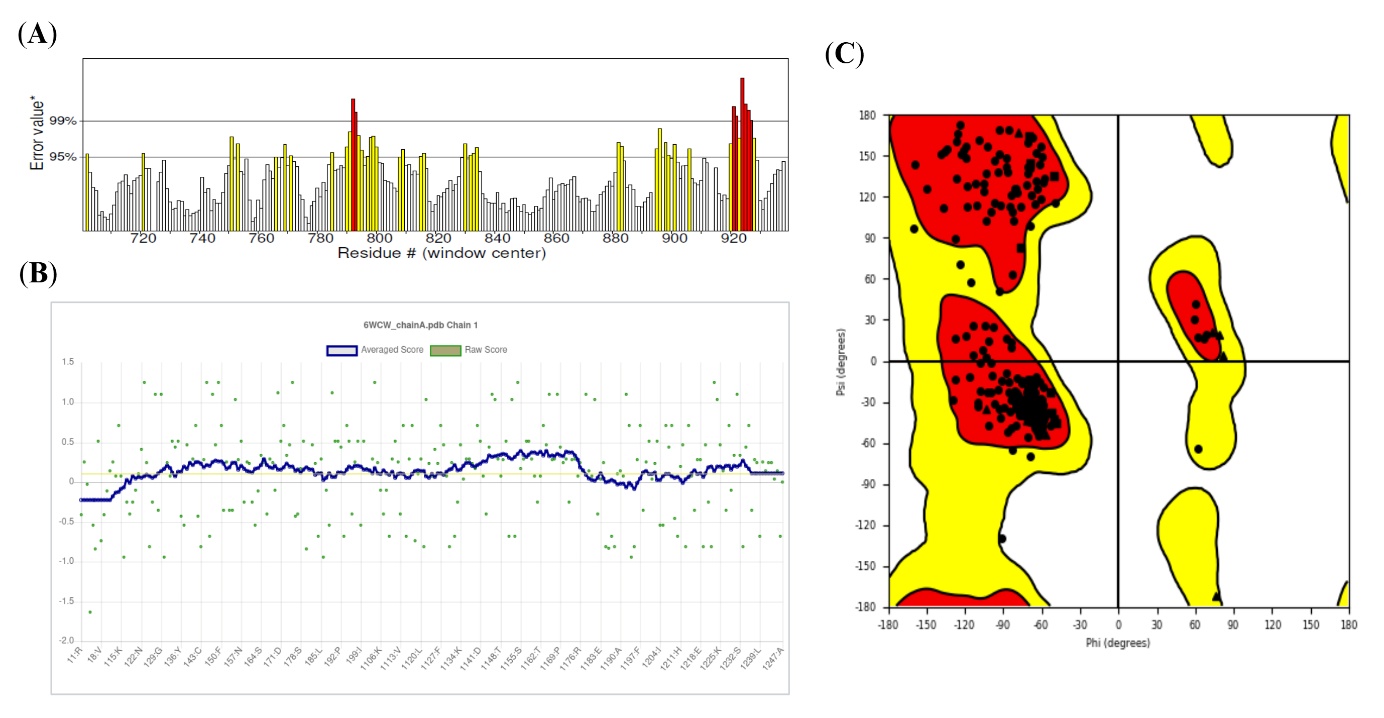

**Figure S7. Quality assessment of the isolated RUBICON chain of PDB 6WCW.** (A) Residue-wise ERRAT profile. (B) Verify3D profile. (C) PROCHECK Ramachandran plot. The three panels use the thresholds, shading and display conventions of Figure S2B-D and cover only the experimentally resolved C-terminal region, so the residue axes span a shorter interval than in the full-length assessments.

**Table S10.** Static Zn^2+^-coordination geometry before and after restrained refinement, comparing prepared Model_20 with Model_1330, the representative structure of the restrained-refinement stage.

| **Zn^2+^ coordination site** | **Coordination motif** | **Coordinating residues** | **Zn^2+^-donor distance rangeᵃ (Å)** | | **Mean Zn^2+^-donor distanceᵃ (Å)** | |
| --- | --- | --- | --- | --- | --- | --- |
|  |  |  | **Model_20** | **Model_1330** | **Model_20** | **Model_1330** |
| B: Zn²⁺ | Cys_4_ | Cys721, Cys724, Cys747, Cys750 | 2.24-2.37 | 2.29-2.48 | 2.31 | 2.37 |
| C: Zn²⁺ | Cys_3_His_1_ | Cys826, His877, Cys881, Cys884 | 2.14-2.35 | 2.20-2.45 | 2.25 | 2.34 |
| D: Zn²⁺ | Cys_3_His_1_ | Cys891, Cys894, His920, Cys923 | 2.15-2.48 | 2.14-2.53 | 2.32 | 2.39 |
| E: Zn²⁺ | Cys_4_ | Cys912, Cys915, Cys929, Cys932 | 2.18-2.30 | 2.27-2.40 | 2.26 | 2.34 |

^a^Measured between each Zn^2+^ ion and its four coordinating donor atoms: cysteine Sγ, His877 Nε2 or His920 Nδ1 as applicable. Values are single-structure measurements, not trajectory averages.

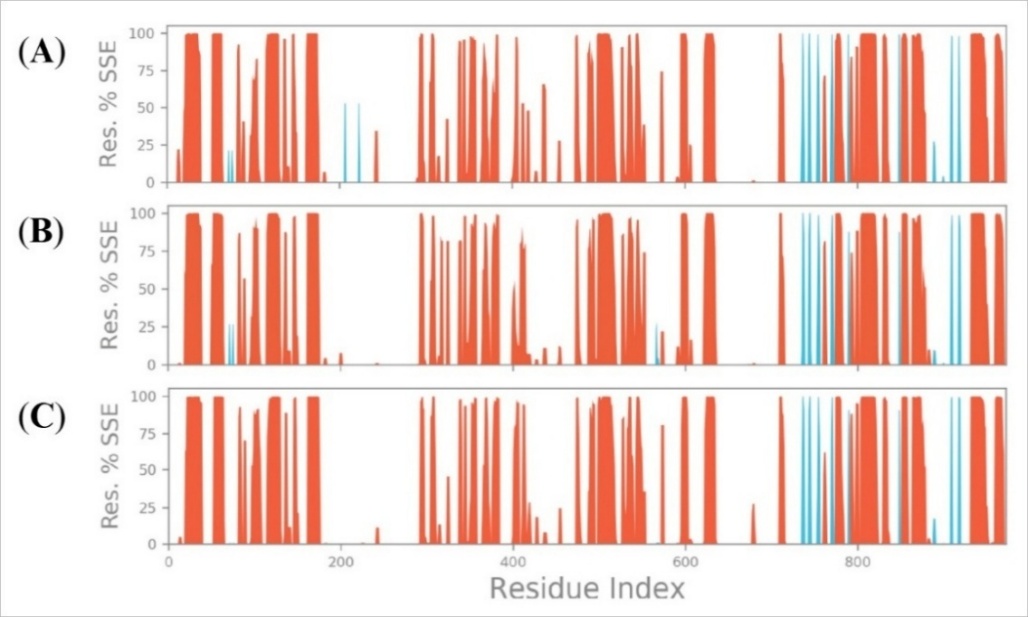

**Figure S8.** **Residue-level secondary-structure occupancy during the three Model_1330 restraint-release simulations.** Panels A to C correspond to replicas 1 to 3, each simulation covering 10 ns with the RUN- and RH domain-directed Cα restraints removed and the temporary Zn^2+^ and donor-atom restraints retained. Orange and blue give the proportion of analysed frames in which each residue adopted α-helical and *β*-strand conformations, plotted against residue number across the full 972-residue sequence.

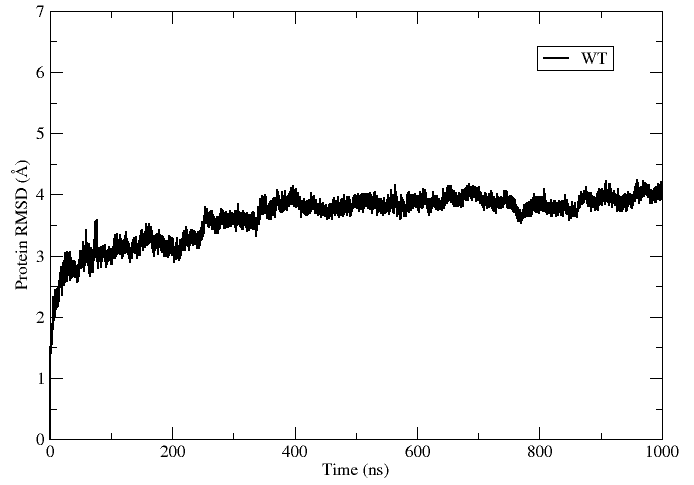

**Figure S9. Protein-backbone RMSD over the 1 μs refinement trajectory initiated from Model_1330.** The trajectory was run without regional Cα restraints, coordinates were recorded every 50 ps, and RMSD was measured against the Model_1330 starting coordinates. The 500 to 1,000 ns portion of the trace is the interval from which the representative structure Model_7247 was subsequently selected. Its identifier is the frame index within that interval.

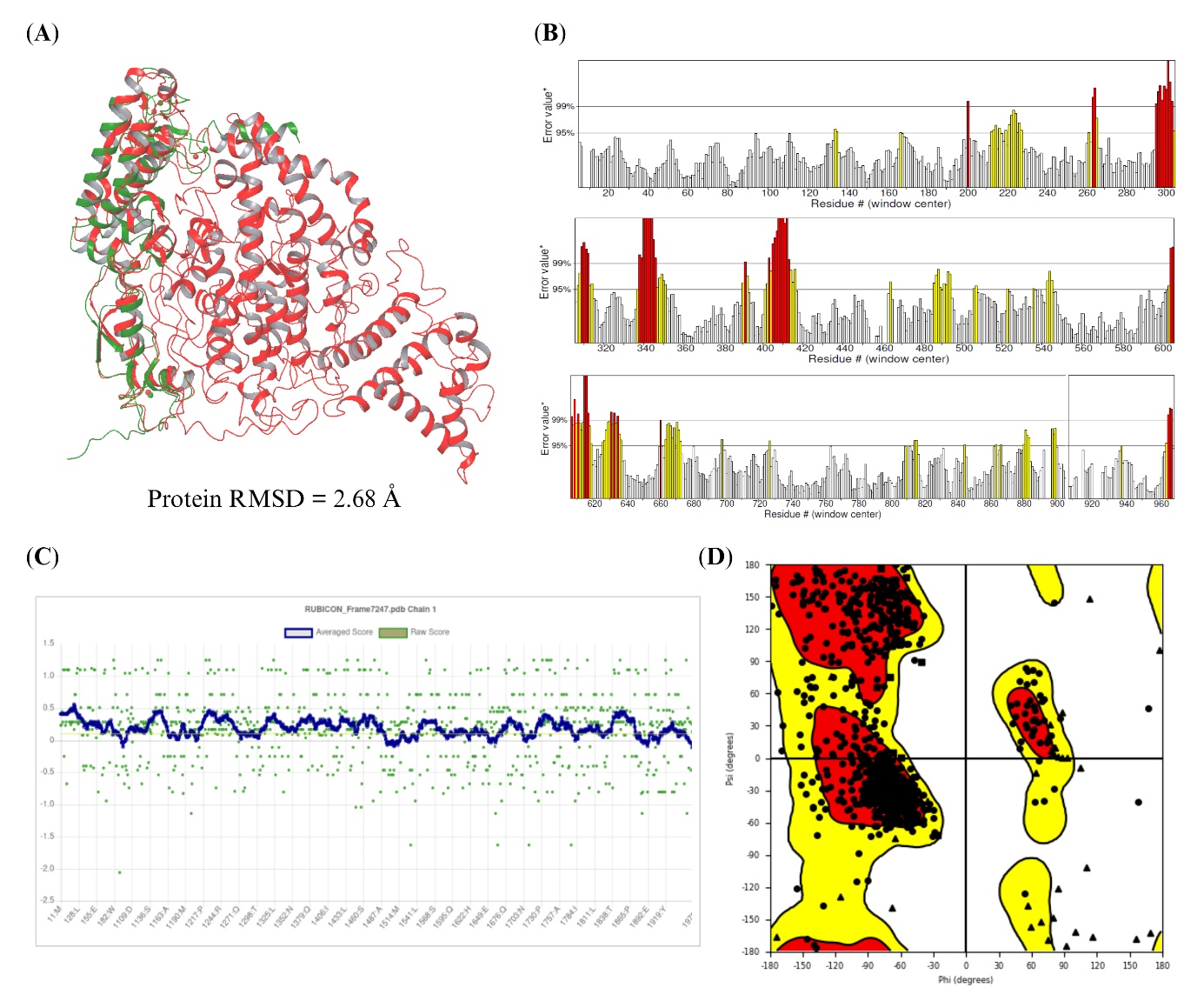

**Figure S10. Model_7247 compared with the experimental RH domain and assessed for stereochemical quality.** (A) Backbone superposition on the RUBICON chain of PDB 6WCW, Model_7247 in red and the experimental structure in green, using the same overlapping residues and the same exclusions as Figure S6A. (B) Residue-wise ERRAT profile. (C) Verify3D profile. (D) PROCHECK Ramachandran plot.

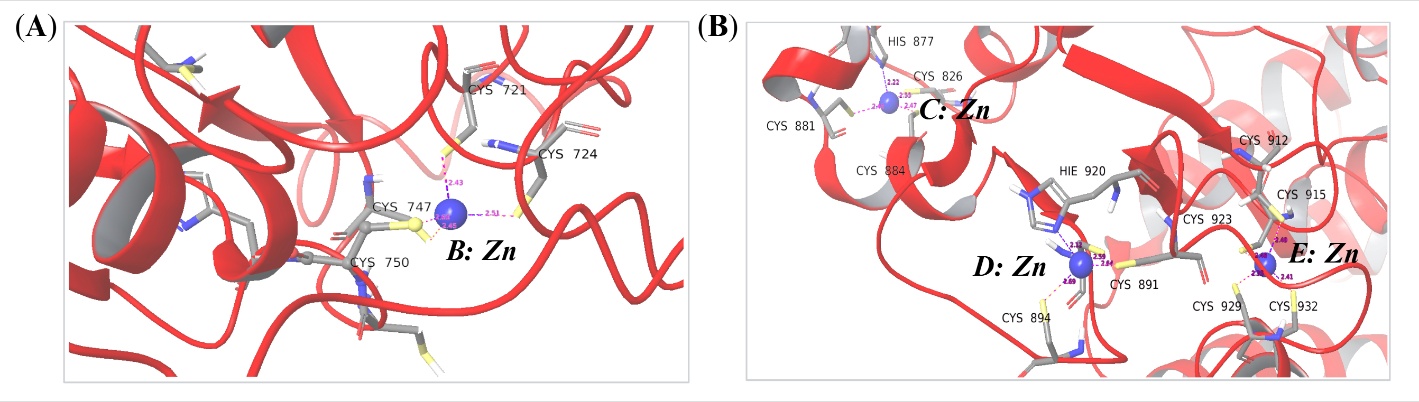

**Figure S11. Zn^2+^-coordination environments in Model_7247.** Enlarged views of (A) site B and (B) the adjacent sites C to E, with Zn^2+^ ions shown as blue spheres and the coordinating cysteine and histidine residues as grey sticks. Labelled dashed lines give the metal-donor distances measured in this single coordinate set, and residue labels identify each donor.

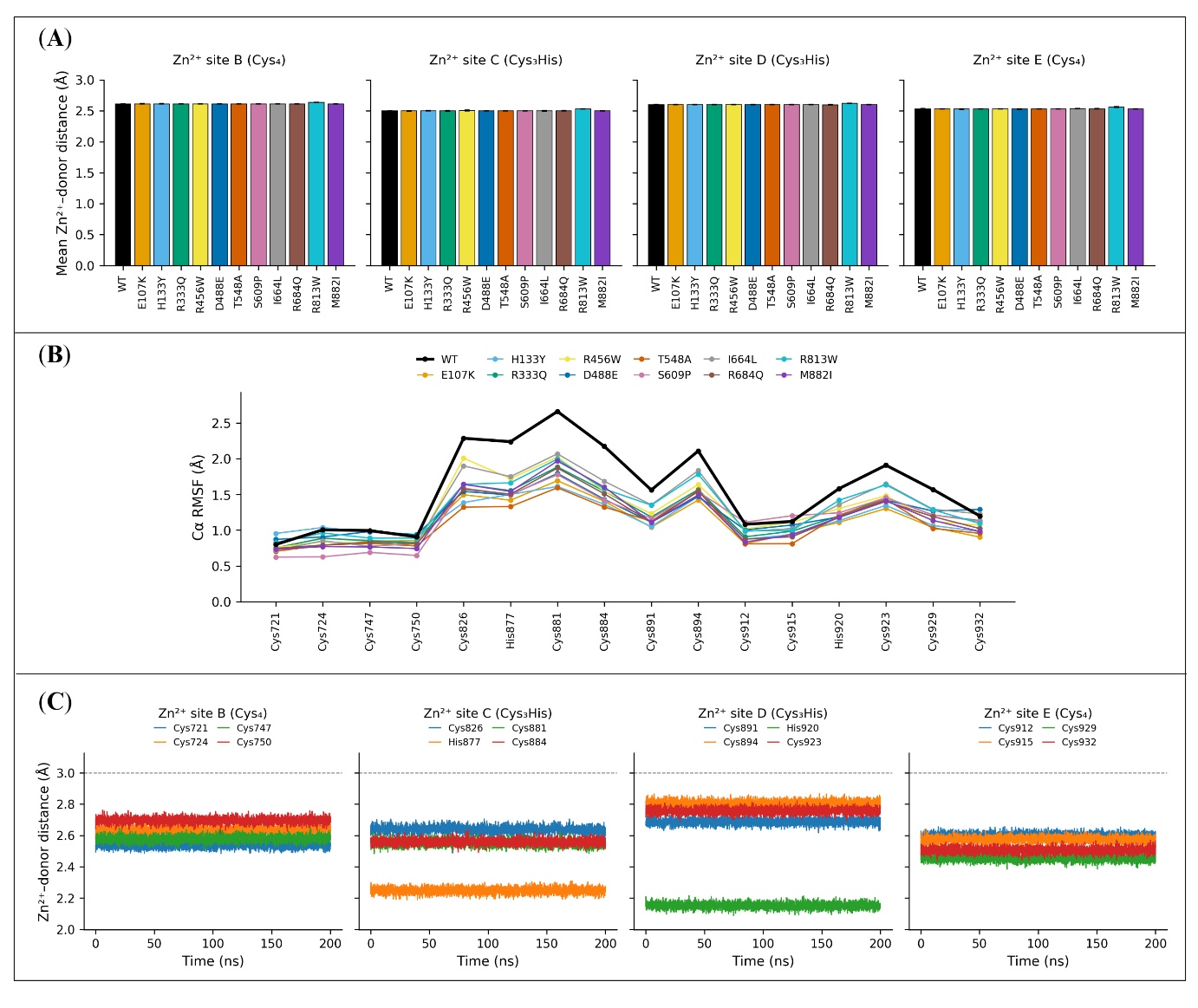

**Figure S12. Zn^2+^-donor distances and coordinating-residue flexibility in all 12 production systems.** (A) Donor distances for sites B to E, each averaged over the four native donors of the site and shown as the mean ± standard deviation (SD) of the three replica-level averages (sample SD; ddof = 1). (B) Replica-averaged Cα RMSF of the 16 coordinating residues over the final 100 ns. (C) Donor-specific distance profiles for WT over the full 200 ns interval, shown as the pointwise mean ± SD across replicas, with a dashed line at the 3.0 Å criterion for a preserved native contact.

**Table S11.** Profile means and between-replica sample SDs for WT RUBICON and 11 missense variants across the whole protein (residues 1-972).

| **System** | **RMSD^a^ (Å)** | **RMSF^a^ (Å)** | **R_g_^a^ (Å)** | **SASA^a^ (Å²)** | **SSC (%)** | | |
| --- | --- | --- | --- | --- | --- | --- | --- |
|  |  |  |  |  | **Helix^b^** | **Sheet^b^** | **Loop^b^** |
| WT | 2.61 ± 0.36 | 1.08 ± 0.21 | 31.83 ± 0.73 | 38949 ± 464 | 34.04 ± 0.13 | 1.75 ± 0.04 | 64.22 ± 0.16 |
| E107K | 2.33 ± 0.18 | 0.98 ± 0.18 | 32.84 ± 1.70 | 38725 ± 506 | 33.20 ± 0.78 | 1.69 ± 0.06 | 65.10 ± 0.82 |
| H133Y | 2.67 ± 0.22 | 1.02 ± 0.21 | 32.22 ± 1.80 | 38821 ± 380 | 33.58 ± 0.85 | 1.61 ± 0.27 | 64.80 ± 0.73 |
| R333Q | 2.30 ± 0.13 | 0.97 ± 0.16 | 32.20 ± 1.67 | 38500 ± 448 | 33.28 ± 0.38 | 1.75 ± 0.03 | 64.97 ± 0.38 |
| R456W | 2.30 ± 0.13 | 1.03 ± 0.21 | 33.00 ± 1.62 | 38804 ± 487 | 34.24 ± 0.74 | 1.88 ± 0.06 | 63.89 ± 0.76 |
| D488E | 2.28 ± 0.25 | 1.06 ± 0.23 | 32.36 ± 1.91 | 38992 ± 582 | 33.97 ± 0.76 | 1.84 ± 0.02 | 64.20 ± 0.75 |
| T548A | 2.10 ± 0.22 | 0.96 ± 0.16 | 32.46 ± 1.55 | 38811 ± 449 | 33.60 ± 1.00 | 1.81 ± 0.10 | 64.59 ± 1.09 |
| S609P | 2.38 ± 0.24 | 1.00 ± 0.21 | 32.07 ± 1.42 | 38923 ± 520 | 34.43 ± 0.37 | 1.77 ± 0.03 | 63.80 ± 0.39 |
| I664L | 2.14 ± 0.25 | 1.00 ± 0.18 | 32.17 ± 1.60 | 38641 ± 477 | 34.01 ± 0.48 | 1.81 ± 0.04 | 64.18 ± 0.51 |
| R684Q | 2.46 ± 0.10 | 0.99 ± 0.19 | 32.88 ± 2.62 | 38628 ± 659 | 34.23 ± 0.39 | 1.79 ± 0.08 | 63.99 ± 0.47 |
| R813W | 2.49 ± 0.29 | 1.05 ± 0.22 | 32.44 ± 1.77 | 39188 ± 932 | 33.87 ± 0.28 | 1.71 ± 0.09 | 64.43 ± 0.29 |
| M882I | 2.34 ± 0.09 | 0.99 ± 0.19 | 32.93 ± 2.17 | 39107 ± 543 | 33.94 ± 0.69 | 1.80 ± 0.08 | 64.27 ± 0.76 |

^a^Mean of the replica-averaged profile followed by the pointwise sample SD across the three replicate profiles (ddof = 1), averaged over the analysis interval. ^b^Mean ± sample SD of the three replica-level averages (ddof = 1). The dispersion in column a quantifies pointwise divergence among the three replica profiles, whereas Table S11 reports the SD across the three replica-level means.

**Table S12.** Profile means and between-replica sample SDs across the RUN domain (residues 49-189).

| **System^a^** | **RMSD^b^ (Å)** | **RMSF^b^ (Å)** | **R_g_^b^ (Å)** | **SASA^b^ (Å²)** | **SSC(%)** | | |
| --- | --- | --- | --- | --- | --- | --- | --- |
|  |  |  |  |  | **Helix^c^** | **Sheet^c,d^** | **Loop^c^** |
| WT | 1.45 ± 0.30 | 1.18 ± 0.11 | 15.44 ± 0.10 | 6221 ± 133 | 49.75 ± 1.31 | < 0.1 | 50.25 ± 1.31 |
| E107K | 1.64 ± 0.21 | 1.14 ± 0.16 | 15.44 ± 0.09 | 6198 ± 149 | 48.29 ± 0.82 | < 0.1 | 51.71 ± 0.82 |
| H133Y | 1.48 ± 0.27 | 1.13 ± 0.19 | 15.36 ± 0.08 | 6122 ± 143 | 47.64 ± 2.59 | < 0.1 | 52.36 ± 2.59 |
| R333Q | 1.79 ± 0.21 | 1.10 ± 0.17 | 15.34 ± 0.09 | 6062 ± 133 | 47.60 ± 1.80 | < 0.1 | 52.39 ± 1.80 |
| R456W | 1.54 ± 0.30 | 1.19 ± 0.24 | 15.39 ± 0.10 | 6154 ± 128 | 49.71 ± 0.91 | < 0.1 | 50.29 ± 0.91 |
| D488E | 1.42 ± 0.36 | 1.21 ± 0.24 | 15.42 ± 0.13 | 6181 ± 164 | 48.99 ± 1.73 | < 0.1 | 51.01 ± 1.73 |
| T548A | 1.65 ± 0.38 | 1.06 ± 0.16 | 15.38 ± 0.10 | 6076 ± 194 | 50.30 ± 0.84 | < 0.1 | 49.70 ± 0.84 |
| S609P | 1.64 ± 0.26 | 1.10 ± 0.18 | 15.34 ± 0.10 | 6159 ± 145 | 50.37 ± 1.91 | < 0.1 | 49.63 ± 1.91 |
| I664L | 1.34 ± 0.13 | 1.05 ± 0.12 | 15.40 ± 0.15 | 6090 ± 154 | 49.17 ± 0.76 | < 0.1 | 50.83 ± 0.76 |
| R684Q | 1.61 ± 0.19 | 1.13 ± 0.17 | 15.34 ± 0.09 | 6106 ± 152 | 48.64 ± 1.42 | < 0.1 | 51.36 ± 1.42 |
| R813W | 1.44 ± 0.28 | 1.16 ± 0.18 | 15.42 ± 0.10 | 6059 ± 176 | 49.11 ± 0.40 | < 0.1 | 50.89 ± 0.40 |
| M882I | 1.32 ± 0.19 | 1.04 ± 0.18 | 15.45 ± 0.07 | 6233 ± 164 | 49.81 ± 0.40 | < 0.1 | 50.19 ± 0.40 |

^a^All 12 systems are listed, although the main text compares only WT and the variants located within this interval. ^b^Mean of the replica-averaged profile followed by the pointwise sample SD across the three replicate profiles (ddof = 1), averaged over the analysis interval. ^c^Mean ± sample SD of the three replica-level averages (ddof = 1). ^d^β-sheet occupancy remained below 0.1% in every system, corresponding to less than one residue of the 141 in this region.

**Table S13.** Profile means and between-replica sample SDs across the central region (residues 190-720).

| **System^a^** | **RMSD^b^ (Å)** | **RMSF^b^ (Å)** | **R_g_^b^ (Å)** | **SASA^b^ (Å²)** | **SSC (%)** | | |
| --- | --- | --- | --- | --- | --- | --- | --- |
|  |  |  |  |  | **Helix^c^** | **Sheet^c,d^** | **Loop^c^** |
| WT | 2.40 ± 0.36 | 0.97 ± 0.18 | 22.96 ± 0.09 | 17983 ± 273 | 27.34 ± 0.57 | < 0.1 | 72.66 ± 0.57 |
| E107K | 2.17 ± 0.13 | 0.91 ± 0.19 | 23.03 ± 0.06 | 17760 ± 283 | 27.95 ± 0.26 | < 0.1 | 72.05 ± 0.26 |
| H133Y | 2.35 ± 0.10 | 0.98 ± 0.23 | 22.99 ± 0.10 | 17926 ± 371 | 28.44 ± 2.19 | < 0.1 | 71.51 ± 2.20 |
| R333Q | 2.12 ± 0.21 | 0.88 ± 0.16 | 22.94 ± 0.05 | 17551 ± 403 | 28.34 ± 1.05 | < 0.1 | 71.63 ± 1.09 |
| R456W | 2.01 ± 0.14 | 0.96 ± 0.22 | 22.97 ± 0.06 | 17946 ± 347 | 28.90 ± 0.04 | < 0.1 | 71.10 ± 0.04 |
| D488E | 2.13 ± 0.09 | 0.98 ± 0.23 | 22.92 ± 0.10 | 17955 ± 340 | 28.74 ± 1.44 | < 0.1 | 71.26 ± 1.44 |
| T548A | 1.94 ± 0.21 | 0.92 ± 0.18 | 22.99 ± 0.09 | 18035 ± 317 | 27.24 ± 1.37 | < 0.1 | 72.74 ± 1.36 |
| S609P | 2.28 ± 0.21 | 0.92 ± 0.21 | 22.98 ± 0.13 | 18036 ± 309 | 28.22 ± 1.42 | < 0.1 | 71.77 ± 1.42 |
| I664L | 1.92 ± 0.17 | 0.92 ± 0.18 | 22.94 ± 0.05 | 17708 ± 329 | 28.71 ± 0.93 | < 0.1 | 71.29 ± 0.93 |
| R684Q | 2.37 ± 0.15 | 0.90 ± 0.19 | 22.90 ± 0.05 | 17693 ± 375 | 29.33 ± 0.71 | < 0.1 | 70.67 ± 0.71 |
| R813W | 2.32 ± 0.26 | 0.97 ± 0.21 | 23.00 ± 0.06 | 18318 ± 565 | 28.09 ± 1.11 | < 0.1 | 71.91 ± 1.11 |
| M882I | 2.29 ± 0.13 | 0.92 ± 0.20 | 23.00 ± 0.12 | 17977 ± 418 | 28.01 ± 0.58 | < 0.1 | 71.97 ± 0.62 |

^a^All 12 systems are listed, although the main text compares only WT and the variants located within this interval. ^b^Mean of the replica-averaged profile followed by the pointwise sample SD across the three replicate profiles (ddof = 1), averaged over the analysis interval. ^c^Mean ± sample SD of the three replica-level averages (ddof = 1). ᵈβ-sheet occupancy remained below 0.1% in every system, corresponding to less than one residue of the 531 in this region.

**Table S14.** Profile means and between-replica sample SDs across the RH domain (residues 721-972).

| **System^a^** | **RMSD^b^ (Å)** | **RMSF^b^ (Å)** | **R_g_^b^ (Å)** | **SASA^b^ (Å²)** | **SSC (%)** | | |
| --- | --- | --- | --- | --- | --- | --- | --- |
|  |  |  |  |  | **Helix^c^** | **Sheet^c^** | **Loop^c^** |
| WT | 2.40 ± 0.46 | 1.29 ± 0.32 | 27.66 ± 0.77 | 12769 ± 319 | 37.12 ± 1.93 | 6.73 ± 0.14 | 56.14 ± 1.87 |
| E107K | 2.14 ± 0.20 | 1.01 ± 0.16 | 27.20 ± 0.39 | 12782 ± 357 | 33.99 ± 2.87 | 6.53 ± 0.23 | 59.48 ± 3.02 |
| H133Y | 2.12 ± 0.18 | 1.05 ± 0.17 | 26.66 ± 0.45 | 12749 ± 220 | 34.56 ± 3.44 | 6.12 ± 0.98 | 59.32 ± 4.41 |
| R333Q | 2.12 ± 0.25 | 1.12 ± 0.16 | 28.28 ± 1.78 | 12924 ± 261 | 33.74 ± 0.64 | 6.68 ± 0.03 | 59.59 ± 0.68 |
| R456W | 2.13 ± 0.27 | 1.09 ± 0.15 | 27.10 ± 0.46 | 12649 ± 270 | 34.73 ± 2.29 | 7.24 ± 0.24 | 58.02 ± 2.41 |
| D488E | 2.16 ± 0.37 | 1.15 ± 0.25 | 27.92 ± 1.64 | 12821 ± 324 | 34.82 ± 0.63 | 7.07 ± 0.08 | 58.11 ± 0.58 |
| T548A | 1.77 ± 0.29 | 1.01 ± 0.13 | 27.02 ± 0.53 | 12801 ± 174 | 35.58 ± 3.52 | 6.94 ± 0.34 | 57.48 ± 3.79 |
| S609P | 1.87 ± 0.24 | 1.07 ± 0.18 | 27.79 ± 1.36 | 12715 ± 274 | 36.70 ± 2.37 | 6.81 ± 0.09 | 56.49 ± 2.41 |
| I664L | 2.06 ± 0.39 | 1.16 ± 0.25 | 27.20 ± 0.19 | 12923 ± 248 | 35.00 ± 0.72 | 6.96 ± 0.14 | 58.04 ± 0.85 |
| R684Q | 2.18 ± 0.37 | 1.07 ± 0.18 | 27.14 ± 0.45 | 12855 ± 361 | 34.25 ± 1.94 | 6.88 ± 0.32 | 58.87 ± 2.01 |
| R813W | 2.27 ± 0.24 | 1.17 ± 0.26 | 26.87 ± 0.28 | 12819 ± 293 | 35.67 ± 2.20 | 6.57 ± 0.33 | 57.76 ± 2.48 |
| M882I | 2.20 ± 0.23 | 1.07 ± 0.17 | 27.66 ± 0.86 | 12893 ± 197 | 35.39 ± 1.24 | 6.87 ± 0.26 | 57.74 ± 1.36 |

^a^All 12 systems are listed, although the main text compares only WT and the variants located within this interval. ^b^Mean of the replica-averaged profile followed by the pointwise sample SD across the three replicate profiles (ddof = 1), averaged over the analysis interval. ^c^Mean ± sample SD of the three replica-level averages (ddof = 1). For RMSD, RMSF, R_g_ and SASA, the dispersion reported here quantifies pointwise divergence among the three replica profiles. The main text and Table S18 instead report the sample SD across the three replica-level means; the two quantities describe different sources of variation and are not expected to be identical.

**Table S15.** Temporal and sequence dependence of the analysed profiles and the number of profile points retained after dependence-aware subsampling.

| **Region** | **Observable** | **N^a^** | **tau_int (samples)ᵇ** | **tau_int (ns)ᵇ** | ***g*ᶜ** | **n_sub_ (WT)** | **n_sub_ (variants)** |
| --- | --- | --- | --- | --- | --- | --- | --- |
| Whole protein | RMSD | 2001 | 87.11 | 4.36 | 711.10 | 3 | 3-11 |
|  | RMSF | 972 | 12.83 | n.a.ᵇ | 25.70 | 38 | 49-81 |
|  | R_g_ | 201 | 7.07 | 3.54 | 14.10 | 15 | 9-67 |
|  | SASA | 201 | 7.60 | 3.80 | 15.20 | 14 | 6-101 |
| RUN domain | RMSD | 2001 | 49.28 | 2.46 | 364.40 | 6 | 7-8 |
|  | RMSF | 141 | 9.70 | n.a.ᵇ | 19.40 | 8 | 6-13 |
|  | R_g_ | 201 | 9.95 | 4.97 | 19.90 | 11 | 13-17 |
|  | SASA | 201 | 10.04 | 5.02 | 20.10 | 11 | 10-41 |
| Central region | RMSD | 2001 | 81.25 | 4.06 | 617.30 | 4 | 3-5 |
|  | RMSF | 531 | 5.49 | n.a.ᵇ | 11.00 | 49 | 28-54 |
|  | R_g_ | 201 | 3.63 | 1.82 | 7.30 | 29 | 8-51 |
|  | SASA | 201 | 23.37 | 11.68 | 46.70 | 5 | 6-51 |
| RH domain | RMSD | 2001 | 91.65 | 4.58 | 760.30 | 3 | 8-65 |
|  | RMSF | 252 | 7.24 | n.a.ᵇ | 14.50 | 18 | 18-23 |
|  | R_g_ | 201 | 15.05 | 7.53 | 30.10 | 7 | 5-34 |
|  | SASA | 201 | 4.07 | 2.03 | 8.10 | 26 | 6-9 |

^a^Stored frames for RMSD, R_g_ and SASA; residue positions for Cα RMSF. ^b^Not applicable to Cα RMSF, whose lag variable is sequence separation in residues. For RMSF, the retained-point count refers to residue-profile points after sequence-order thinning and is not a count of independent observations. ^c^For RMSD, *g* was derived from a τint estimate taken over a wider lag range than the value tabulated here. The final two columns give the number of points actually retained after integer-stride subsampling, which differs from the continuous ratio *N*/*g* through stride rounding and endpoint inclusion.

**Table S16.** Distributional and variance test results for the 88 WT-versus-variant comparisons. The tests compare replica-averaged profiles and are exploratory; they do not constitute independent-trajectory-level inference.

| **Region** | **Observable** | **Variant^a^** | **KS D** | **KS *q*^b^** | **MWU *q*^b^** | **AD A^2^** | **AD *q*^b^** | **BF F** | **BF *q*^b^** |
| --- | --- | --- | --- | --- | --- | --- | --- | --- | --- |
| Whole | RMSD | E107K | 0.667 | 0.427 | 0.393 | 1.532 | 0.172 | 2.541 | 0.266 |
| Whole | RMSD | H133Y | 0.667 | 0.427 | 0.489 | 1.321 | 0.172 | 2.524 | 0.266 |
| Whole | RMSD | R333Q | 0.667 | 0.449 | 0.458 | 0.712 | 0.250 | 1.453 | 0.346 |
| Whole | RMSD | R456W | 0.667 | 0.449 | 0.393 | 1.322 | 0.172 | 1.737 | 0.346 |
| Whole | RMSD | D488E | 0.667 | 0.723 | 0.489 | 0.031 | 0.250 | 0.119 | 0.775 |
| Whole | RMSD | T548A | 1.000 | 0.183 | 0.183 | 4.230 | 0.073 | 4.751 | 0.266 |
| Whole | RMSD | S609P | 0.500 | 0.723 | 0.489 | -0.237 | 0.250 | 0.091 | 0.775 |
| Whole | RMSD | I664L | 1.000 | 0.314 | 0.314 | 3.122 | 0.096 | 1.446 | 0.346 |
| Whole | RMSD | R684Q | 0.556 | 0.625 | 0.530 | 0.129 | 0.250 | 2.696 | 0.266 |
| Whole | RMSD | R813W | 0.394 | 0.725 | 0.769 | 0.248 | 0.250 | 3.725 | 0.266 |
| Whole | RMSD | M882I | 0.667 | 0.427 | 0.458 | 1.309 | 0.172 | 4.412 | 0.266 |
| Whole | RMSF | E107K | 0.127 | 0.872 | 0.876 | -0.524 | 0.250 | 0.137 | 0.828 |
| Whole | RMSF | H133Y | 0.127 | 0.872 | 0.978 | -0.724 | 0.250 | 0.100 | 0.828 |
| Whole | RMSF | R333Q | 0.222 | 0.797 | 0.876 | 1.029 | 0.250 | 1.077 | 0.668 |
| Whole | RMSF | R456W | 0.118 | 0.872 | 0.978 | -0.882 | 0.250 | 0.681 | 0.668 |
| Whole | RMSF | D488E | 0.119 | 0.872 | 0.978 | -0.928 | 0.250 | 0.847 | 0.668 |
| Whole | RMSF | T548A | 0.145 | 0.872 | 0.876 | -0.223 | 0.250 | 0.004 | 0.951 |
| Whole | RMSF | S609P | 0.128 | 0.872 | 0.978 | -0.710 | 0.250 | 0.641 | 0.668 |
| Whole | RMSF | I664L | 0.218 | 0.797 | 0.876 | -0.224 | 0.250 | 0.187 | 0.828 |
| Whole | RMSF | R684Q | 0.237 | 0.797 | 0.876 | 0.919 | 0.250 | 1.423 | 0.668 |
| Whole | RMSF | R813W | 0.165 | 0.872 | 0.905 | -0.517 | 0.250 | 1.209 | 0.668 |
| Whole | RMSF | M882I | 0.165 | 0.872 | 0.876 | 0.058 | 0.250 | 0.792 | 0.668 |

**Table S16.** (continued)

| **Region** | **Observable** | **Variant^a^** | **KS D** | **KS *q*^b^** | **MWU *q*^b^** | **AD A^2^** | **AD *q*^b^** | **BF F** | **BF *q*^b^** |
| --- | --- | --- | --- | --- | --- | --- | --- | --- | --- |
| Whole | R_g_ | E107K | 0.591 | 5.39e^-04^ | 0.002 | 9.362 | 0.003 | 16.339 | 3.33e^-04^ |
| Whole | R_g_ | H133Y | 0.412 | 0.049 | 0.538 | 3.601 | 0.018 | 23.408 | 1.60e^-04^ |
| Whole | R_g_ | R333Q | 0.408 | 0.041 | 0.280 | 3.309 | 0.020 | 17.107 | 3.33e^-04^ |
| Whole | R_g_ | R456W | 0.688 | 4.10e^-05^ | 3.19e^-05^ | 15.063 | 0.003 | 14.163 | 7.00e^-04^ |
| Whole | R_g_ | D488E | 0.586 | 0.004 | 0.032 | 5.565 | 0.005 | 7.994 | 0.008 |
| Whole | R_g_ | T548A | 0.400 | 0.184 | 0.681 | 2.512 | 0.033 | 12.814 | 0.002 |
| Whole | R_g_ | S609P | 0.403 | 0.041 | 0.914 | 2.613 | 0.033 | 9.352 | 0.004 |
| Whole | R_g_ | I664L | 0.532 | 0.004 | 0.051 | 4.477 | 0.010 | 13.551 | 8.82e^-04^ |
| Whole | R_g_ | R684Q | 0.606 | 4.78e^-04^ | 0.004 | 8.519 | 0.003 | 18.016 | 3.22e^-04^ |
| Whole | R_g_ | R813W | 0.444 | 0.182 | 0.626 | 1.012 | 0.125 | 4.990 | 0.036 |
| Whole | R_g_ | M882I | 0.608 | 0.002 | 0.004 | 6.878 | 0.003 | 14.120 | 8.82e^-04^ |
| Whole | SASA | E107K | 0.527 | 0.006 | 0.014 | 8.590 | 0.004 | 6.792 | 0.040 |
| Whole | SASA | H133Y | 0.357 | 0.784 | 0.202 | 0.666 | 0.241 | 0.015 | 0.958 |
| Whole | SASA | R333Q | 0.571 | 0.182 | 0.044 | 3.129 | 0.048 | 0.164 | 0.958 |
| Whole | SASA | R456W | 0.429 | 0.323 | 0.143 | 1.632 | 0.126 | 0.003 | 0.958 |
| Whole | SASA | D488E | 0.333 | 0.793 | 0.779 | -0.240 | 0.250 | 0.004 | 0.958 |
| Whole | SASA | T548A | 0.412 | 0.135 | 0.144 | 2.230 | 0.087 | 2.788 | 0.224 |
| Whole | SASA | S609P | 0.286 | 0.793 | 0.589 | -0.426 | 0.250 | 0.032 | 0.958 |
| Whole | SASA | I664L | 0.617 | 3.08e^-04^ | 0.002 | 13.935 | 0.004 | 10.486 | 0.014 |
| Whole | SASA | R684Q | 0.786 | 3.08e^-04^ | 0.007 | 7.598 | 0.004 | 10.984 | 0.014 |
| Whole | SASA | R813W | 0.308 | 0.323 | 0.230 | 1.223 | 0.160 | 3.498 | 0.184 |
| Whole | SASA | M882I | 0.214 | 0.981 | 0.699 | -0.888 | 0.250 | 0.126 | 0.958 |

**Table S16.** (continued)

| **Region** | **Observable** | **Variant^a^** | **KS D** | **KS *q*^b^** | **MWU *q*^b^** | **AD A^2^** | **AD *q*^b^** | **BF F** | **BF *q*^b^** |
| --- | --- | --- | --- | --- | --- | --- | --- | --- | --- |
| RUN domain | RMSD | E107K | 0.714 | 0.077 | 0.070 | 2.572 | 0.057 | 0.774 | 0.783 |
| RUN domain | RMSD | H133Y | 0.542 | 0.192 | 0.228 | 0.271 | 0.250 | 0.079 | 0.783 |
| RUN domain | RMSF | E107K | 0.231 | 0.898 | 1.000 | -0.935 | 0.250 | 0.026 | 0.874 |
| RUN domain | RMSF | H133Y | 0.333 | 0.898 | 1.000 | -0.546 | 0.250 | 0.737 | 0.815 |
| RUN domain | R_g_ | E107K | 0.364 | 0.324 | 0.487 | 0.347 | 0.240 | 3.901 | 0.061 |
| RUN domain | R_g_ | H133Y | 0.610 | 0.016 | 0.004 | 6.074 | 0.003 | 5.207 | 0.061 |
| RUN domain | SASA | E107K | 0.224 | 0.686 | 0.622 | -0.291 | 0.250 | 3.016 | 0.089 |
| RUN domain | SASA | H133Y | 0.636 | 0.022 | 0.076 | 2.625 | 0.055 | 4.921 | 0.078 |
| Central region | RMSD | R333Q | 1.000 | 0.080 | 0.080 | 4.175 | 0.024 | 0.052 | 0.876 |
| Central region | RMSD | R456W | 1.000 | 0.080 | 0.080 | 3.122 | 0.024 | 0.185 | 0.876 |
| Central region | RMSD | D488E | 1.000 | 0.080 | 0.080 | 3.122 | 0.024 | 0.758 | 0.876 |
| Central region | RMSD | T548A | 1.000 | 0.080 | 0.080 | 3.122 | 0.024 | 0.027 | 0.876 |
| Central region | RMSD | S609P | 0.667 | 0.467 | 0.267 | 0.741 | 0.190 | 0.798 | 0.876 |
| Central region | RMSD | I664L | 1.000 | 0.080 | 0.080 | 3.687 | 0.024 | 6.104 | 0.339 |
| Central region | RMSD | R684Q | 0.250 | 1.000 | 1.000 | -1.024 | 0.250 | 0.159 | 0.876 |
| Central region | RMSF | R333Q | 0.267 | 0.096 | 0.162 | 2.950 | 0.071 | 0.120 | 0.992 |
| Central region | RMSF | R456W | 0.190 | 0.306 | 0.364 | 0.777 | 0.183 | 0.073 | 0.992 |
| Central region | RMSF | D488E | 0.316 | 0.096 | 0.293 | 2.031 | 0.083 | 0.545 | 0.992 |
| Central region | RMSF | T548A | 0.226 | 0.265 | 0.293 | 1.159 | 0.152 | 0.002 | 0.992 |
| Central region | RMSF | S609P | 0.265 | 0.096 | 0.162 | 2.233 | 0.083 | 9.68e^-05^ | 0.992 |
| Central region | RMSF | I664L | 0.182 | 0.386 | 0.332 | 0.372 | 0.234 | 0.003 | 0.992 |
| Central region | RMSF | R684Q | 0.441 | 0.005 | 0.015 | 7.099 | 0.007 | 1.49e^-04^ | 0.992 |

**Table S16.** (continued)

| **Region** | **Observable** | **Variant^a^** | **KS D** | **KS *q*^b^** | **MWU *q*^b^** | **AD A^2^** | **AD *q*^b^** | **BF F** | **BF *q*^b^** |
| --- | --- | --- | --- | --- | --- | --- | --- | --- | --- |
| Central region | R_g_ | R333Q | 0.375 | 0.308 | 0.360 | 0.667 | 0.245 | 1.962 | 0.660 |
| Central region | R_g_ | R456W | 0.226 | 0.797 | 0.918 | -0.720 | 0.250 | 0.050 | 0.824 |
| Central region | R_g_ | D488E | 0.591 | 0.031 | 0.004 | 6.178 | 0.003 | 0.199 | 0.824 |
| Central region | R_g_ | T548A | 0.509 | 0.020 | 0.002 | 7.528 | 0.003 | 0.225 | 0.824 |
| Central region | R_g_ | S609P | 0.280 | 0.635 | 0.765 | -0.358 | 0.250 | 0.575 | 0.824 |
| Central region | R_g_ | I664L | 0.322 | 0.058 | 0.005 | 5.229 | 0.005 | 0.091 | 0.824 |
| Central region | R_g_ | R684Q | 0.826 | 1.67e^-07^ | 5.33e^-07^ | 19.681 | 0.003 | 1.781 | 0.660 |
| Central region | SASA | R333Q | 1.000 | 6.88e^-04^ | 3.44e^-04^ | 8.454 | 0.004 | 1.093 | 0.497 |
| Central region | SASA | R456W | 0.300 | 0.838 | 0.708 | -0.663 | 0.250 | 1.040 | 0.497 |
| Central region | SASA | D488E | 0.633 | 0.311 | 0.220 | 1.083 | 0.204 | 0.099 | 0.760 |
| Central region | SASA | T548A | 0.366 | 0.555 | 0.823 | -0.621 | 0.250 | 0.645 | 0.497 |
| Central region | SASA | S609P | 0.429 | 0.555 | 0.823 | -0.424 | 0.250 | 1.633 | 0.497 |
| Central region | SASA | I664L | 0.824 | 0.004 | 3.44e^-04^ | 10.072 | 0.004 | 0.748 | 0.497 |
| Central region | SASA | R684Q | 0.800 | 0.017 | 0.004 | 5.919 | 0.004 | 0.766 | 0.497 |
| RH domain | RMSD | R813W | 0.451 | 0.927 | 1.000 | 1.859 | 0.111 | 24.925 | 9.24e^-06^ |
| RH domain | RMSD | M882I | 0.333 | 0.927 | 1.000 | -0.260 | 0.250 | 2.137 | 0.178 |
| RH domain | RMSF | R813W | 0.333 | 0.275 | 0.351 | 0.193 | 0.250 | 1.327 | 0.515 |
| RH domain | RMSF | M882I | 0.350 | 0.254 | 0.351 | 0.218 | 0.250 | 0.033 | 0.856 |
| RH domain | R_g_ | R813W | 1.000 | 0.005 | 0.005 | 5.657 | 0.004 | 2.516 | 0.288 |
| RH domain | R_g_ | M882I | 0.307 | 0.533 | 0.647 | -0.551 | 0.250 | 0.259 | 0.613 |
| RH domain | SASA | R813W | 0.389 | 0.211 | 0.086 | 1.301 | 0.095 | 1.873 | 0.361 |
| RH domain | SASA | M882I | 0.756 | 0.006 | 0.024 | 6.144 | 0.003 | 0.525 | 0.474 |

^a^All 11 variants are listed for the complete protein. Regional rows list only the variants located within the interval concerned, that is two in the RUN domain, seven in the central region and two in the RH domain. These groupings define the sets within which *P* values were adjusted separately for each test. ^b^KS, Kolmogorov-Smirnov; MWU, Mann-Whitney U; AD, Anderson-Darling; BF, Brown-Forsythe. A Benjamini-Hochberg (BH)-adjusted *q*-value is the smallest false-discovery rate (FDR) at which the comparison would be declared significant within its family. For RMSF, the four tests compare marginal RMSF-value distributions after sequence-lag subsampling. They do not preserve same-residue WT-mutant pairing and are reported only as exploratory profile descriptors.

**Table S17.** Profile-level effect magnitudes, moving-block bootstrap confidence intervals (CIs) and exploratory adjusted values for all 88 predefined WT-versus-variant comparisons.

| **Region** | **Observable** | **Variant^a^** | **Δ**  **(variant - WT)^b^** | **95% CI^c^** | **WT**  **between-replica scale^b^** | **\|Δ\| / scale^d^** | **KS *q*^e^** | **MWU *q*^e^** | **AD *q*^e^** | **BF *q*^e^** |
| --- | --- | --- | --- | --- | --- | --- | --- | --- | --- | --- |
| Whole | RMSD | E107K | -0.29 | -0.43 to -0.16 | 0.36 | 0.81 | 0.427 | 0.393 | 0.172 | 0.266 |
| Whole | RMSD | H133Y | +0.06 | -0.07 to 0.22 | 0.36 | 0.17 | 0.427 | 0.489 | 0.172 | 0.266 |
| Whole | RMSD | R333Q | -0.31 | -0.47 to -0.18 | 0.36 | 0.86 | 0.449 | 0.458 | 0.250 | 0.346 |
| Whole | RMSD | R456W | -0.32 | -0.48 to -0.18 | 0.36 | 0.89 | 0.449 | 0.393 | 0.172 | 0.346 |
| Whole | RMSD | D488E | -0.33 | -0.52 to -0.16 | 0.36 | 0.92 | 0.723 | 0.489 | 0.250 | 0.775 |
| Whole | RMSD | T548A | -0.51 | -0.67 to -0.39 | 0.36 | 1.42 | 0.183 | 0.183 | 0.073 | 0.266 |
| Whole | RMSD | S609P | -0.23 | -0.40 to -0.07 | 0.36 | 0.64 | 0.723 | 0.489 | 0.250 | 0.775 |
| Whole | RMSD | I664L | -0.47 | -0.64 to -0.34 | 0.36 | 1.31 | 0.314 | 0.314 | 0.096 | 0.346 |
| Whole | RMSD | R684Q | -0.16 | -0.33 to -0.05 | 0.36 | 0.44 | 0.625 | 0.530 | 0.250 | 0.266 |
| Whole | RMSD | R813W | -0.12 | -0.25 to 0.02 | 0.36 | 0.33 | 0.725 | 0.769 | 0.250 | 0.266 |
| Whole | RMSD | M882I | -0.27 | -0.41 to -0.14 | 0.36 | 0.75 | 0.427 | 0.458 | 0.172 | 0.266 |
| Whole | RMSF | E107K | -0.10 | -0.24 to 0.01 | 0.21 | 0.48 | 0.872 | 0.876 | 0.250 | 0.828 |
| Whole | RMSF | H133Y | -0.06 | -0.20 to 0.07 | 0.21 | 0.29 | 0.872 | 0.978 | 0.250 | 0.828 |
| Whole | RMSF | R333Q | -0.11 | -0.24 to 0.03 | 0.21 | 0.52 | 0.797 | 0.876 | 0.250 | 0.668 |
| Whole | RMSF | R456W | -0.05 | -0.20 to 0.08 | 0.21 | 0.24 | 0.872 | 0.978 | 0.250 | 0.668 |
| Whole | RMSF | D488E | -0.02 | -0.17 to 0.11 | 0.21 | 0.10 | 0.872 | 0.978 | 0.250 | 0.668 |
| Whole | RMSF | T548A | -0.12 | -0.25 to 0.00 | 0.21 | 0.57 | 0.872 | 0.876 | 0.250 | 0.951 |
| Whole | RMSF | S609P | -0.08 | -0.22 to 0.03 | 0.21 | 0.38 | 0.872 | 0.978 | 0.250 | 0.668 |
| Whole | RMSF | I664L | -0.08 | -0.22 to 0.06 | 0.21 | 0.38 | 0.797 | 0.876 | 0.250 | 0.828 |
| Whole | RMSF | R684Q | -0.09 | -0.24 to 0.03 | 0.21 | 0.43 | 0.797 | 0.876 | 0.250 | 0.668 |
| Whole | RMSF | R813W | -0.03 | -0.17 to 0.10 | 0.21 | 0.14 | 0.872 | 0.905 | 0.250 | 0.668 |
| Whole | RMSF | M882I | -0.09 | -0.23 to 0.03 | 0.21 | 0.43 | 0.872 | 0.876 | 0.250 | 0.668 |

**Table S17.** (continued)

| **Region** | **Observable** | **Variant^a^** | **Δ**  **(variant - WT)^b^** | **95% CI^c^** | **WT**  **between-replica scale^b^** | **\|Δ\| / scale^c^** | **KS *q*^d^** | **MWU *q*^d^** | **AD *q*^d^** | **BF *q*^d^** |
| --- | --- | --- | --- | --- | --- | --- | --- | --- | --- | --- |
| Whole | R_g_ | E107K | +1.01 | 0.65 to 1.33 | 0.73 | 1.38 | 5.39e^−04^ | 0.002 | 0.003 | 3.33e^−04^ |
| Whole | R_g_ | H133Y | +0.40 | -0.01 to 0.80 | 0.73 | 0.55 | 0.049 | 0.538 | 0.018 | 1.60e^−04^ |
| Whole | R_g_ | R333Q | +0.38 | 0.07 to 0.71 | 0.73 | 0.52 | 0.041 | 0.280 | 0.020 | 3.33e^−04^ |
| Whole | R_g_ | R456W | +1.17 | 0.90 to 1.50 | 0.73 | 1.60 | 4.10e^−05^ | 3.19e^−05^ | 0.003 | 7.00e^−04^ |
| Whole | R_g_ | D488E | +0.53 | 0.18 to 1.04 | 0.73 | 0.73 | 0.004 | 0.032 | 0.005 | 0.008 |
| Whole | R_g_ | T548A | +0.63 | 0.23 to 1.13 | 0.73 | 0.86 | 0.184 | 0.681 | 0.033 | 0.002 |
| Whole | R_g_ | S609P | +0.25 | -0.02 to 0.52 | 0.73 | 0.34 | 0.041 | 0.914 | 0.033 | 0.004 |
| Whole | R_g_ | I664L | +0.34 | 0.03 to 0.63 | 0.73 | 0.47 | 0.004 | 0.051 | 0.010 | 8.82e^−04^ |
| Whole | R_g_ | R684Q | +1.05 | 0.70 to 1.42 | 0.73 | 1.44 | 4.78e^−04^ | 0.004 | 0.003 | 3.22e^−04^ |
| Whole | R_g_ | R813W | +0.61 | 0.13 to 1.19 | 0.73 | 0.84 | 0.182 | 0.626 | 0.125 | 0.036 |
| Whole | R_g_ | M882I | +1.11 | 0.63 to 1.59 | 0.73 | 1.52 | 0.002 | 0.004 | 0.003 | 8.82e^−04^ |
| Whole | SASA | E107K | -224 | -330 to -154 | 464 | 0.48 | 0.006 | 0.014 | 0.004 | 0.040 |
| Whole | SASA | H133Y | -128 | -312 to 10 | 464 | 0.28 | 0.784 | 0.202 | 0.241 | 0.958 |
| Whole | SASA | R333Q | -449 | -630 to -327 | 464 | 0.97 | 0.182 | 0.044 | 0.048 | 0.958 |
| Whole | SASA | R456W | -146 | -304 to -1 | 464 | 0.31 | 0.323 | 0.143 | 0.126 | 0.958 |
| Whole | SASA | D488E | +43 | -123 to 313 | 464 | 0.09 | 0.793 | 0.779 | 0.250 | 0.958 |
| Whole | SASA | T548A | -138 | -249 to -49 | 464 | 0.30 | 0.135 | 0.144 | 0.087 | 0.224 |
| Whole | SASA | S609P | -26 | -192 to 121 | 464 | 0.06 | 0.793 | 0.589 | 0.250 | 0.958 |
| Whole | SASA | I664L | -308 | -408 to -238 | 464 | 0.66 | 3.08e^−04^ | 0.002 | 0.004 | 0.014 |
| Whole | SASA | R684Q | -321 | -442 to -238 | 464 | 0.69 | 3.08e^−04^ | 0.007 | 0.004 | 0.014 |
| Whole | SASA | R813W | +239 | 144 to 331 | 464 | 0.52 | 0.323 | 0.230 | 0.160 | 0.184 |
| Whole | SASA | M882I | +158 | 7 to 344 | 464 | 0.34 | 0.981 | 0.699 | 0.250 | 0.958 |

**Table S17.** (continued)

| **Region** | **Observable** | **Variant^a^** | **Δ**  **(variant - WT)^b^** | **95% CI^c^** | **WT**  **between-replica scale^b^** | **\|Δ\| / scale^c^** | **KS *q*^d^** | **MWU *q*^d^** | **AD *q*^d^** | **BF *q*^d^** |
| --- | --- | --- | --- | --- | --- | --- | --- | --- | --- | --- |
| RUN domain | RMSD | E107K | +0.19 | 0.12 to 0.30 | 0.30 | 0.63 | 0.077 | 0.070 | 0.057 | 0.783 |
| RUN domain | RMSD | H133Y | +0.03 | -0.04 to 0.12 | 0.30 | 0.10 | 0.192 | 0.228 | 0.250 | 0.783 |
| RUN domain | RMSF | E107K | -0.04 | -0.48 to 0.37 | 0.11 | 0.36 | 0.898 | 1.000 | 0.250 | 0.874 |
| RUN domain | RMSF | H133Y | -0.05 | -0.52 to 0.32 | 0.11 | 0.45 | 0.898 | 1.000 | 0.250 | 0.815 |
| RUN domain | R_g_ | E107K | +0.00 | -0.02 to 0.03 | 0.10 | 0.00 | 0.324 | 0.487 | 0.240 | 0.061 |
| RUN domain | R_g_ | H133Y | -0.08 | -0.10 to -0.04 | 0.10 | 0.80 | 0.016 | 0.004 | 0.003 | 0.061 |
| RUN domain | SASA | E107K | -22 | -64 to 21 | 133 | 0.17 | 0.686 | 0.622 | 0.250 | 0.089 |
| RUN domain | SASA | H133Y | -99 | -148 to -48 | 133 | 0.74 | 0.022 | 0.076 | 0.055 | 0.078 |
| Central region | RMSD | R333Q | -0.29 | -0.36 to -0.22 | 0.36 | 0.81 | 0.080 | 0.080 | 0.024 | 0.876 |
| Central region | RMSD | R456W | -0.39 | -0.48 to -0.31 | 0.36 | 1.08 | 0.080 | 0.080 | 0.024 | 0.876 |
| Central region | RMSD | D488E | -0.28 | -0.39 to -0.16 | 0.36 | 0.78 | 0.080 | 0.080 | 0.024 | 0.876 |
| Central region | RMSD | T548A | -0.46 | -0.55 to -0.39 | 0.36 | 1.28 | 0.080 | 0.080 | 0.024 | 0.876 |
| Central region | RMSD | S609P | -0.12 | -0.22 to -0.00 | 0.36 | 0.33 | 0.467 | 0.267 | 0.190 | 0.876 |
| Central region | RMSD | I664L | -0.48 | -0.57 to -0.40 | 0.36 | 1.33 | 0.080 | 0.080 | 0.024 | 0.339 |
| Central region | RMSD | R684Q | -0.04 | -0.11 to 0.02 | 0.36 | 0.11 | 1.000 | 1.000 | 0.250 | 0.876 |
| Central region | RMSF | R333Q | -0.09 | -0.20 to 0.04 | 0.18 | 0.50 | 0.096 | 0.162 | 0.071 | 0.992 |
| Central region | RMSF | R456W | -0.01 | -0.12 to 0.11 | 0.18 | 0.06 | 0.306 | 0.364 | 0.183 | 0.992 |
| Central region | RMSF | D488E | +0.01 | -0.13 to 0.16 | 0.18 | 0.06 | 0.096 | 0.293 | 0.083 | 0.992 |
| Central region | RMSF | T548A | -0.05 | -0.17 to 0.07 | 0.18 | 0.28 | 0.265 | 0.293 | 0.152 | 0.992 |
| Central region | RMSF | S609P | -0.04 | -0.15 to 0.06 | 0.18 | 0.22 | 0.096 | 0.162 | 0.083 | 0.992 |
| Central region | RMSF | I664L | -0.04 | -0.16 to 0.08 | 0.18 | 0.22 | 0.386 | 0.332 | 0.234 | 0.992 |
| Central region | RMSF | R684Q | -0.07 | -0.20 to 0.08 | 0.18 | 0.39 | 0.005 | 0.015 | 0.007 | 0.992 |

**Table S17.** (continued)

| **Region** | **Observable** | **Variant^a^** | **Δ**  **(variant - WT)^b^** | **95% CI^c^** | **WT**  **between-replica scale^b^** | **\|Δ\| / scale^c^** | **KS *q*^d^** | **MWU *q*^d^** | **AD *q*^d^** | **BF *q*^d^** |
| --- | --- | --- | --- | --- | --- | --- | --- | --- | --- | --- |
| Central region | R_g_ | R333Q | -0.02 | -0.04 to -0.01 | 0.09 | 0.22 | 0.308 | 0.360 | 0.245 | 0.660 |
| Central region | R_g_ | R456W | +0.00 | -0.01 to 0.02 | 0.09 | 0.00 | 0.797 | 0.918 | 0.250 | 0.824 |
| Central region | R_g_ | D488E | -0.04 | -0.06 to -0.02 | 0.09 | 0.44 | 0.031 | 0.004 | 0.003 | 0.824 |
| Central region | R_g_ | T548A | +0.03 | 0.02 to 0.04 | 0.09 | 0.33 | 0.020 | 0.002 | 0.003 | 0.824 |
| Central region | R_g_ | S609P | +0.01 | 0.01 to 0.03 | 0.09 | 0.11 | 0.635 | 0.765 | 0.250 | 0.824 |
| Central region | R_g_ | I664L | -0.02 | -0.03 to -0.01 | 0.09 | 0.22 | 0.058 | 0.005 | 0.005 | 0.824 |
| Central region | R_g_ | R684Q | -0.06 | -0.07 to -0.05 | 0.09 | 0.67 | 1.67e^−07^ | 5.33e^−07^ | 0.003 | 0.660 |
| Central region | SASA | R333Q | -432 | -636 to -299 | 273 | 1.58 | 6.88e^−04^ | 3.44e^−04^ | 0.004 | 0.497 |
| Central region | SASA | R456W | -37 | -223 to 123 | 273 | 0.14 | 0.838 | 0.708 | 0.250 | 0.497 |
| Central region | SASA | D488E | -28 | -203 to 214 | 273 | 0.10 | 0.311 | 0.220 | 0.204 | 0.760 |
| Central region | SASA | T548A | +52 | -133 to 183 | 273 | 0.19 | 0.555 | 0.823 | 0.250 | 0.497 |
| Central region | SASA | S609P | +53 | -144 to 195 | 273 | 0.19 | 0.555 | 0.823 | 0.250 | 0.497 |
| Central region | SASA | I664L | -275 | -461 to -150 | 273 | 1.01 | 0.004 | 3.44e^−04^ | 0.004 | 0.497 |
| Central region | SASA | R684Q | -290 | -511 to -176 | 273 | 1.06 | 0.017 | 0.004 | 0.004 | 0.497 |
| RH domain | RMSD | R813W | -0.13 | -0.44 to 0.16 | 0.46 | 0.28 | 0.927 | 1.000 | 0.111 | 9.24e^−06^ |
| RH domain | RMSD | M882I | -0.19 | -0.45 to 0.16 | 0.46 | 0.41 | 0.927 | 1.000 | 0.250 | 0.178 |
| RH domain | RMSF | R813W | -0.12 | -0.38 to 0.11 | 0.32 | 0.38 | 0.275 | 0.351 | 0.250 | 0.515 |
| RH domain | RMSF | M882I | -0.21 | -0.46 to 0.02 | 0.32 | 0.66 | 0.254 | 0.351 | 0.250 | 0.856 |
| RH domain | R_g_ | R813W | -0.79 | -1.25 to -0.32 | 0.77 | 1.03 | 0.005 | 0.005 | 0.004 | 0.288 |
| RH domain | R_g_ | M882I | +0.00 | -0.57 to 0.31 | 0.77 | 0.00 | 0.533 | 0.647 | 0.250 | 0.613 |
| RH domain | SASA | R813W | +50 | -22 to 108 | 319 | 0.16 | 0.211 | 0.086 | 0.095 | 0.361 |
| RH domain | SASA | M882I | +124 | 50 to 248 | 319 | 0.39 | 0.006 | 0.024 | 0.003 | 0.474 |

^a^Regional rows list only the variants located within the interval concerned. ^b^Variant minus WT, in Å for RMSD, RMSF and R_g_ and Å^2^ for SASA. ^c^Moving-block bootstrap 95% CI of the mean difference. ^d^The denominator is the pointwise WT between-replica SD of Tables S4 to S7. ^e^BH-adjusted *q*-values, reproduced from Table S9. Full definitions are given in Methods.

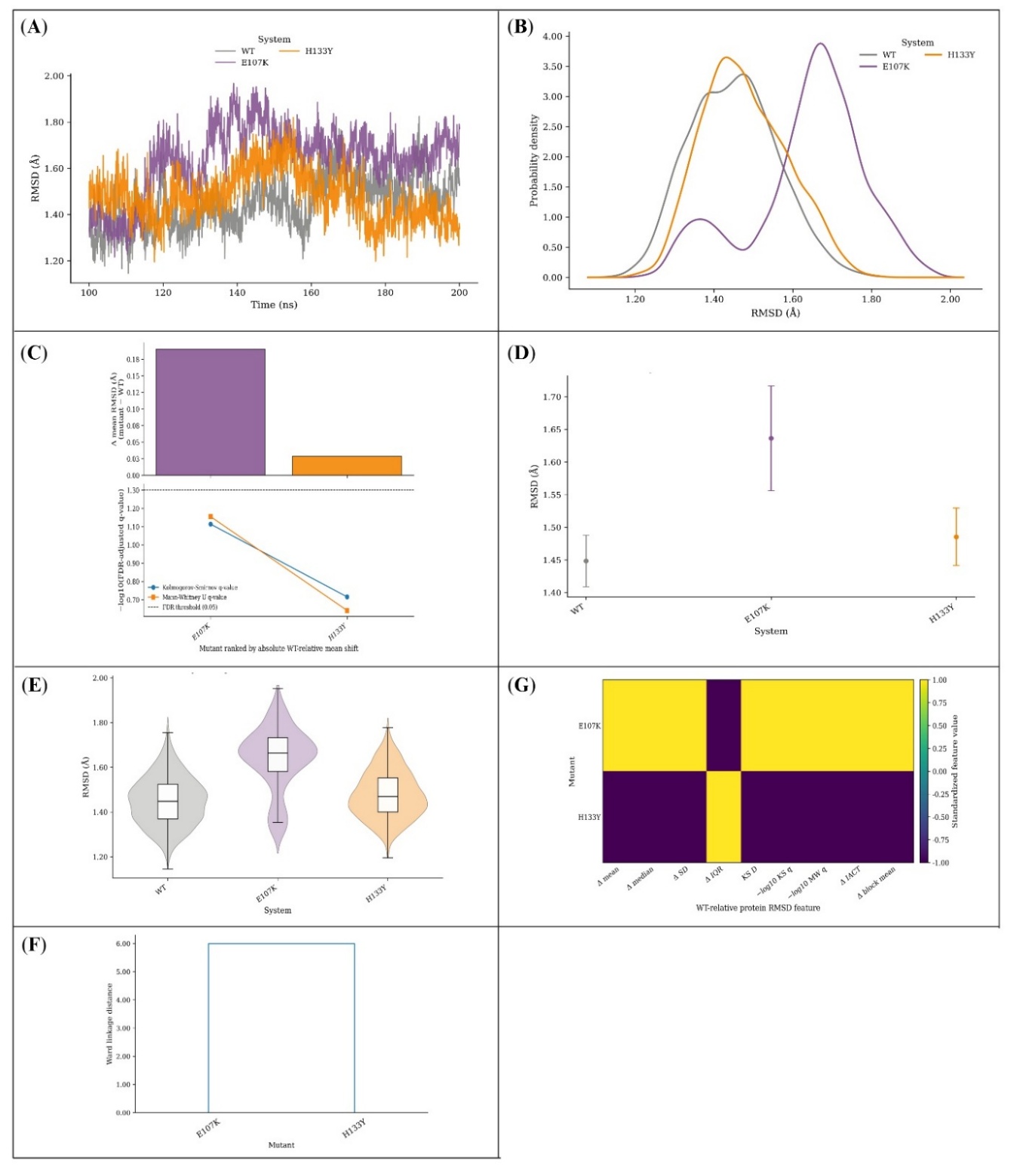

**Figure S13. Backbone RMSD of the RUN domain.** (A) Replica-averaged time series and (B) the corresponding probability densities. (C) WT-relative change in mean RMSD (upper) above the BH-adjusted KS and MWU *q*-values plotted as $-{log}_{10}q$ (lower), where the dashed line marks *q* = 0.05. (D) Block-averaged means with Student’s t-based 95% CIs. (E) Violin plots with embedded boxplots. (F) Ward-linkage dendrogram and (G) heatmap of standardised WT-relative features. This domain contains only two variants, so panel F reduces to a single pairwise split and the standardised values in panel G take only the two extreme levels. Profiles were aligned on the backbone atoms of the domain itself, so displacement is measured within the domain rather than against the whole protein. The *q*-values are exploratory descriptors of the replica-averaged profiles.

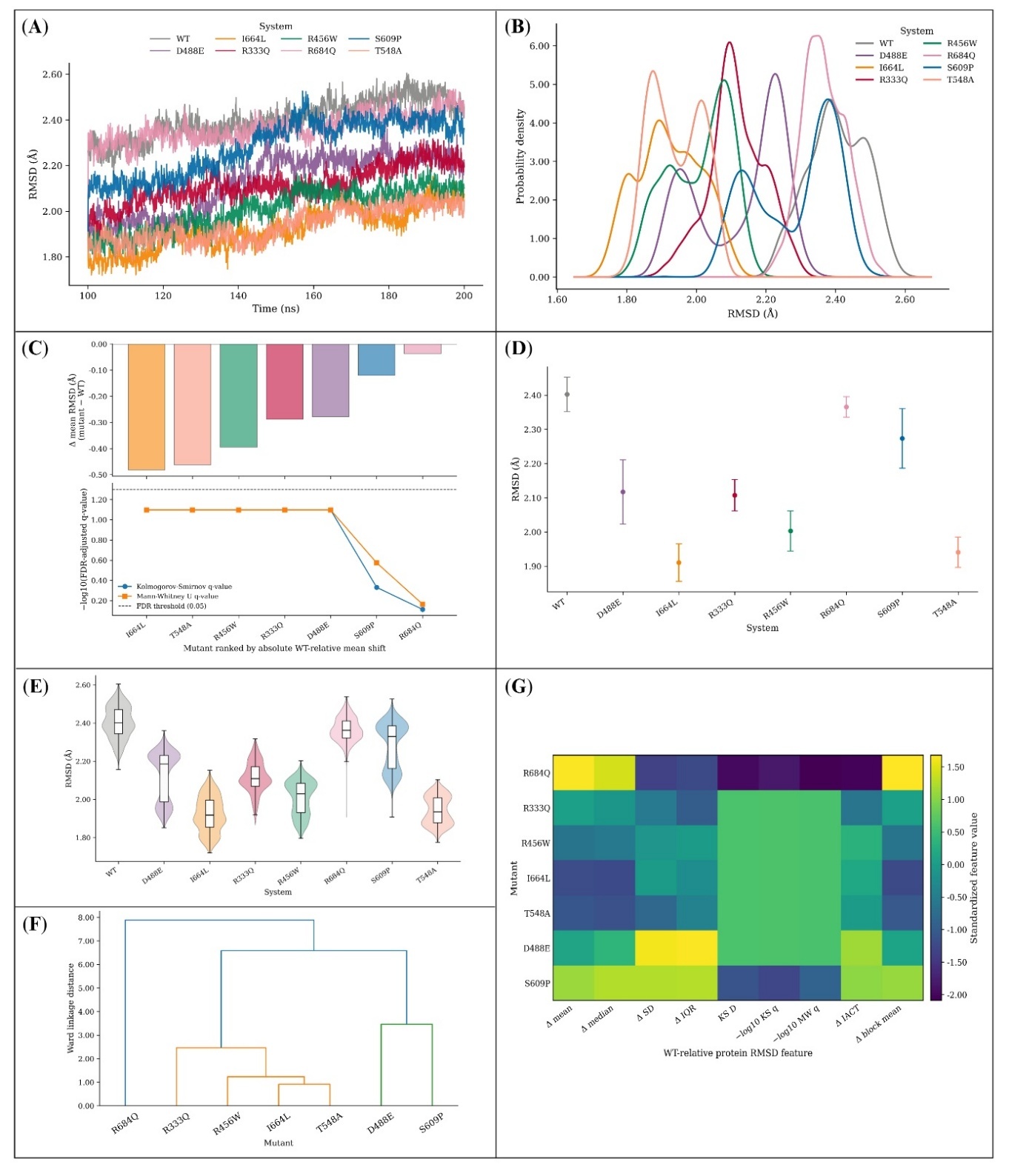

**Figure S14. Backbone RMSD of the central region.** Panels A and B give the replica-averaged time series and its probability density. Panel C shows the WT-relative change in mean RMSD, with BH-adjusted KS and MWU *q*-values below it as $-{log}_{10}q$ and a dashed line at *q* = 0.05. Block-averaged means with Student’s t-based 95% CIs follow in D and violin plots with embedded boxplots in E. Panels F and G cluster the standardised WT-relative features as a Ward-linkage dendrogram and display them as a heatmap. As elsewhere in this series, the *q*-values describe separation between averaged profiles and carry no trajectory-level inference.

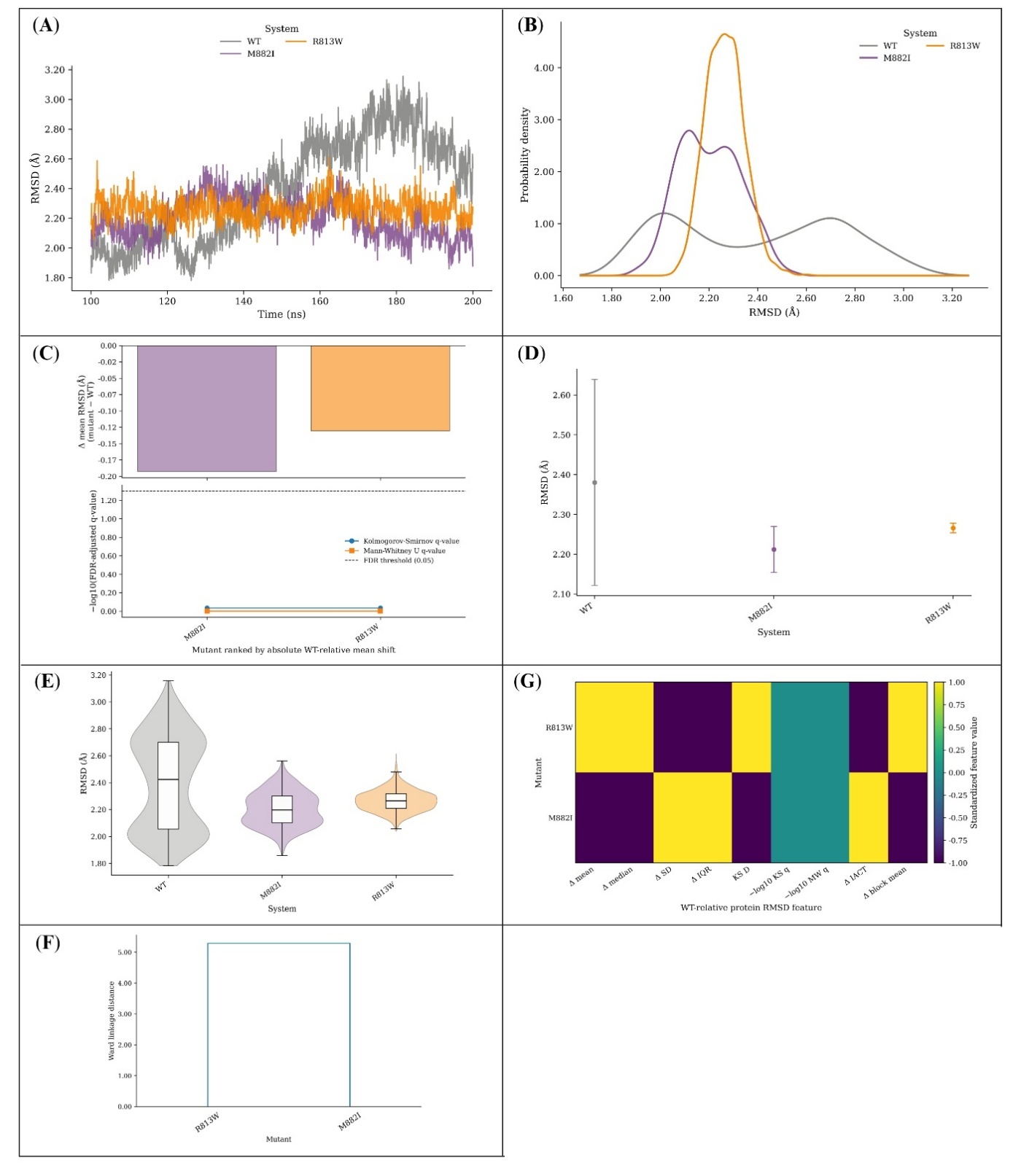

**Figure S15. Backbone RMSD of the RH domain.** Seven panels: the replica-averaged time series (A) and its probability density (B); the WT-relative change in mean RMSD with BH-adjusted KS and MWU *q*-values as $-{log}_{10}q$ against a dashed line at *q* = 0.05 (C); block-averaged means with Student’s t-based 95% CIs (D); violin plots with embedded boxplots (E); and a Ward-linkage dendrogram (F) with its matching heatmap (G) of standardised WT-relative features. Only two variants map to this domain, so panel F shows a pairwise separation rather than a cluster structure. The *q*-values remain exploratory profile-level descriptors.

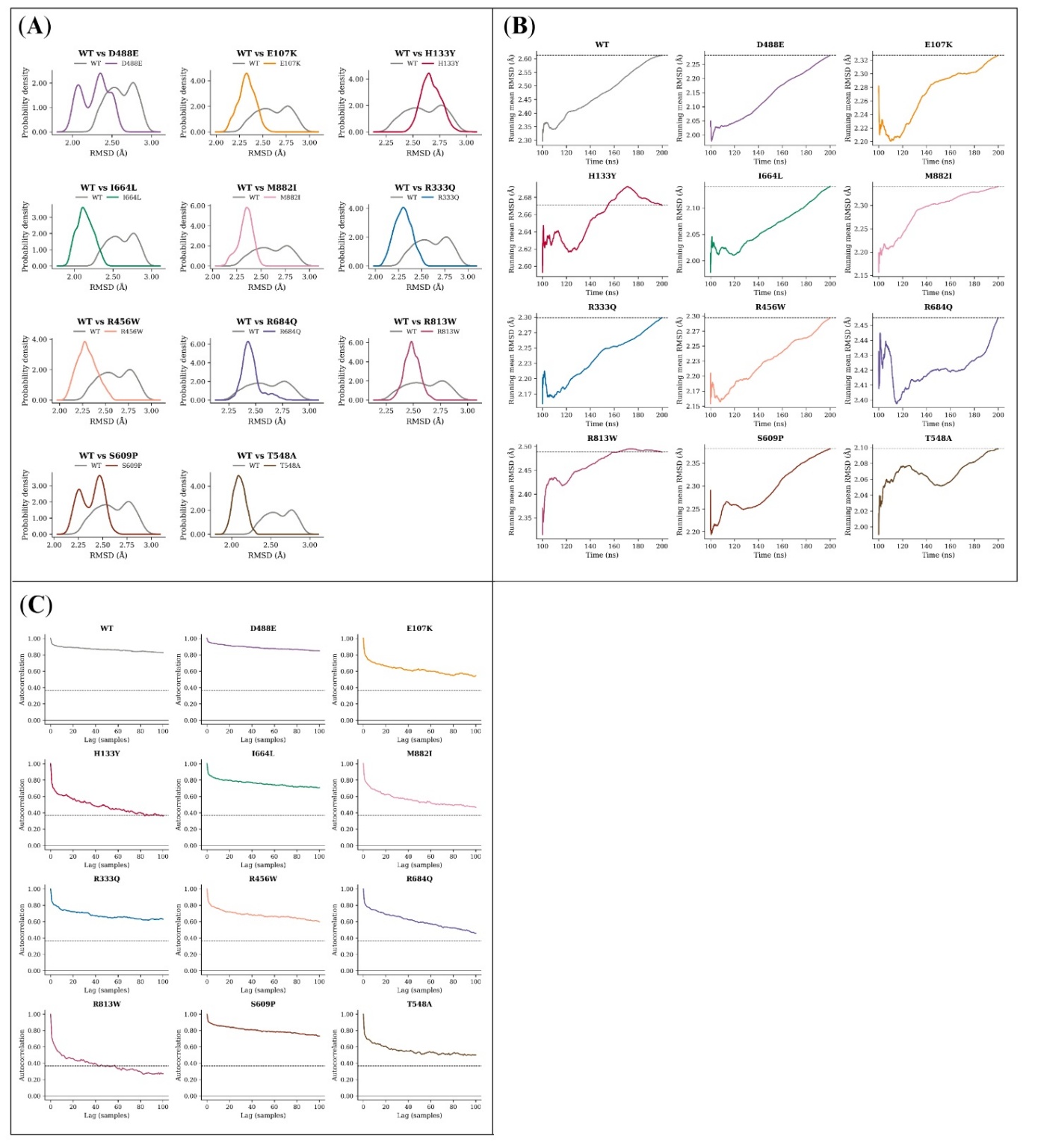

**Figure S16. Distributional and temporal diagnostics of full-length RUBICON RMSD.**
(A) WT paired with each variant as overlaid probability densities. (B) Profile mean accumulated frame by frame; the dashed line marks the final mean. (C) Autocorrelation against temporal lag; the dashed line marks 1/*e*. For display, the autocorrelation curves are truncated at 100 samples; the statistical inefficiency used for RMSD subsampling was estimated over the wider prespecified lag range extending to *N*/2. These diagnostics describe the replica-averaged profiles and should be read alongside the independent-replica summaries in Figures S45 and S46.

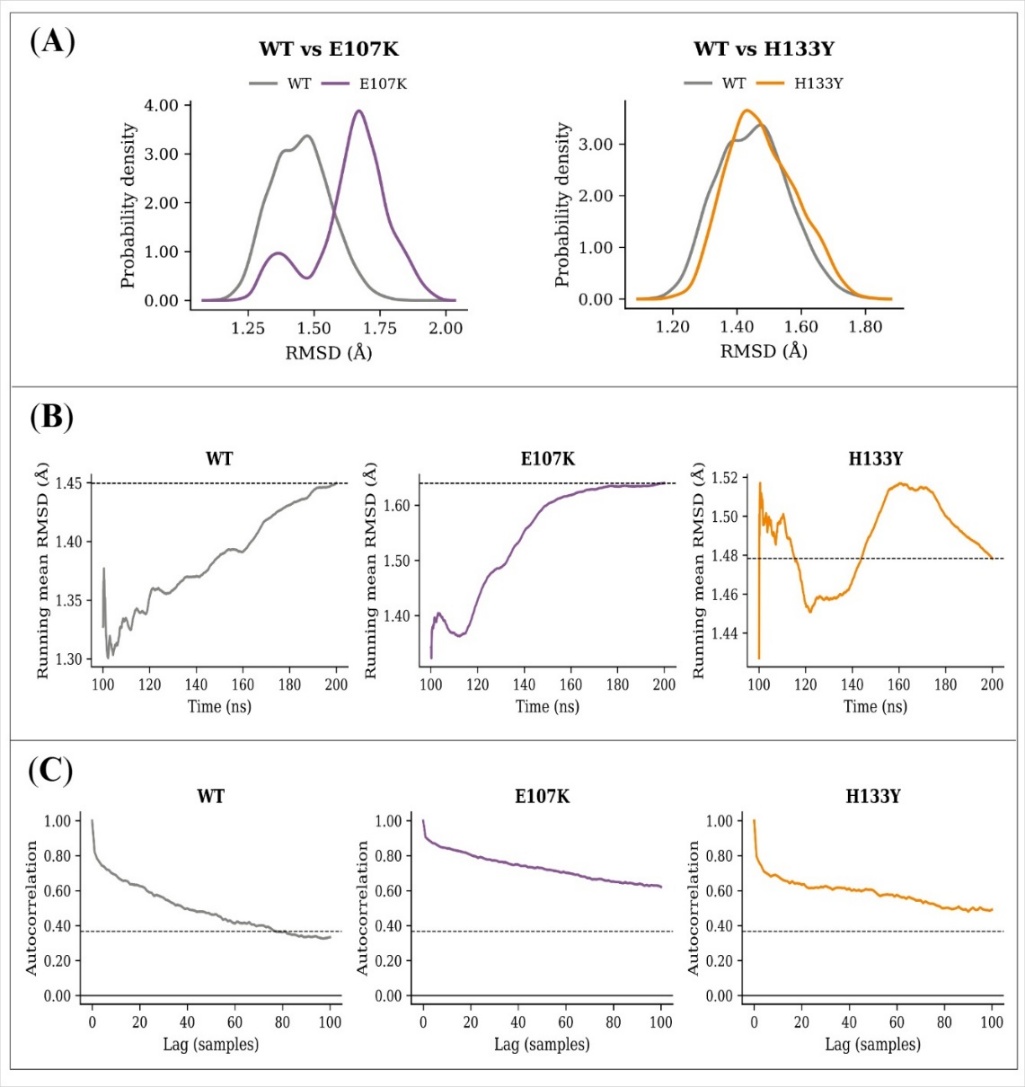

**Figure S17. Distributional and temporal diagnostics of RUN-domain RMSD.** (A) WT overlaid against each variant as probability densities. (B) Running mean of each profile, with a dashed line at its final value. (C) Autocorrelation against temporal lag, with a dashed line at 1/*e*. The autocorrelation is drawn to a lag of 100 samples for display; the statistical inefficiency used for subsampling was estimated over the wider prespecified range.

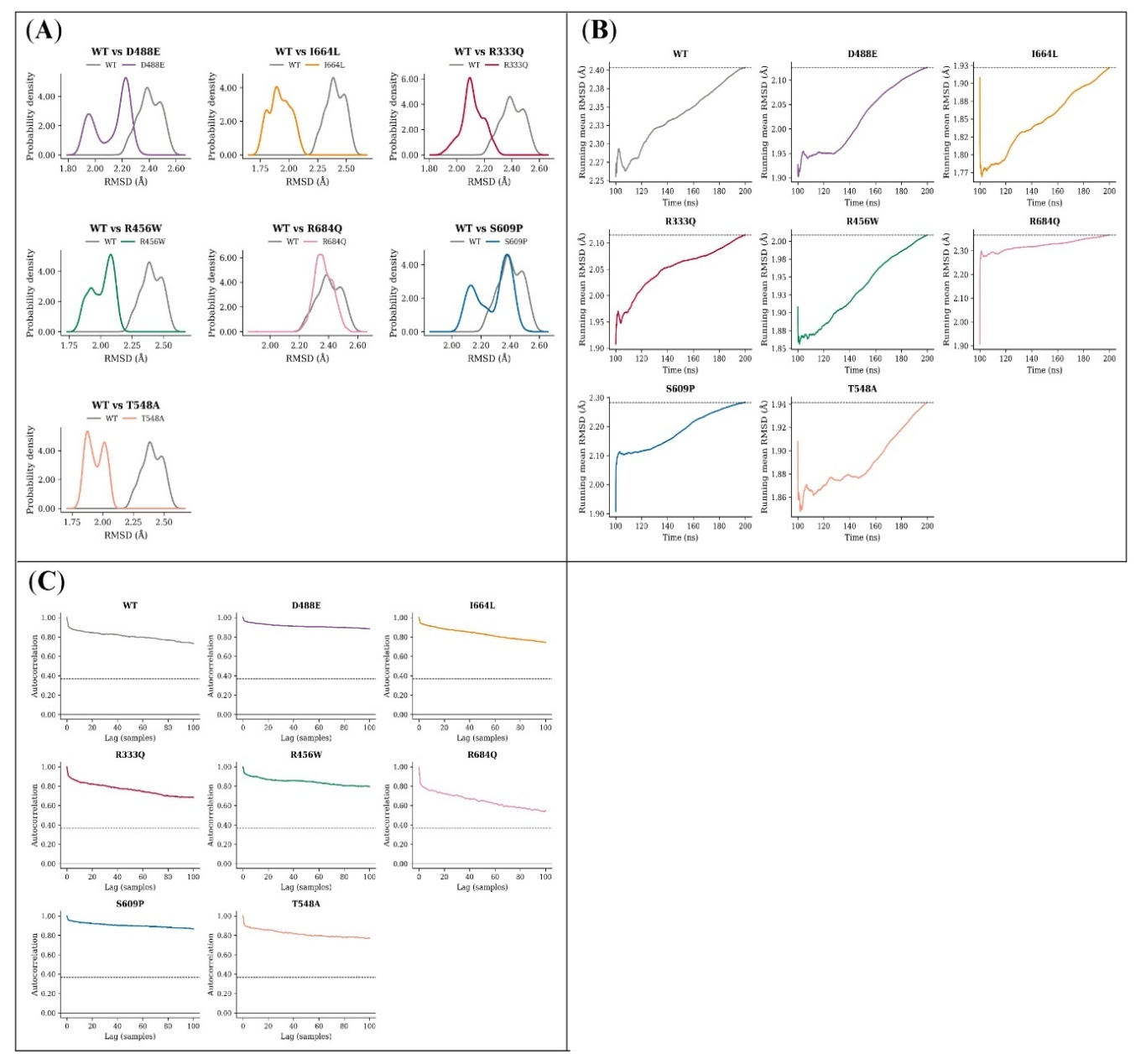

**Figure S18. Distributional and temporal diagnostics of central-region RMSD.** Panel A overlays the WT density on that of each variant in turn. Panel B tracks the profile mean as it accumulates frame by frame, the dashed line marking the final value. Panel C plots autocorrelation against temporal lag, with 1/*e* indicated. The lag axis is truncated at 100 samples for display only.

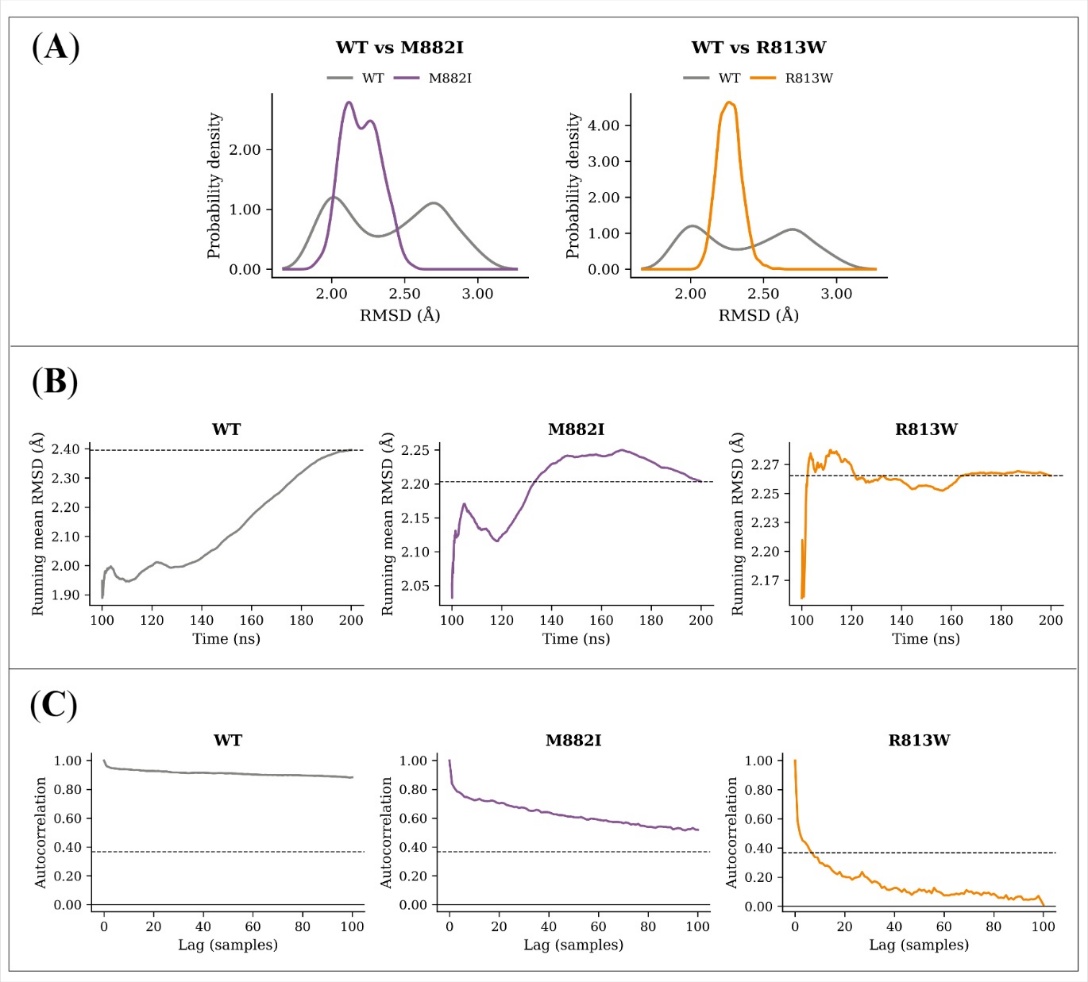

**Figure S19. Distributional and temporal diagnostics of RH domain RMSD.** Three panels: WT-against-variant densities (A), running means with the final value dashed (B), and autocorrelation against temporal lag with the 1/*e* level dashed (C). The displayed lag range is again truncated at 100 samples.

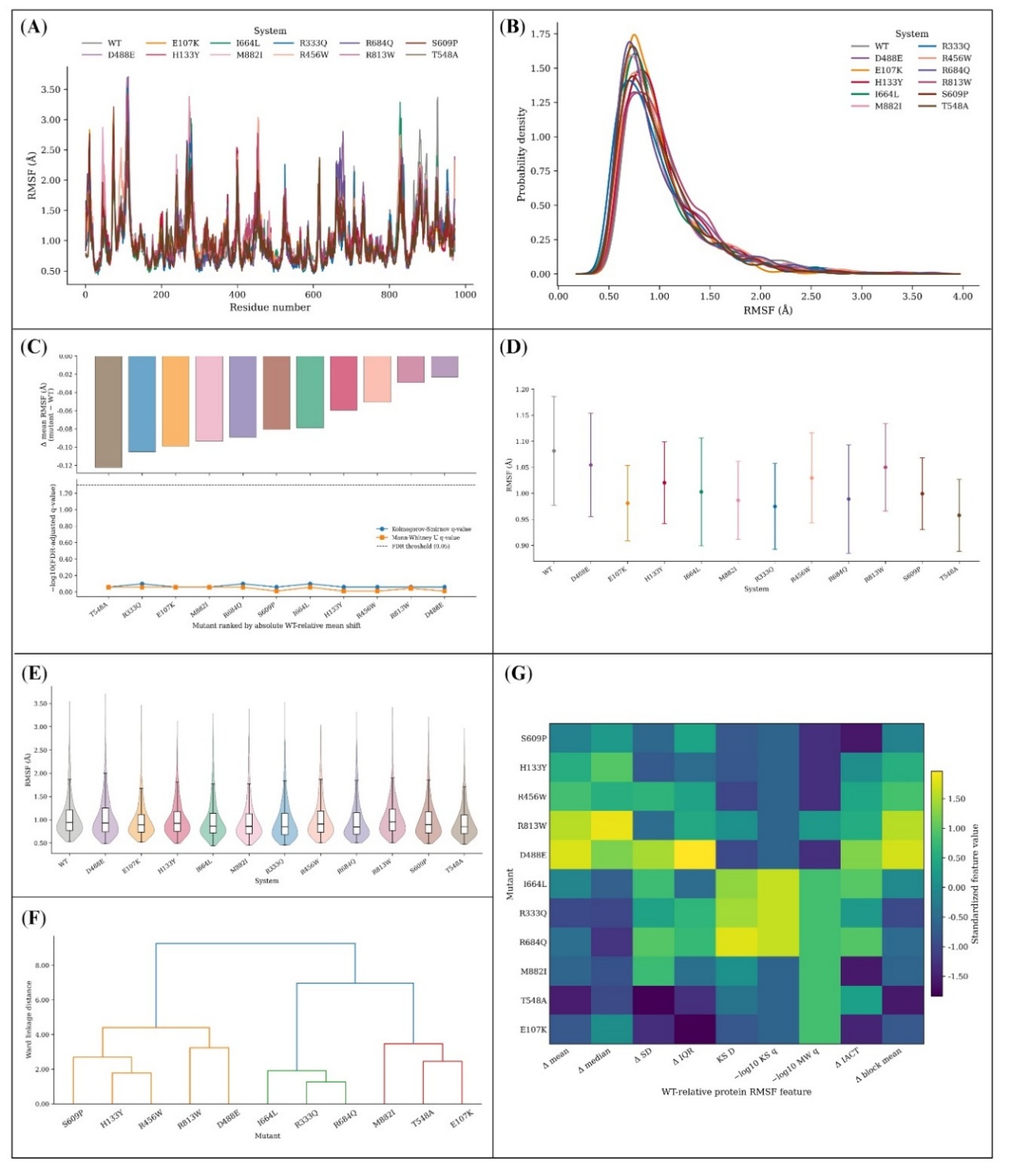

**Figure S20. Cα RMSF across the full length of RUBICON.** (A) Replica-averaged profile against residue number and (B) its probability density. (C) WT-relative change in mean fluctuation (upper) above the BH-adjusted KS and MWU *q*-values plotted as $-{log}_{10}q$ (lower), with a dashed line at *q* = 0.05. (D) Sequence-block means with Student’s t-based 95% CIs. (E) Violin plots with embedded boxplots. (F) Ward-linkage dendrogram and (G) heatmap of standardised WT-relative features. Neighbouring residues fluctuate together, so the *q*-values measure separation between averaged profiles rather than testing independent observations. The unpaired tests also compare marginal RMSF distributions and ignore the natural WT-variant pairing at each residue.

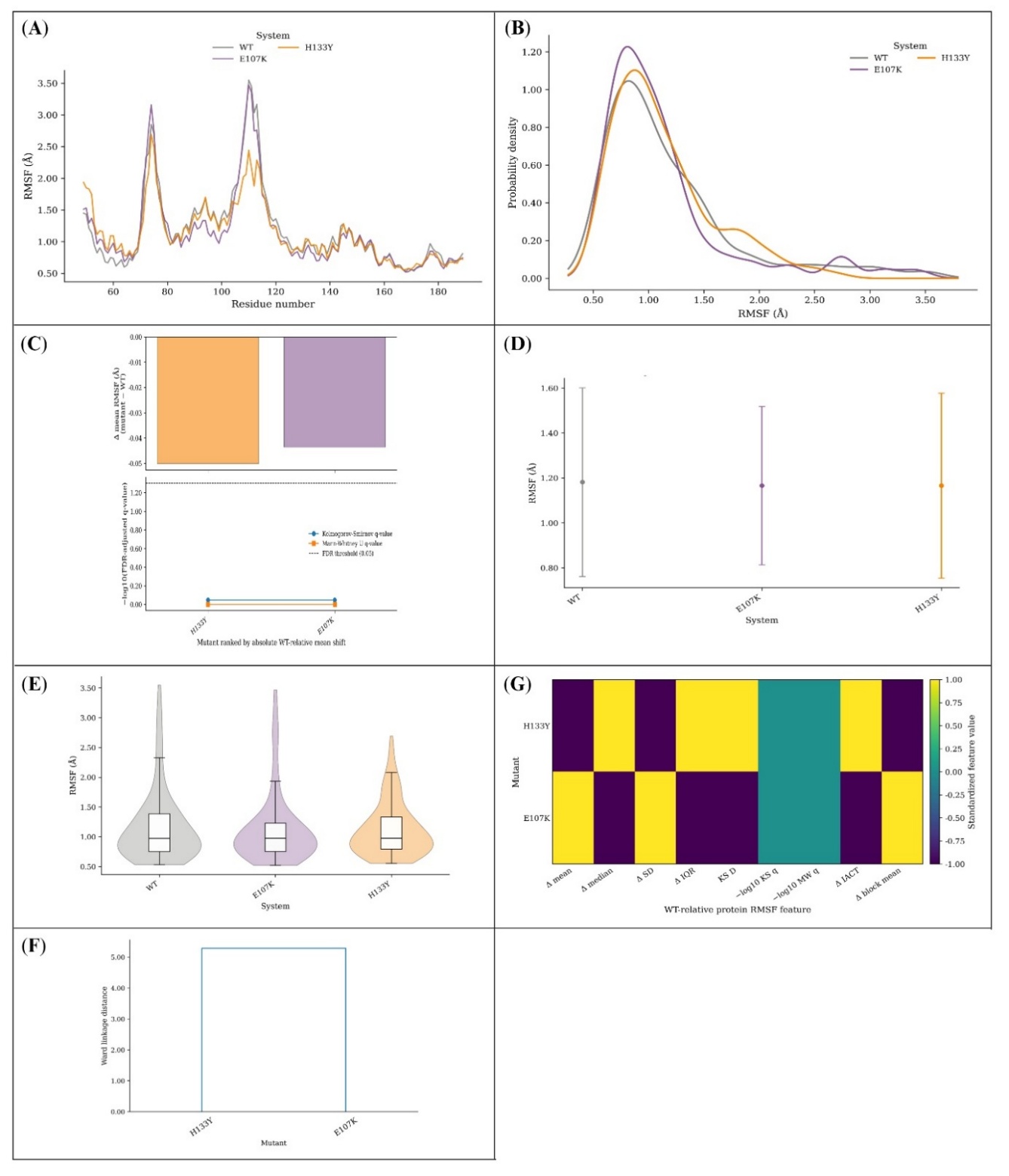

**Figure S21. Cα RMSF across the RUN domain.** Panel A traces the replica-averaged profile against residue number, panel B its probability density. Panel C places the WT-relative change in mean fluctuation above the BH-adjusted KS and MWU *q*-values as $-{log}_{10}q$, the dashed line marking *q* = 0.05. Sequence-block means with Student’s t-based 95% CIs follow in D and violin plots with embedded boxplots in E, while F and G render the standardised WT-relative features as a Ward-linkage dendrogram and a heatmap. Only two variants map to this domain, so panel F reduces to a single split and panel G to two extreme levels.

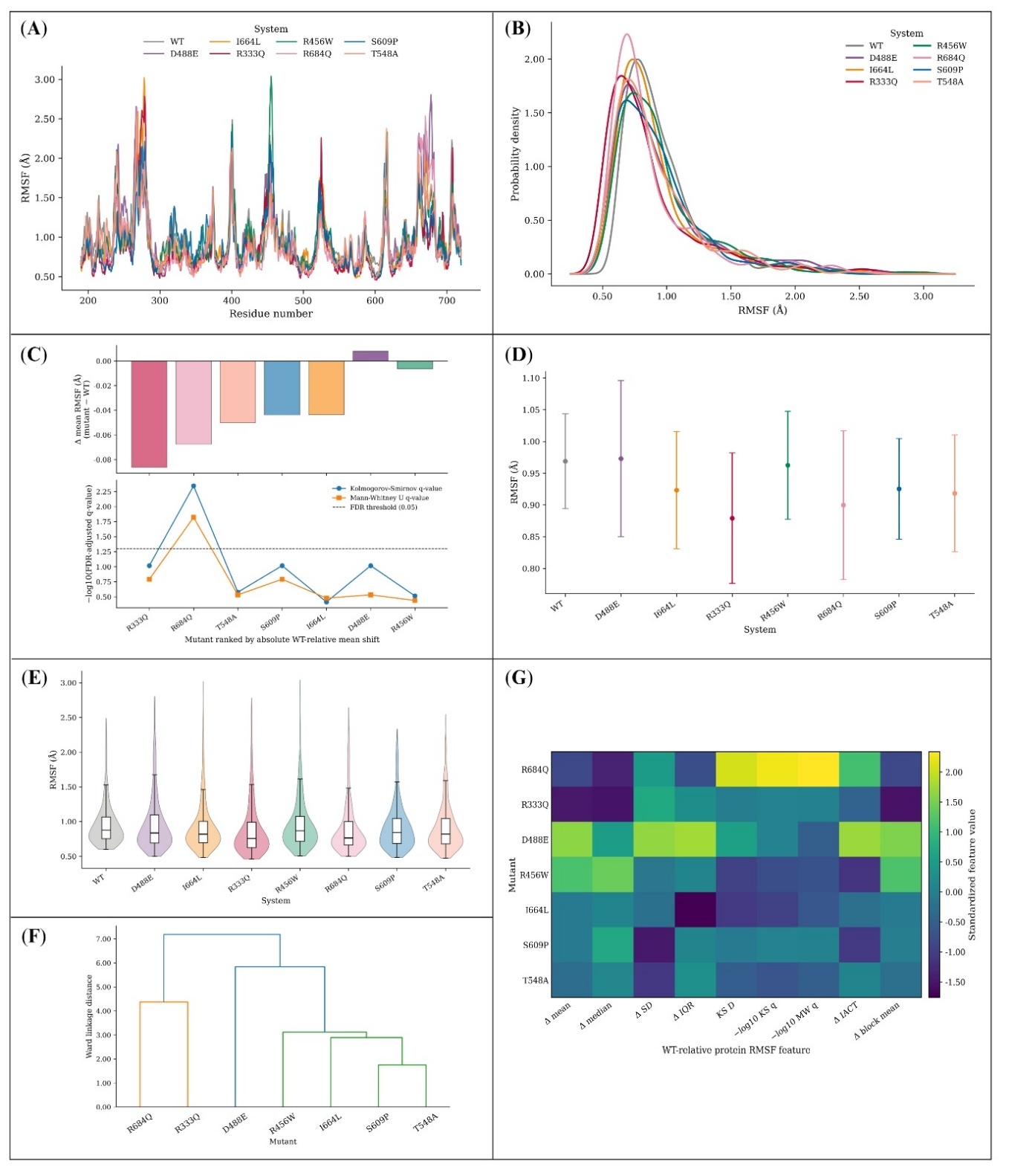

**Figure S22. Cα RMSF across the central region.** The replica-averaged profile against residue number appears in A and its probability density in B. Panel C gives the WT-relative change in mean fluctuation with the BH-adjusted KS and MWU *q*-values below it as $-{log}_{10}q$ and a dashed line at *q* = 0.05. The remaining panels show sequence-block means with Student’s t-based 95% CIs (D), violin plots with embedded boxplots (E), and a Ward-linkage dendrogram (F) with matching heatmap (G) of standardised WT-relative features. The *q*-values are exploratory and do not preserve same-residue pairing.

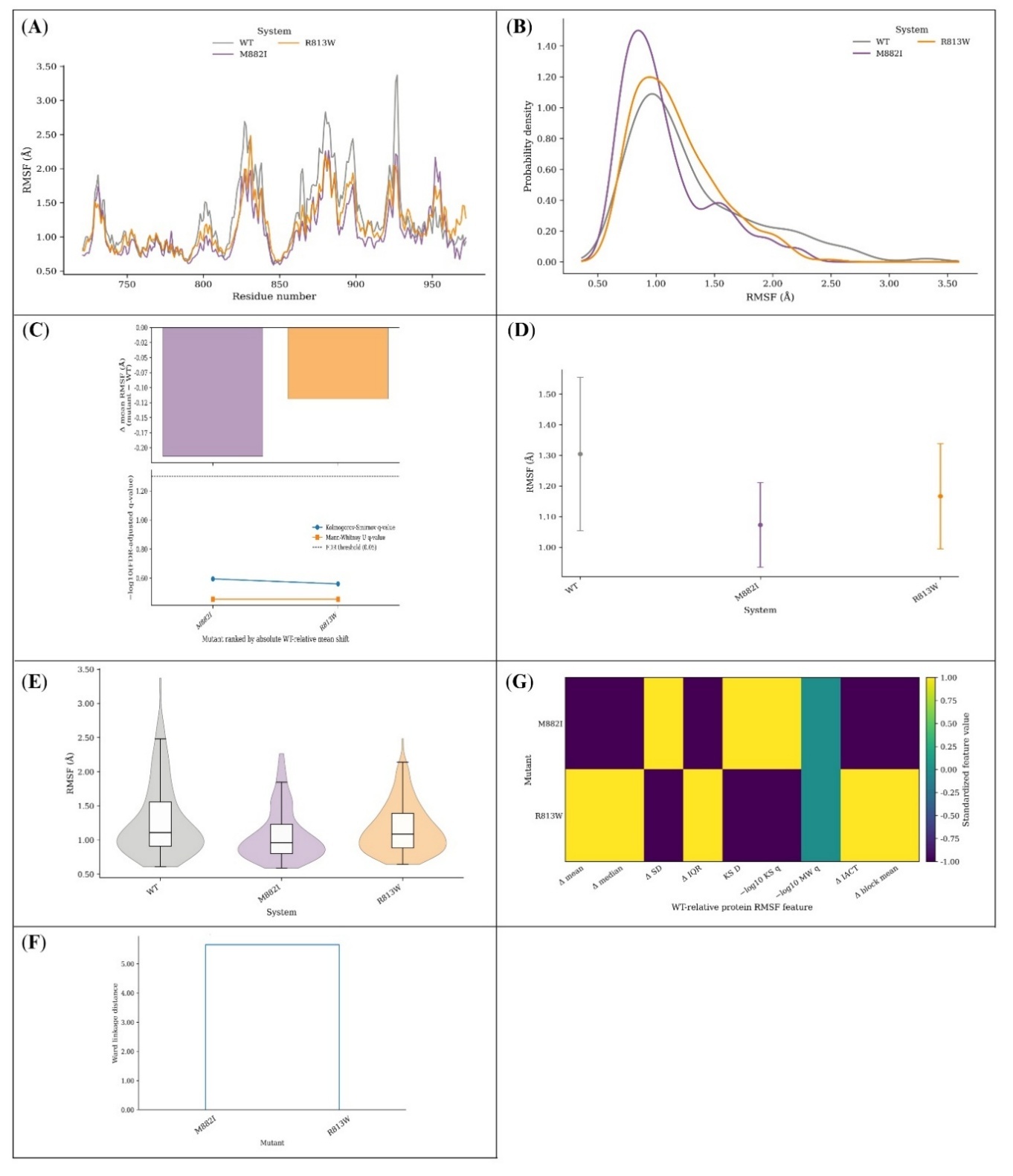

**Figure S23. Cα RMSF across the RH domain.** Panels A and B present the replica-averaged profile against residue number and its probability density. Panel C shows the WT-relative change in mean fluctuation over the BH-adjusted KS and MWU q-values as $-{log}_{10}q$, with *q* = 0.05 dashed. Sequence-block means with Student’s t-based 95% CIs are in D, violin plots with embedded boxplots in E, and the Ward-linkage dendrogram and heatmap of standardised WT-relative features in F and G. This domain also contains only two variants, limiting F to a pairwise separation.

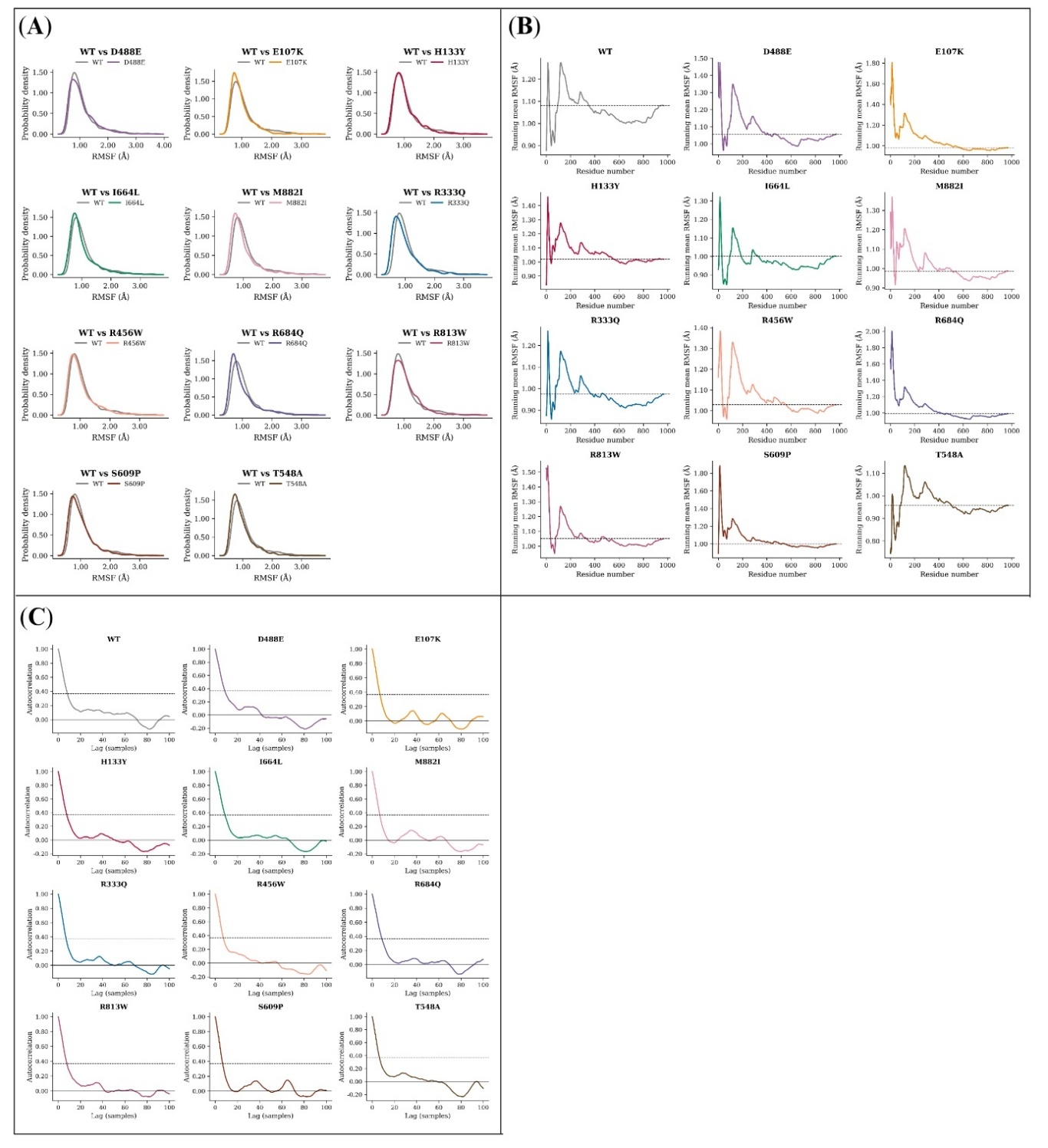

**Figure S24. Sequence diagnostics of full-length Cα RMSF.** (A) WT density overlaid on that of each variant. (B) Running mean along the sequence, dashed at its final value. (C) Autocorrelation against sequence lag, dashed at 1/*e*. One lag unit is one residue, so panel C reports how far fluctuation values remain correlated along the chain rather than in time.

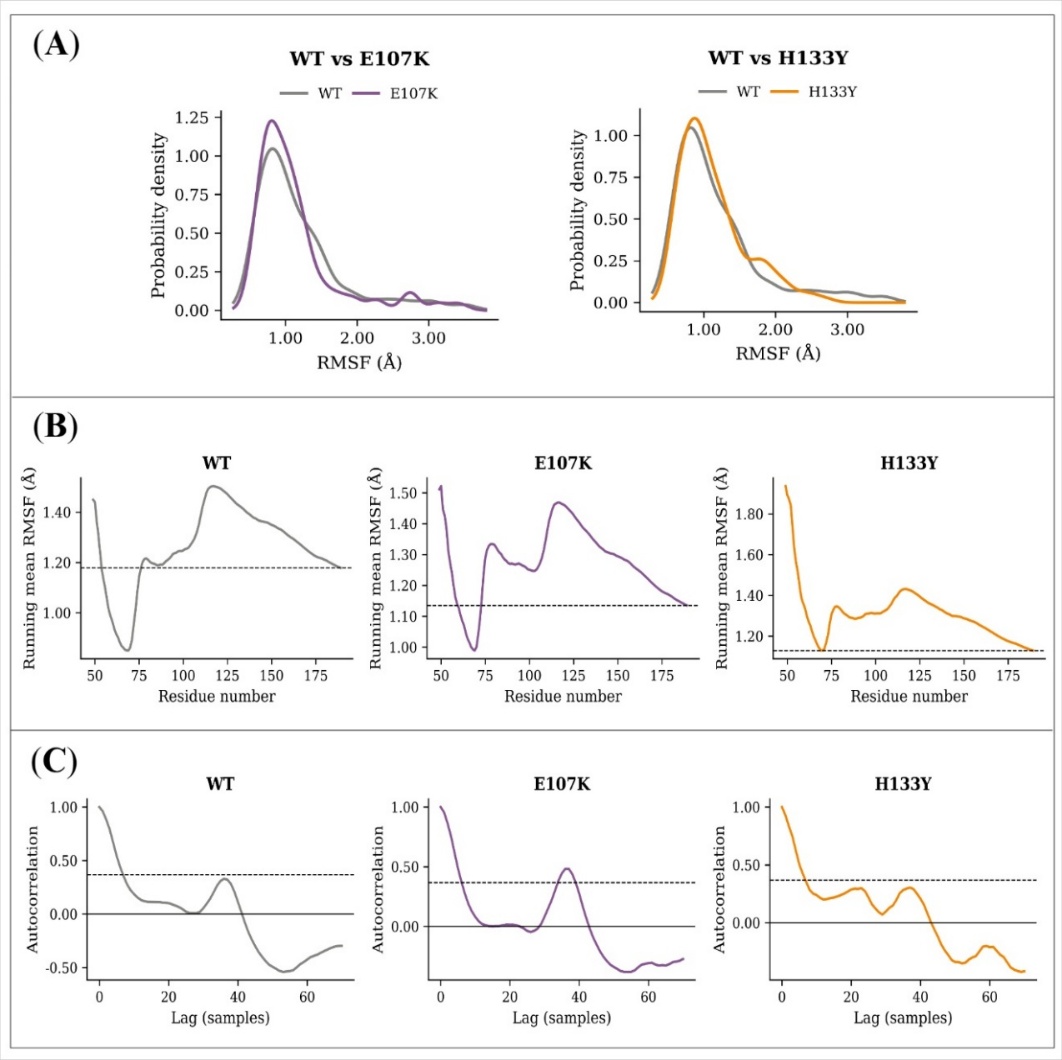

**Figure S25. Sequence diagnostics of RUN-domain Cα RMSF.** Panel A overlays the WT density on each variant in turn, panel B follows the running mean along residues 49 to 189 with a dashed line at its final value, and panel C plots autocorrelation against sequence lag with 1/*e* dashed. The lag axis reaches only 70 residues, half the domain length, and is therefore shorter than in the other figures of this series.

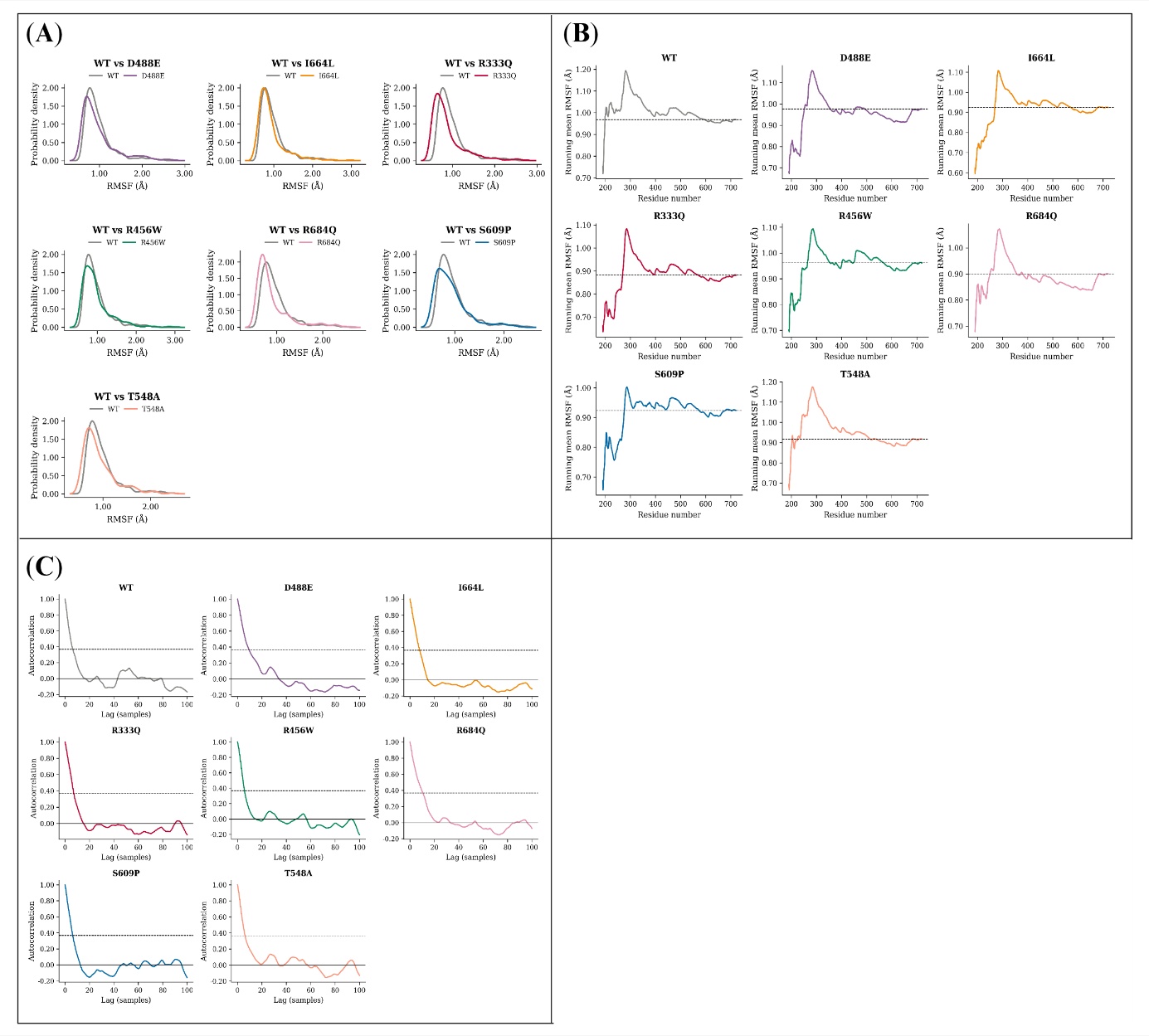

**Figure S26. Sequence diagnostics of central-region Cα RMSF.** (A) Pairwise WT-against-variant probability densities. (B) Running mean accumulated along the sequence, dashed at its final value. (C) Autocorrelation against sequence lag, dashed at 1/*e*.

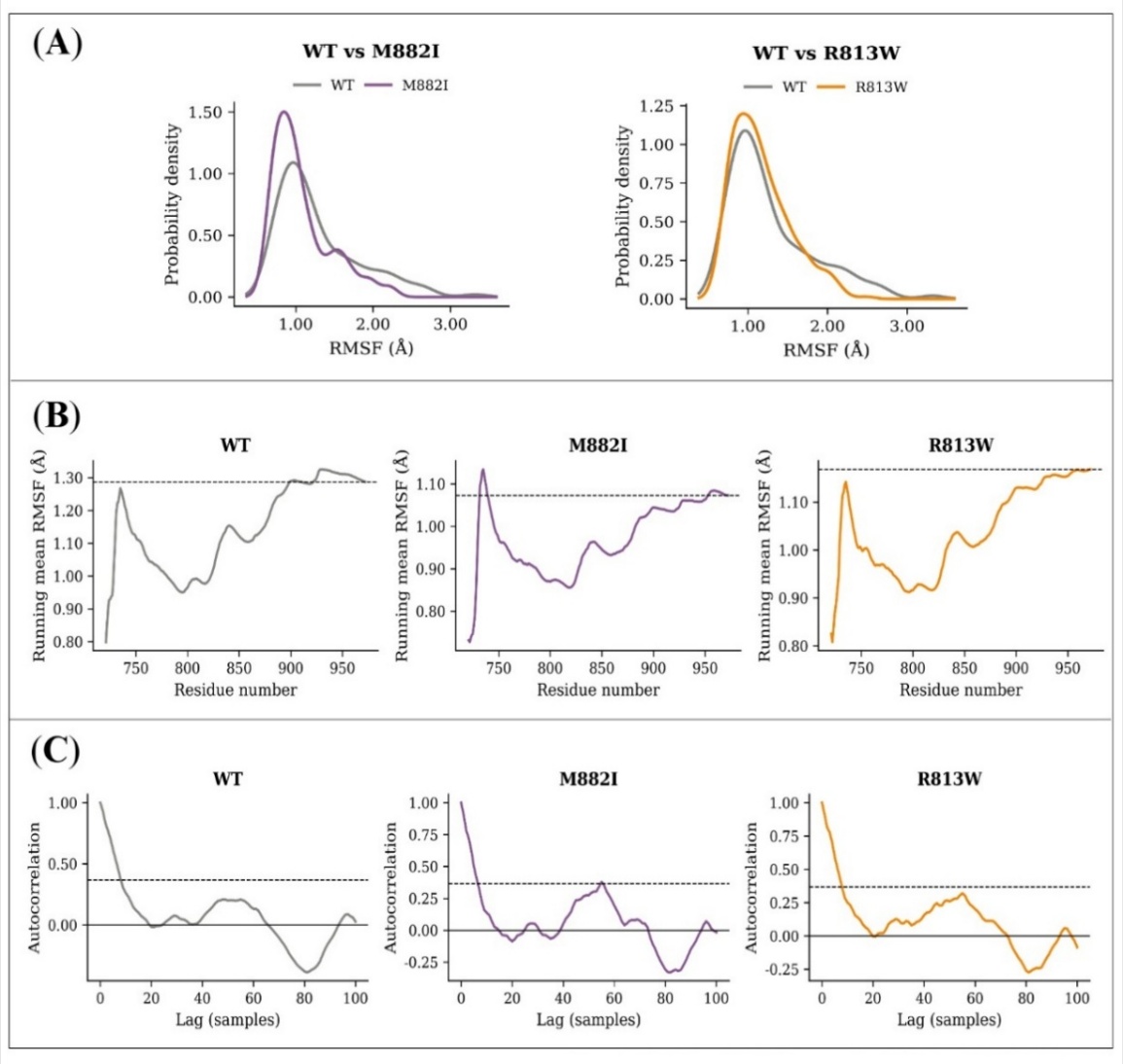

**Figure S27. Sequence diagnostics of RH domain Cα RMSF.** (A) Pairwise WT-against-variant probability densities. (B) Running means with the final value dashed. (C) Autocorrelation against sequence lag with the 1/*e* level dashed.

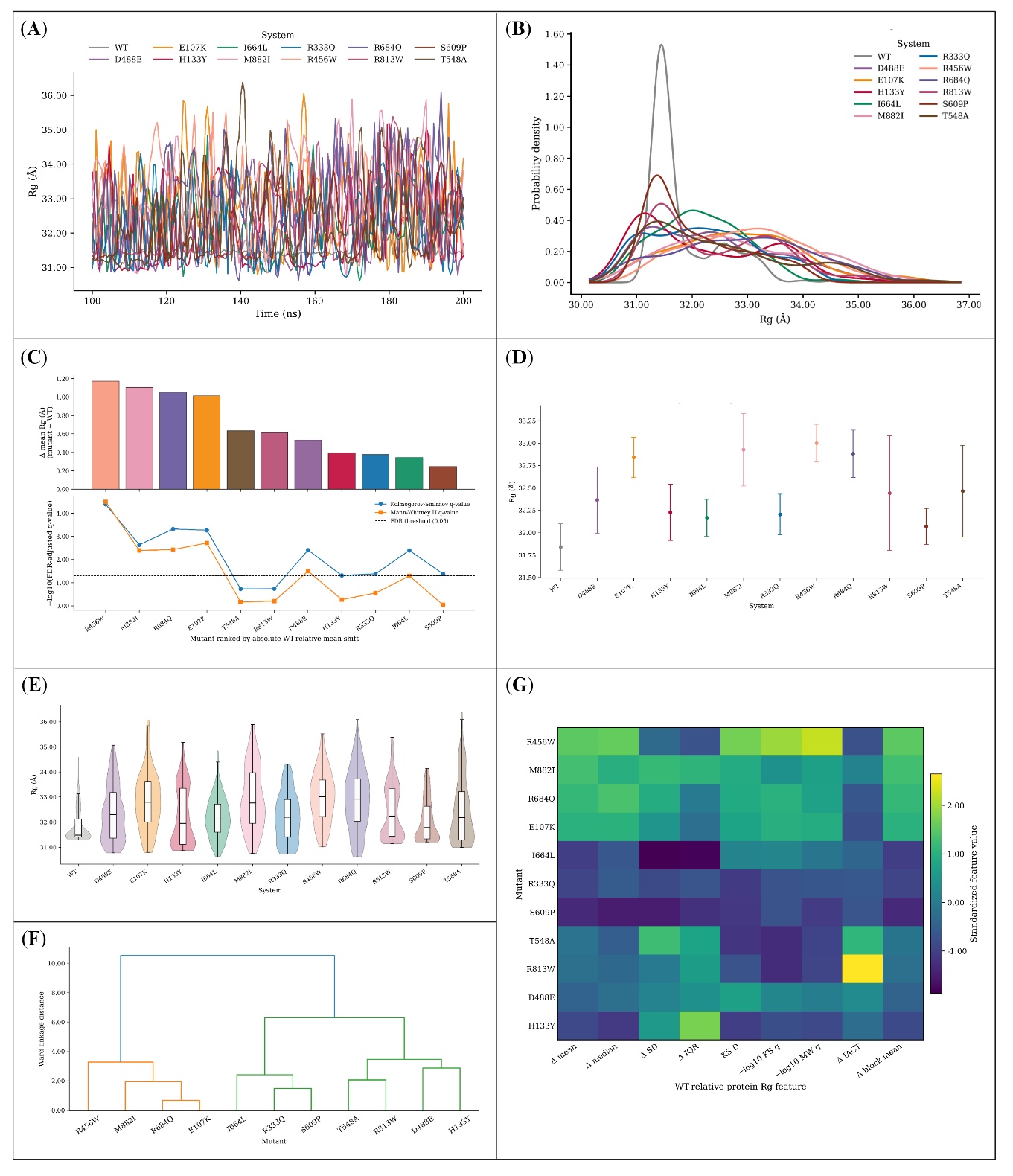

**Figure S28. R_g_ of full-length RUBICON.** (A) Replica-averaged time series and (B) its probability density. (C) WT-relative change in mean R_g_ (upper) above the BH-adjusted KS and MWU *q*-values plotted as $-{log}_{10}q$ (lower), with a dashed line at *q* = 0.05. (D) Block-averaged means with Student’s t-based 95% CIs. (E) Violin plots with embedded boxplots. (F) Ward-linkage dendrogram and (G) heatmap of standardised WT-relative features. Every variant shifts to a larger mean R_g_ than WT, and several cross the exploratory threshold in panel C. These *q*-values compare replica-averaged profiles and are not trajectory-level tests; Table S11 gives the corresponding replica-level summaries.

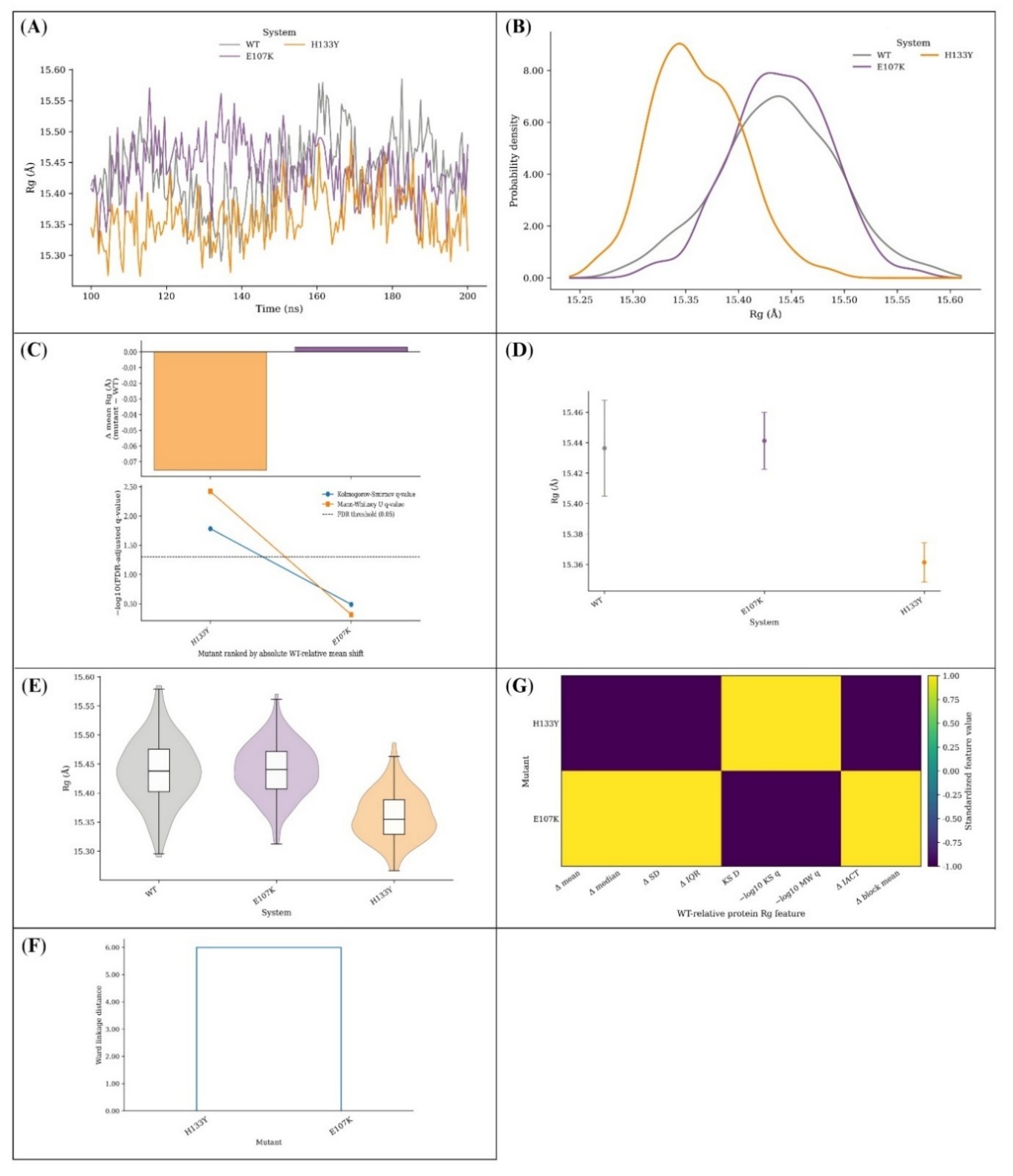

**Figure S29. R_g_ of the RUN domain.** (A) Replica-averaged time series, (B) probability densities, (C) WT-relative change in mean R_g_ over the BH-adjusted KS and MWU *q*-values as $-{log}_{10}q$ with *q* = 0.05 dashed, (D) block-averaged means with Student’s t-based 95% CIs, (E) violin plots with embedded boxplots, (F) Ward-linkage dendrogram and (G) heatmap of standardised WT-relative features. The domain carries only two variants, so panel F reduces to a single split and panel G to two extreme levels. The *q*-values remain exploratory profile-level descriptors.

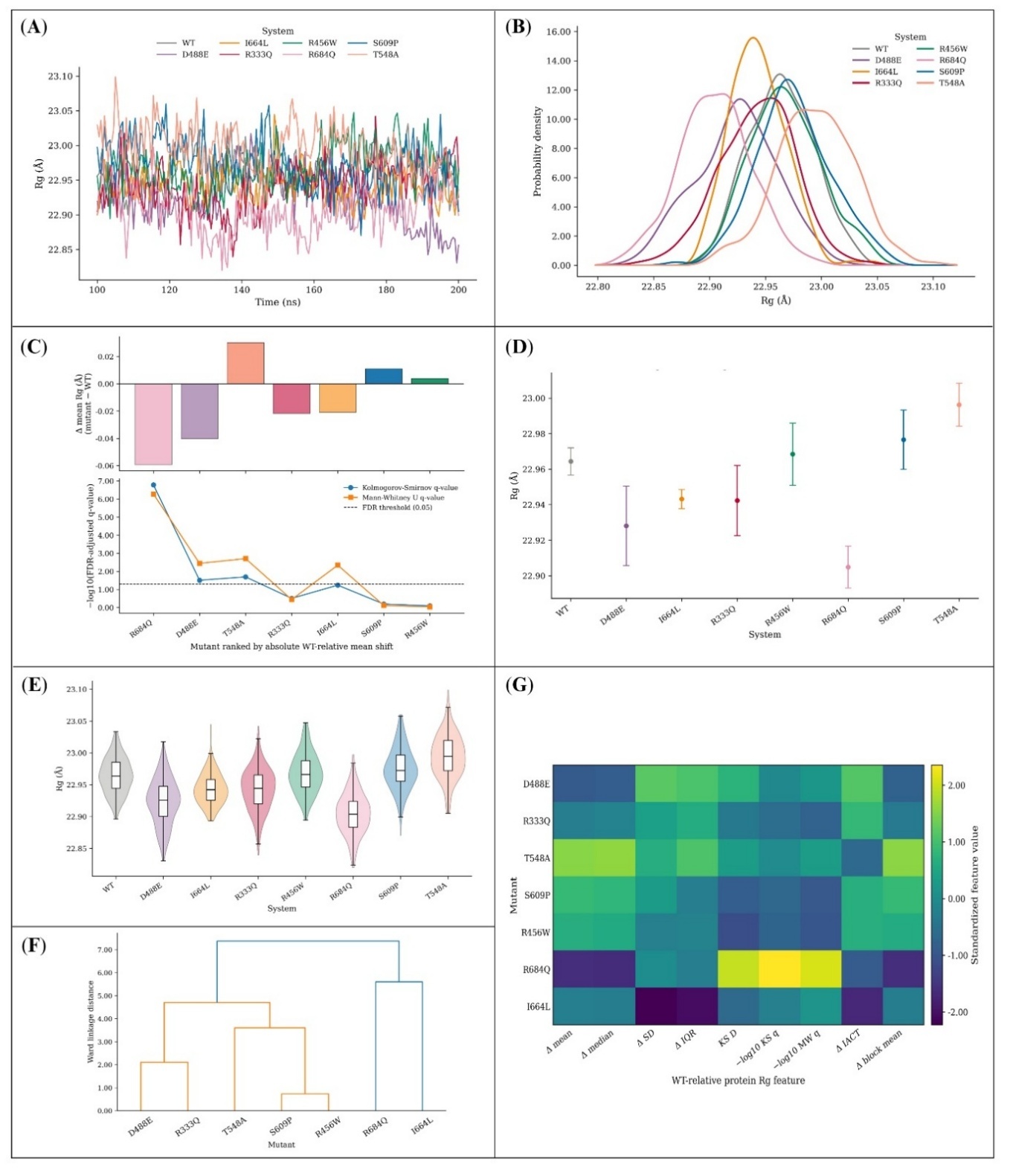

**Figure S30. R_g_ of the central region.** Panel A gives the replica-averaged time series and panel B its probability density. Panel C shows the WT-relative change in mean Rg with the BH-adjusted KS and MWU *q*-values below it as $-{log}_{10}q$ and a dashed line at *q* = 0.05. Block-averaged means with Student’s t-based 95% CIs follow in D, violin plots with embedded boxplots in E, and the Ward-linkage dendrogram with its matching heatmap of standardised WT-relative features in F and G. As throughout this series, the *q*-values describe separation between averaged profiles only.

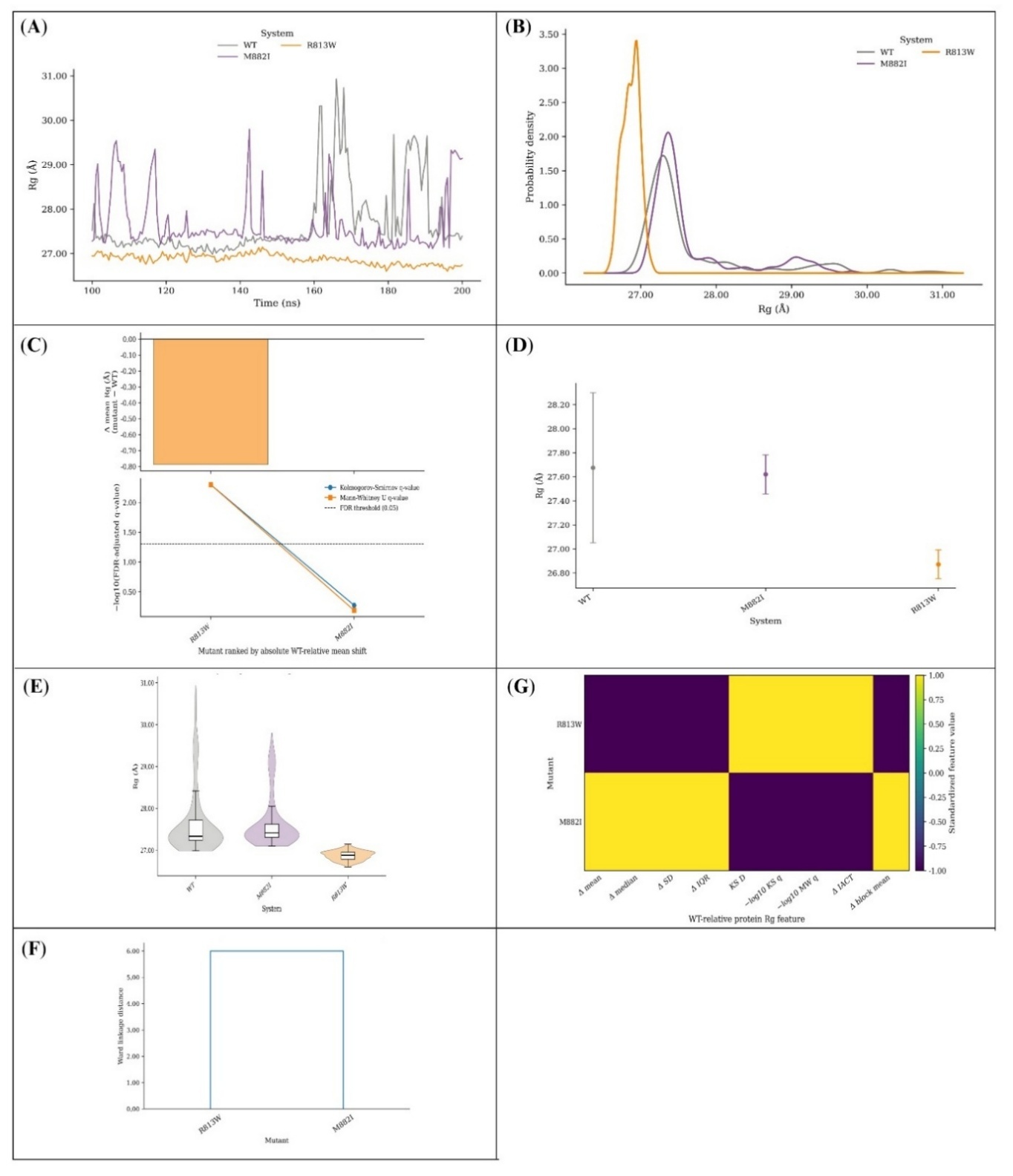

**Figure S31. R_g_ of the RH domain.** Seven panels: replica-averaged time series (A), probability density (B), WT-relative change in mean R_g_ with BH-adjusted KS and MWU *q*-values as $-{log}_{10}q$ against a dashed line at *q* = 0.05 (C), block-averaged means with Student’s t-based 95% CIs (D), violin plots with embedded boxplots (E), and a Ward-linkage dendrogram (F) with the corresponding heatmap (G) of standardised WT-relative features. Only two variants map here, limiting panel F to a pairwise separation. The *q*-values are exploratory.

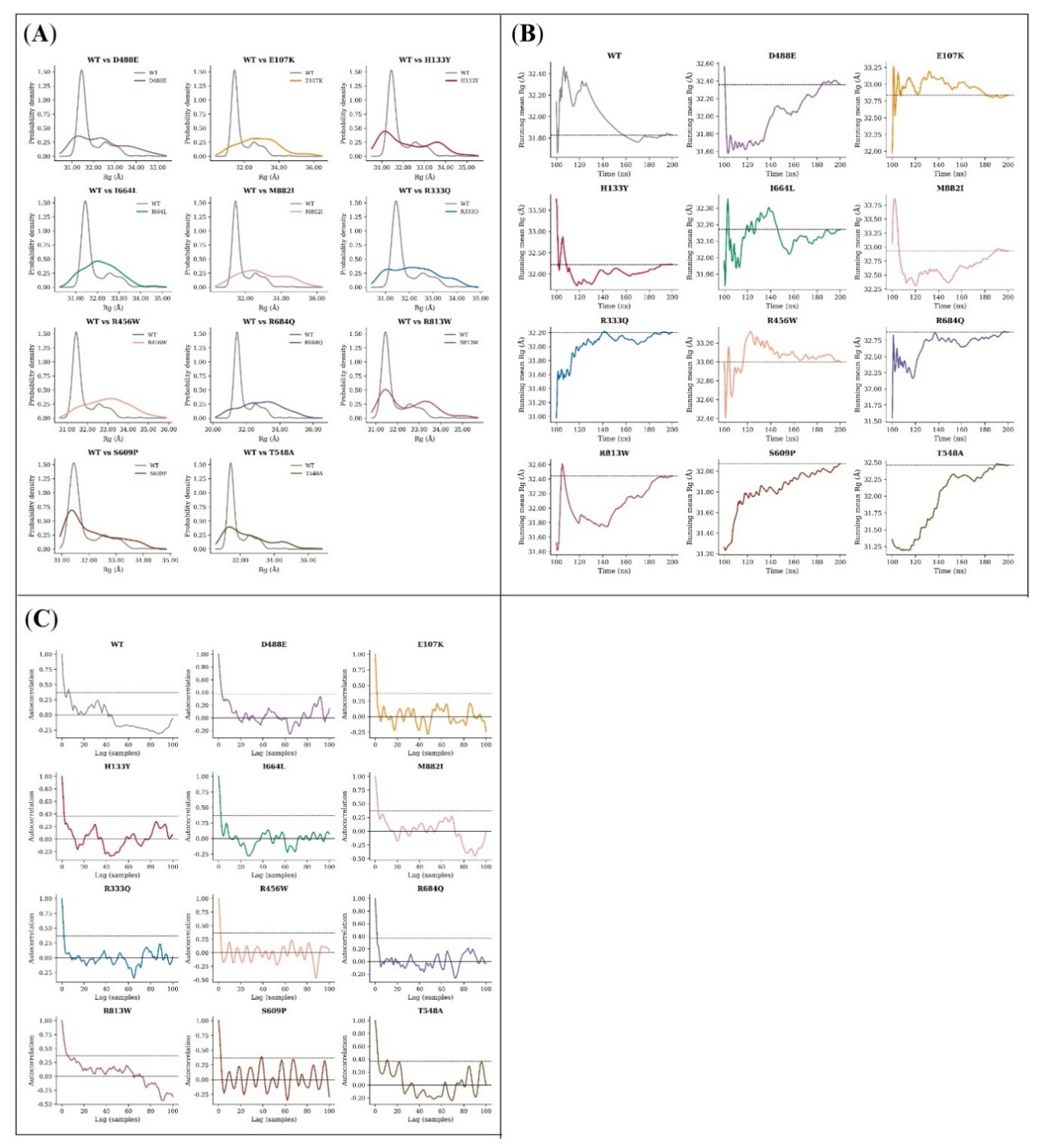

**Figure S32. Distributional and temporal diagnostics of full-length R_g_.** (A) WT density compared with each variant in turn. (B) Running mean accumulated frame by frame, dashed at its final value. (C) Autocorrelation against temporal lag, dashed at 1/*e*. R_g_ was stored every 500 ps, so one lag unit corresponds to 500 ps in all four figures of this series.

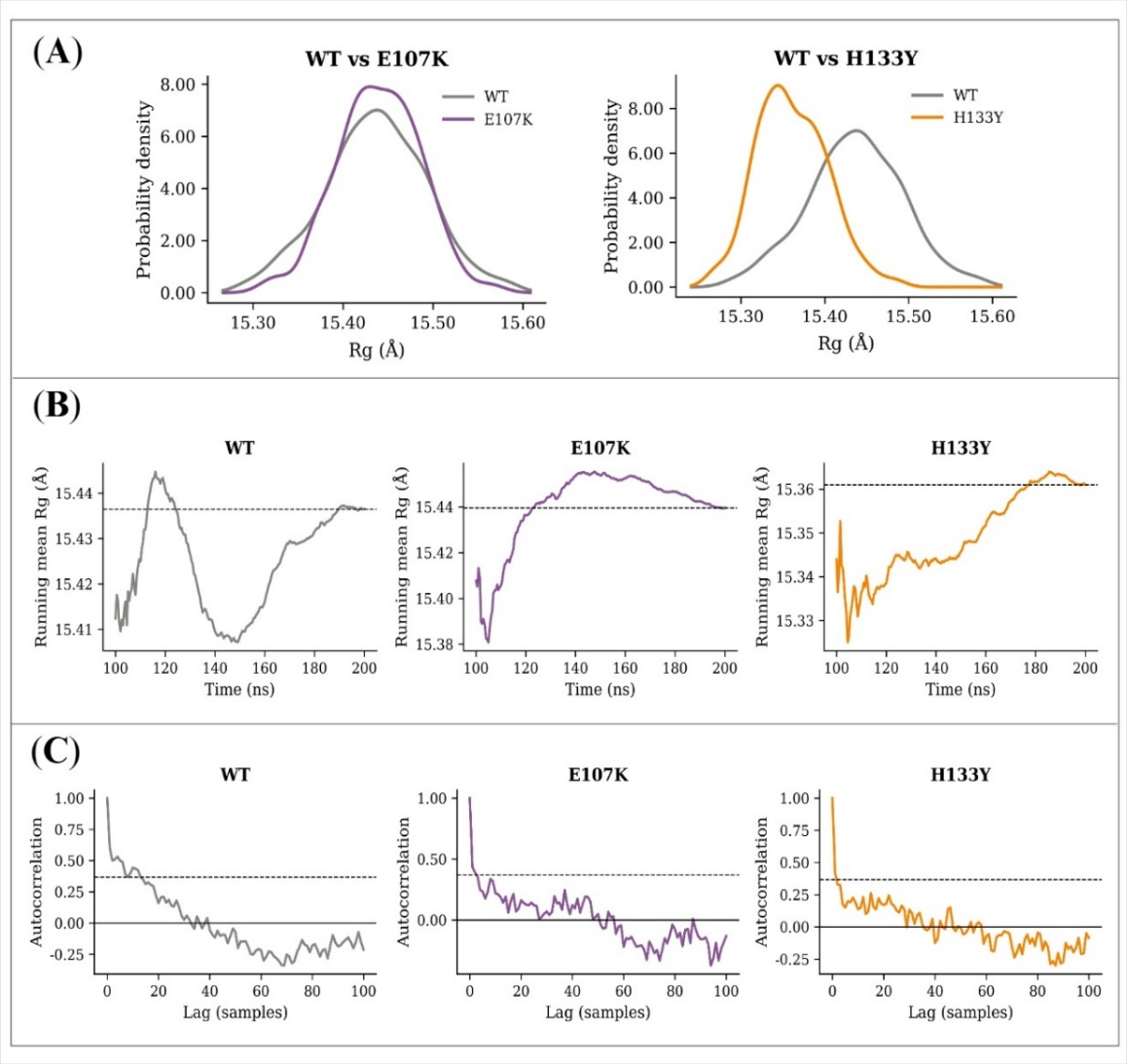

**Figure S33. Distributional and temporal diagnostics of RUN-domain R_g_.** (A) Pairwise WT-against-variant probability densities. (B) Running means, dashed at the final profile mean. (C) Autocorrelation against temporal lag, dashed at 1/*e*.

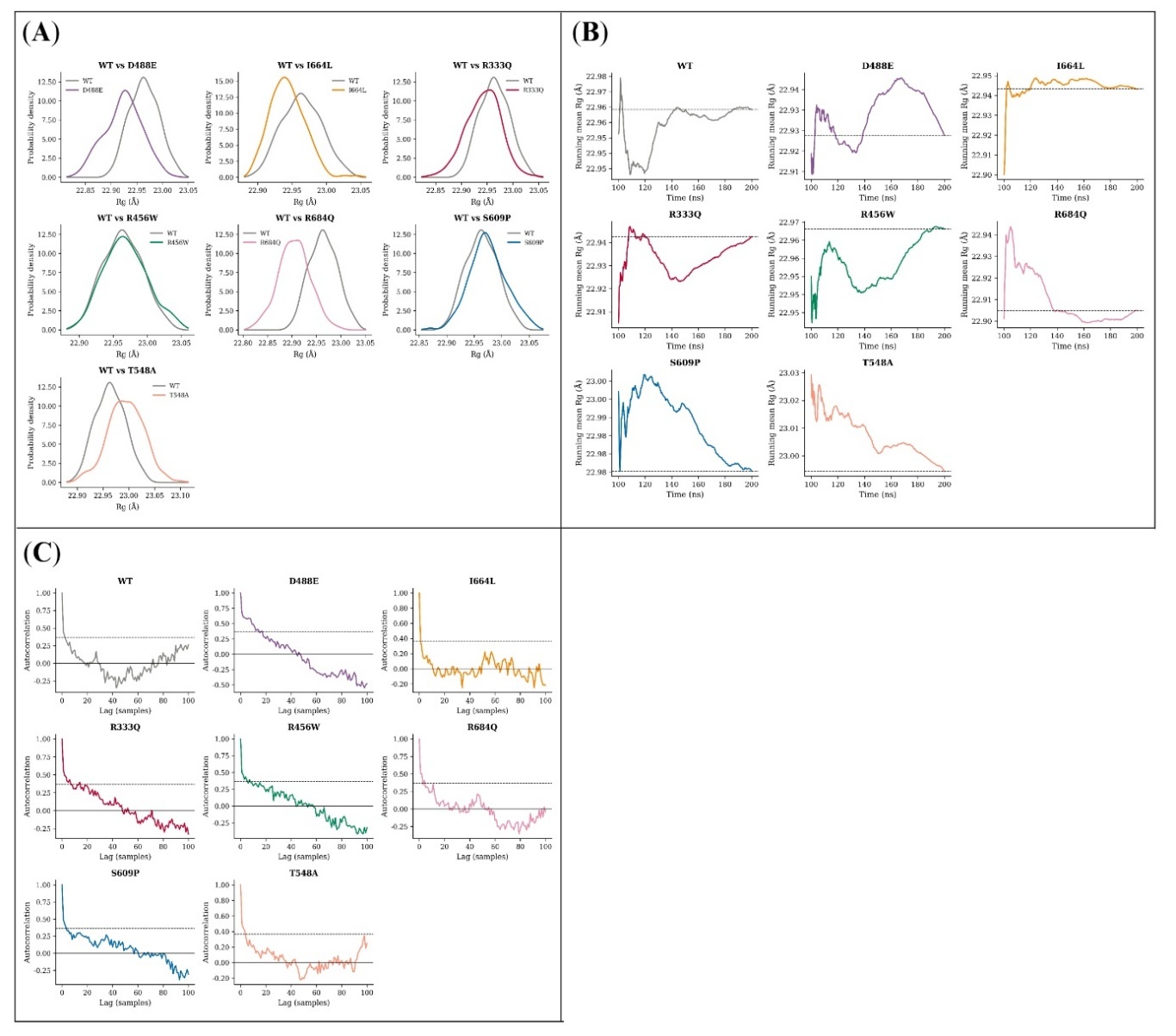

**Figure S34. Distributional and temporal diagnostics of central-region R_g_.** (A) WT overlaid on each variant as probability densities. (B) Profile mean accumulated frame by frame, with the final value dashed. (C) Autocorrelation of each profile against temporal lag, with 1/*e* dashed.

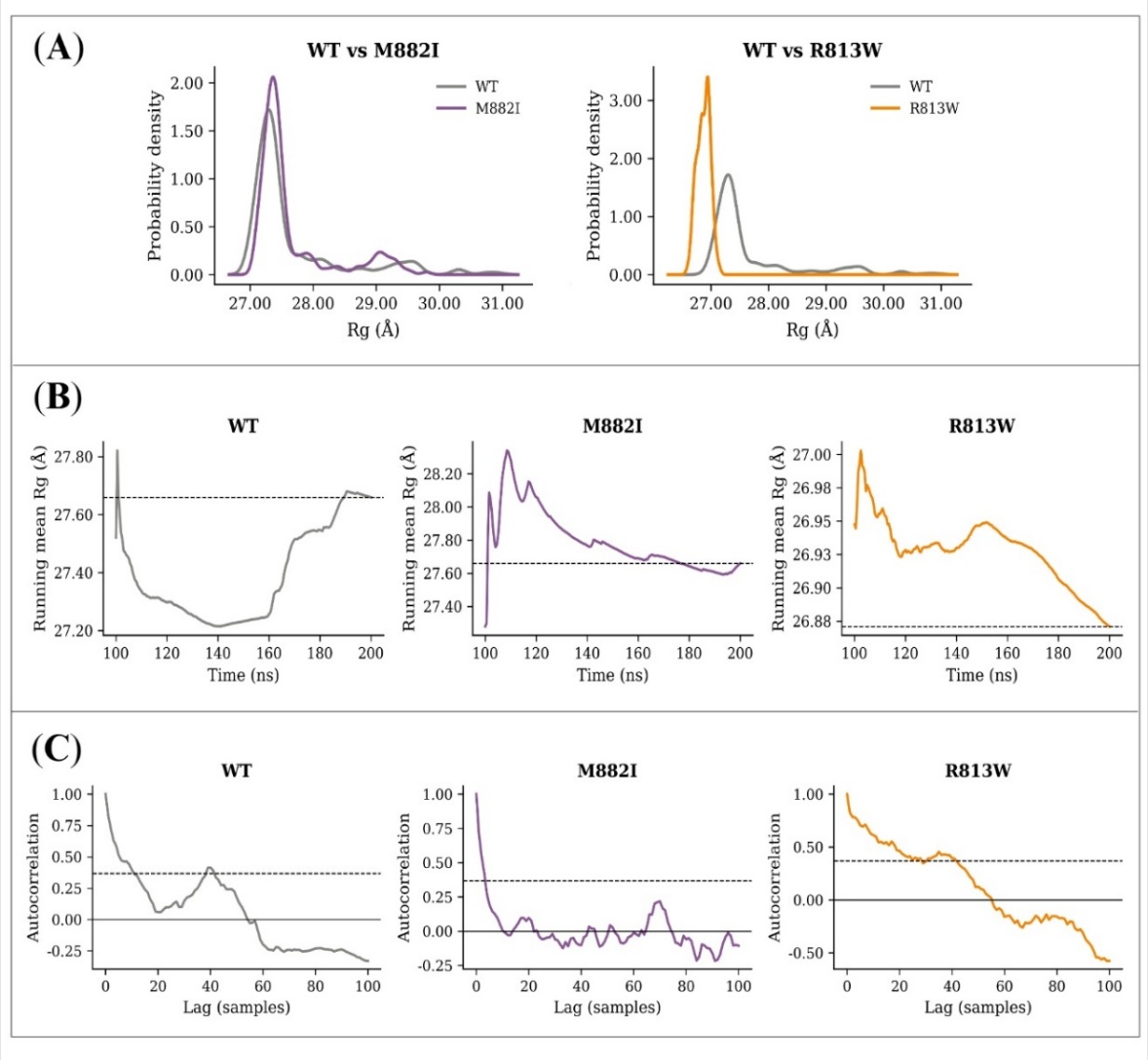

**Figure S35. Distributional and temporal diagnostics of RH domain R_g_.** (A) Pairwise WT-against-variant densities. (B) Running means with the final value dashed. (C) Autocorrelation against temporal lag with the 1/*e* level dashed.

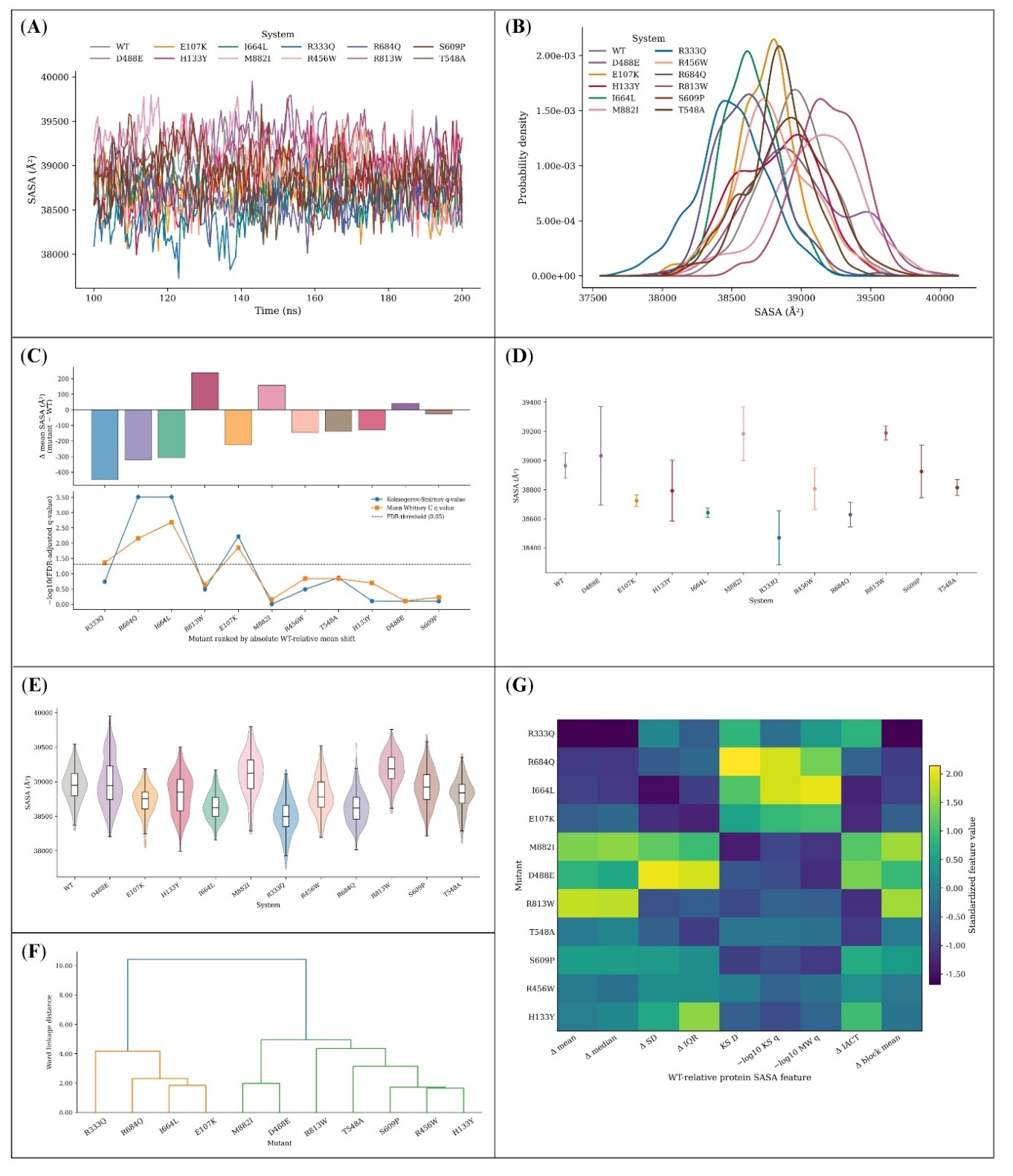

**Figure S36. SASA of full-length RUBICON.** (A) Replica-averaged time series and (B) its probability density. (C) WT-relative change in mean exposure (upper) above the BH-adjusted KS and MWU *q*-values plotted as $-{log}_{10}q$ (lower), with a dashed line at *q* = 0.05. (D) Block-averaged means with Student’s t-based 95% CIs. (E) Violin plots with embedded boxplots. (F) Ward-linkage dendrogram and (G) heatmap of standardised WT-relative features. The *q*-values compare replica-averaged profiles and are not trajectory-level tests; Table S18 gives the replica-level summaries.

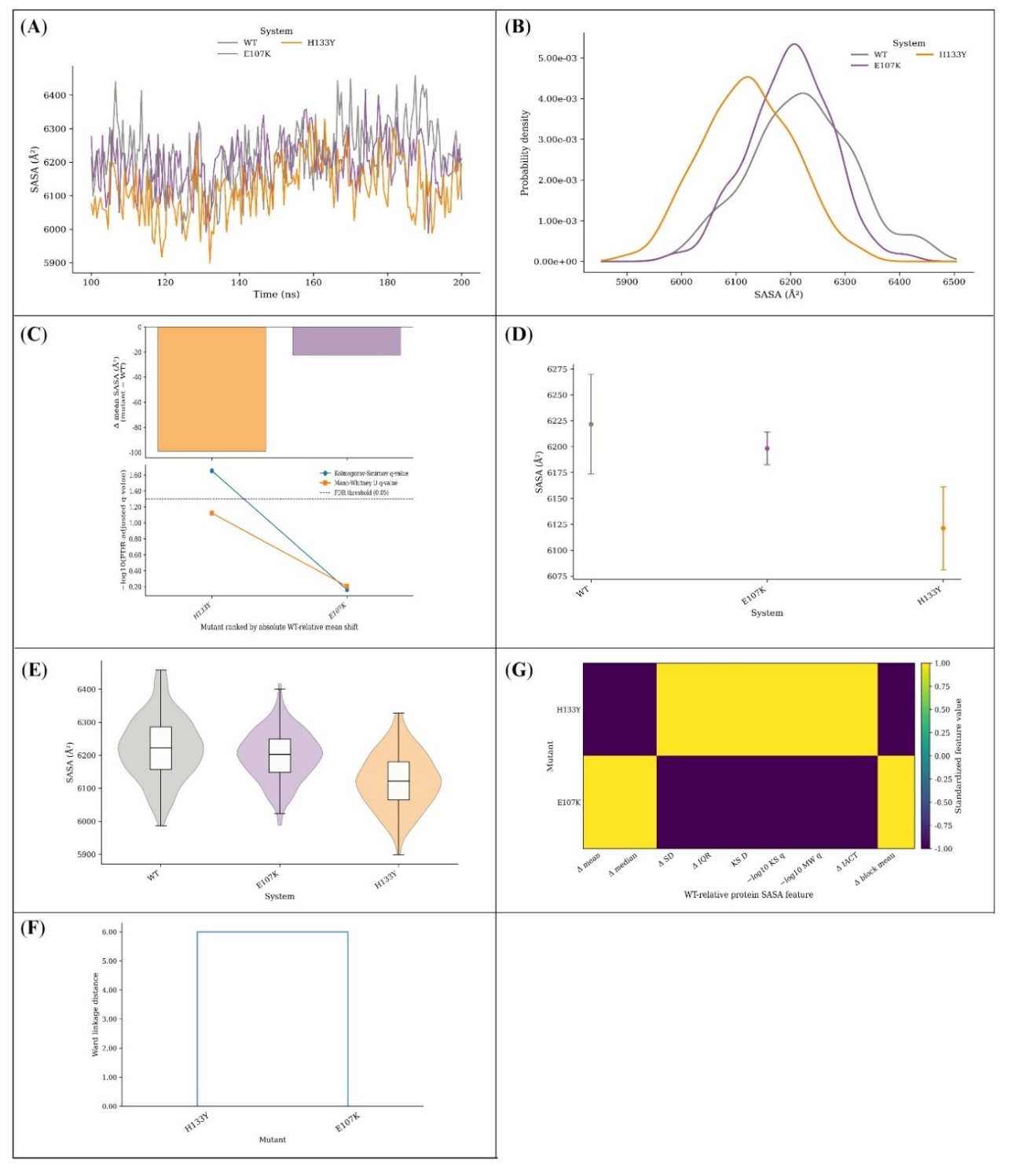

**Figure S37. SASA of the RUN domain.** Panel A traces the replica-averaged time series, panel B its probability density. Panel C places the WT-relative change in mean exposure above the BH-adjusted KS and MWU *q*-values as $-{log}_{10}q$, the dashed line marking *q* = 0.05. Block-averaged means with Student’s t-based 95% CIs follow in D, violin plots with embedded boxplots in E, and the standardised WT-relative features as a Ward-linkage dendrogram and heatmap in F and G. Regional SASA was computed within the intact protein, so neighbouring atoms continue to shield the domain boundaries. With only two variants here, panel F reduces to a single split and panel G to two extreme levels. The *q*-values are exploratory profile-level descriptors.

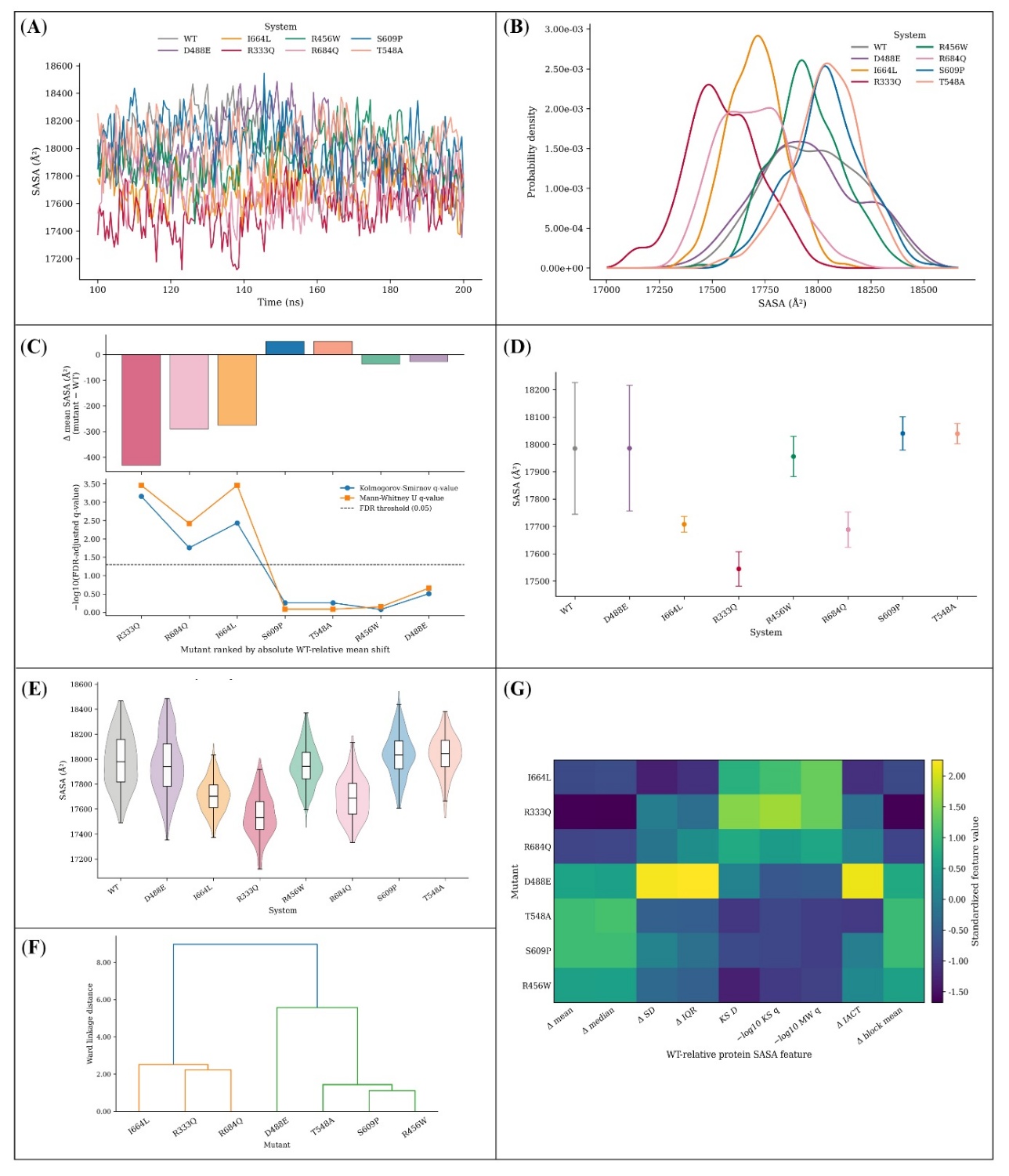

**Figure S38. SASA of the central region.** The replica-averaged time series appears in A and its probability density in B. Panel C reports the WT-relative change in mean exposure with the BH-adjusted KS and MWU *q*-values below it as $-{log}_{10}q$ and a dashed line at *q* = 0.05. The remaining panels give block-averaged means with Student’s t-based 95% CIs (D), violin plots with embedded boxplots (E), and a Ward-linkage dendrogram (F) with matching heatmap (G) of standardised WT-relative features. As throughout this series, the *q*-values describe separation between averaged profiles only.

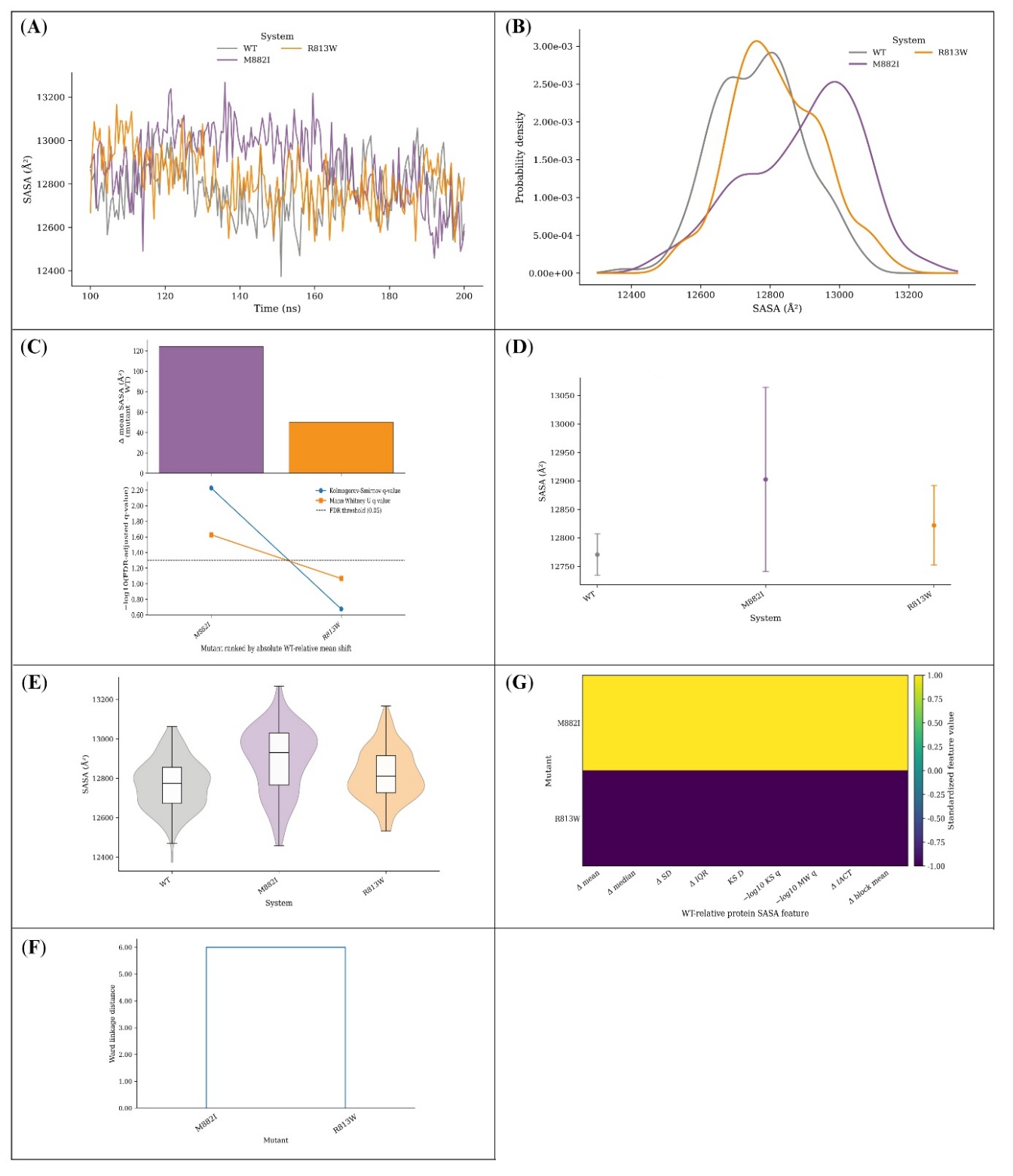

**Figure S39. SASA of the RH domain.** Panels A and B present the replica-averaged time series and its probability density. Panel C shows the WT-relative change in mean exposure over the BH-adjusted KS and MWU *q*-values as $-{log}_{10}q$, with *q* = 0.05 dashed. Block-averaged means with Student’s *t*-based 95% CIs are in D, violin plots with embedded boxplots in E, and the Ward-linkage dendrogram and heatmap of standardised WT-relative features in F and G. This domain also carries only two variants, limiting panel F to a pairwise separation. The *q*-values remain exploratory.

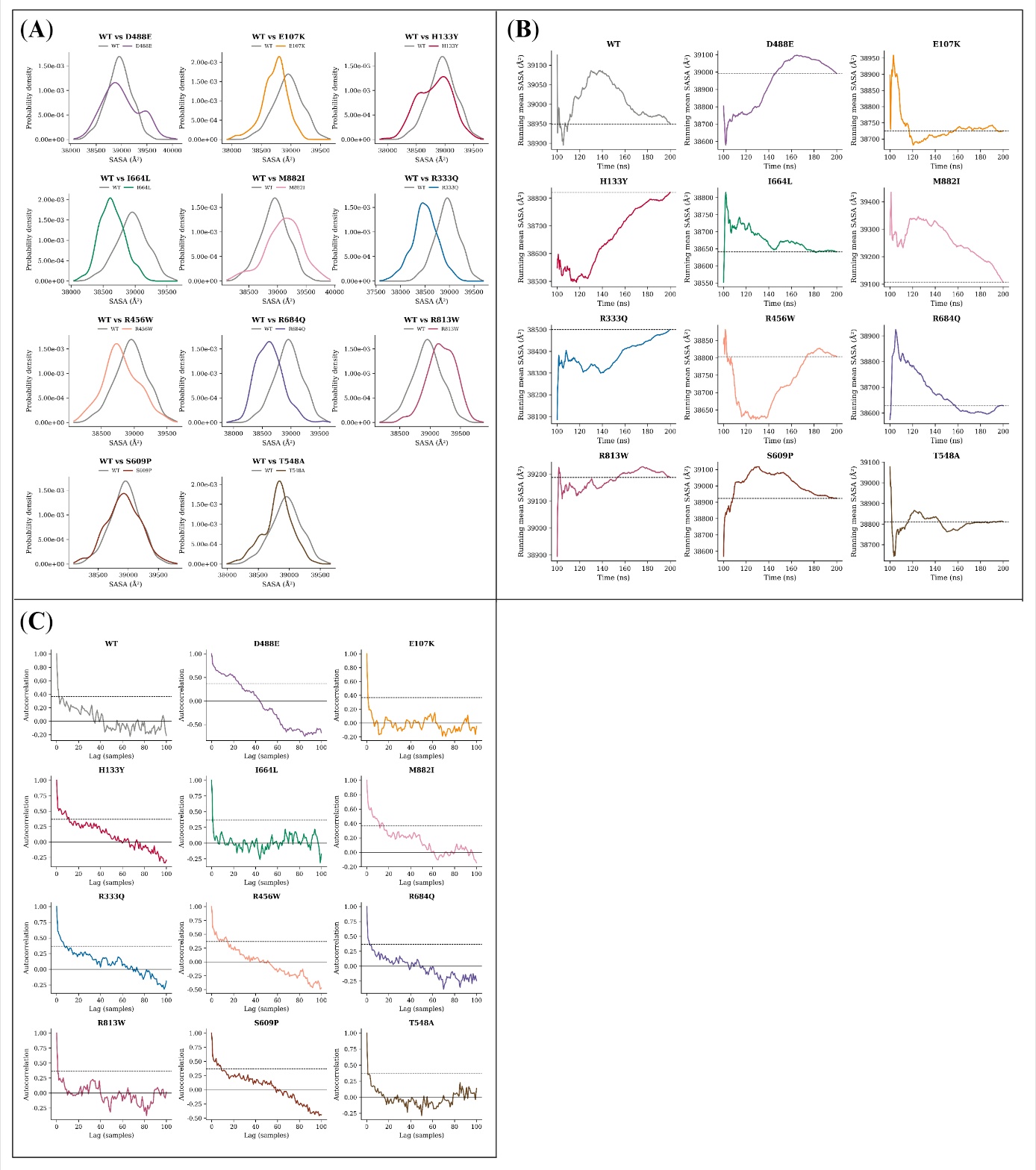

**Figure S40. Distributional and temporal diagnostics of full-length SASA.** (A) WT density overlaid on each variant in turn. (B) Running mean accumulated frame by frame, dashed at its final value. (C) Autocorrelation against temporal lag, dashed at 1/*e*. SASA was stored every 500 ps, so one lag unit corresponds to 500 ps in all four figures of this series.

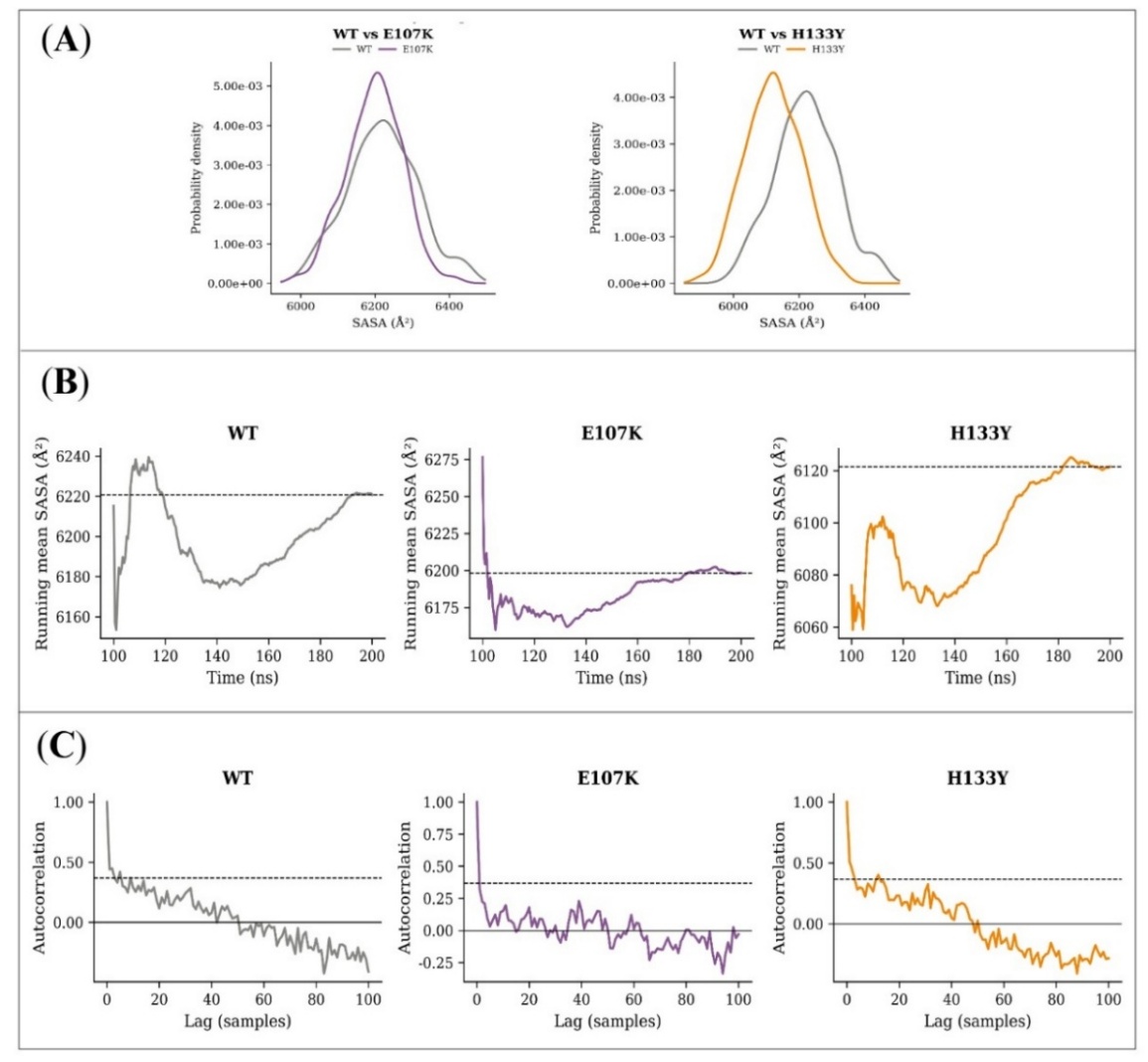

**Figure S41. Distributional and temporal diagnostics of RUN-domain SASA.** (A) Pairwise WT-against-variant probability densities. (B) Running means, dashed at the final profile mean. (C) Autocorrelation against temporal lag, dashed at 1/*e*.

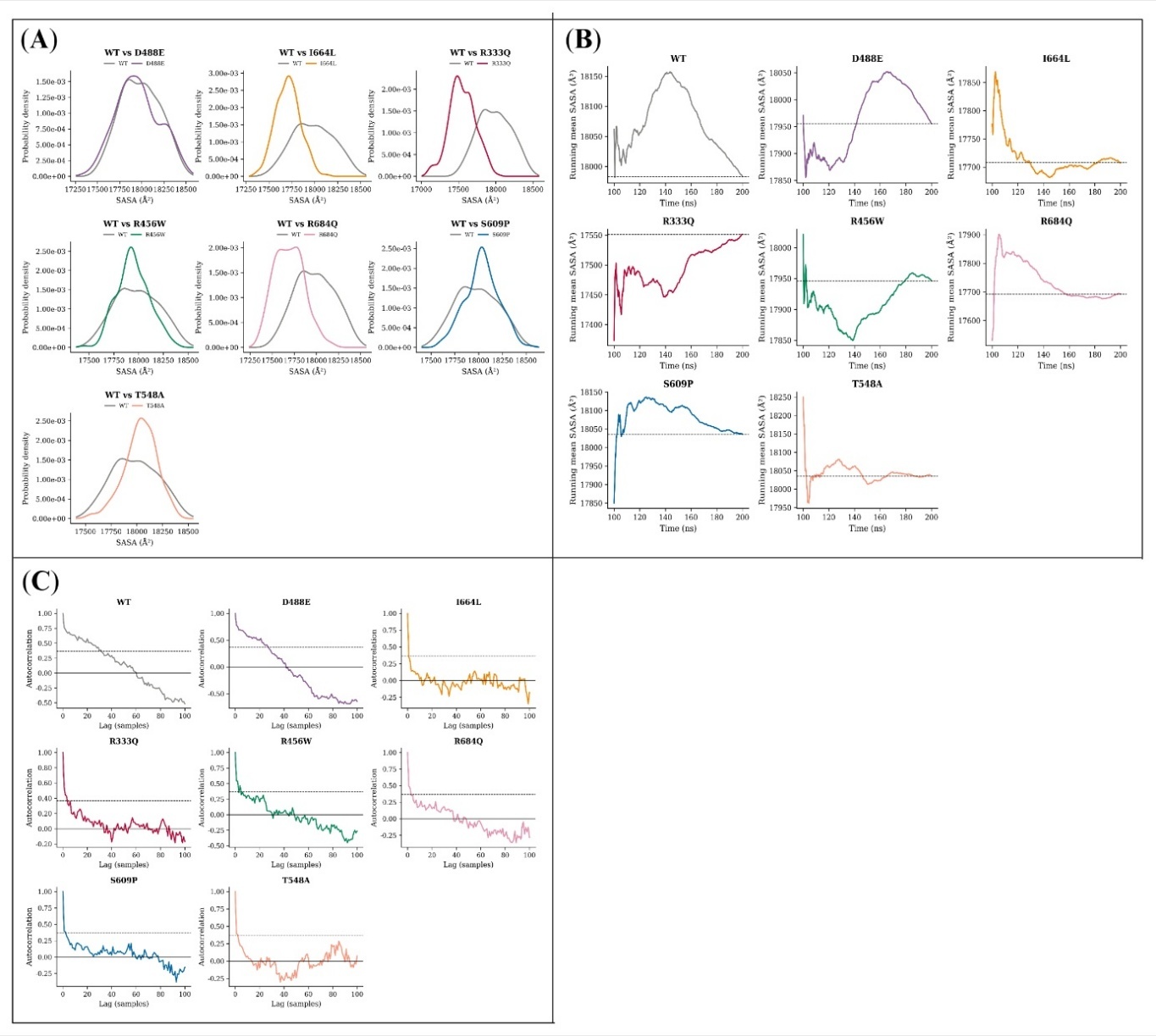

**Figure S42. Distributional and temporal diagnostics of central-region SASA.** (A) WT paired with each variant as overlaid probability densities. (B) Profile mean accumulated frame by frame, with the final value dashed. (C) Autocorrelation of each profile against temporal lag, with 1/*e* dashed.

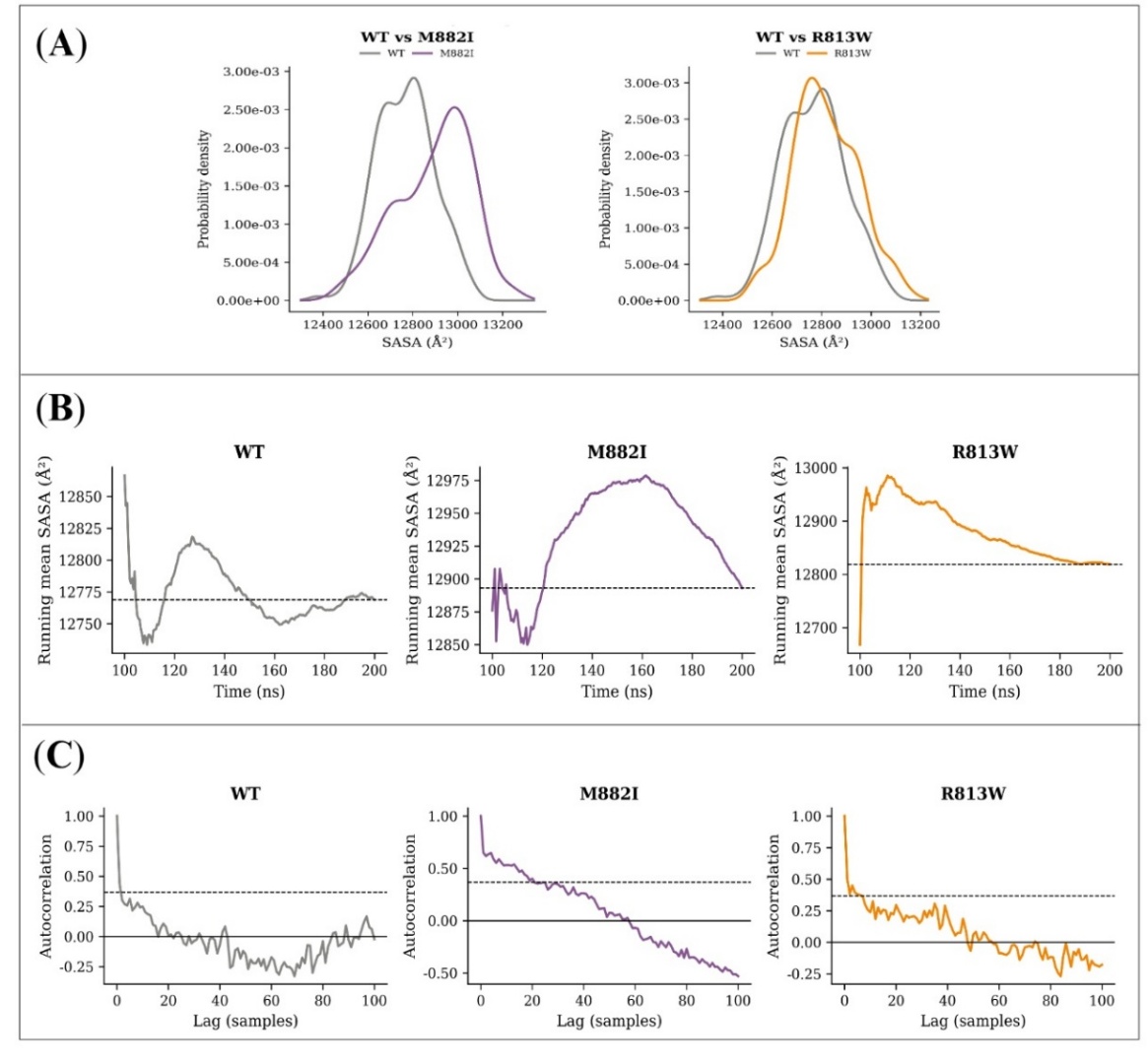

**Figure S43. Distributional and temporal diagnostics of RH domain SASA.** (A) Pairwise WT-against-variant densities. (B) Running means with the final value dashed. (C) Autocorrelation against temporal lag with the 1/*e* level dashed.

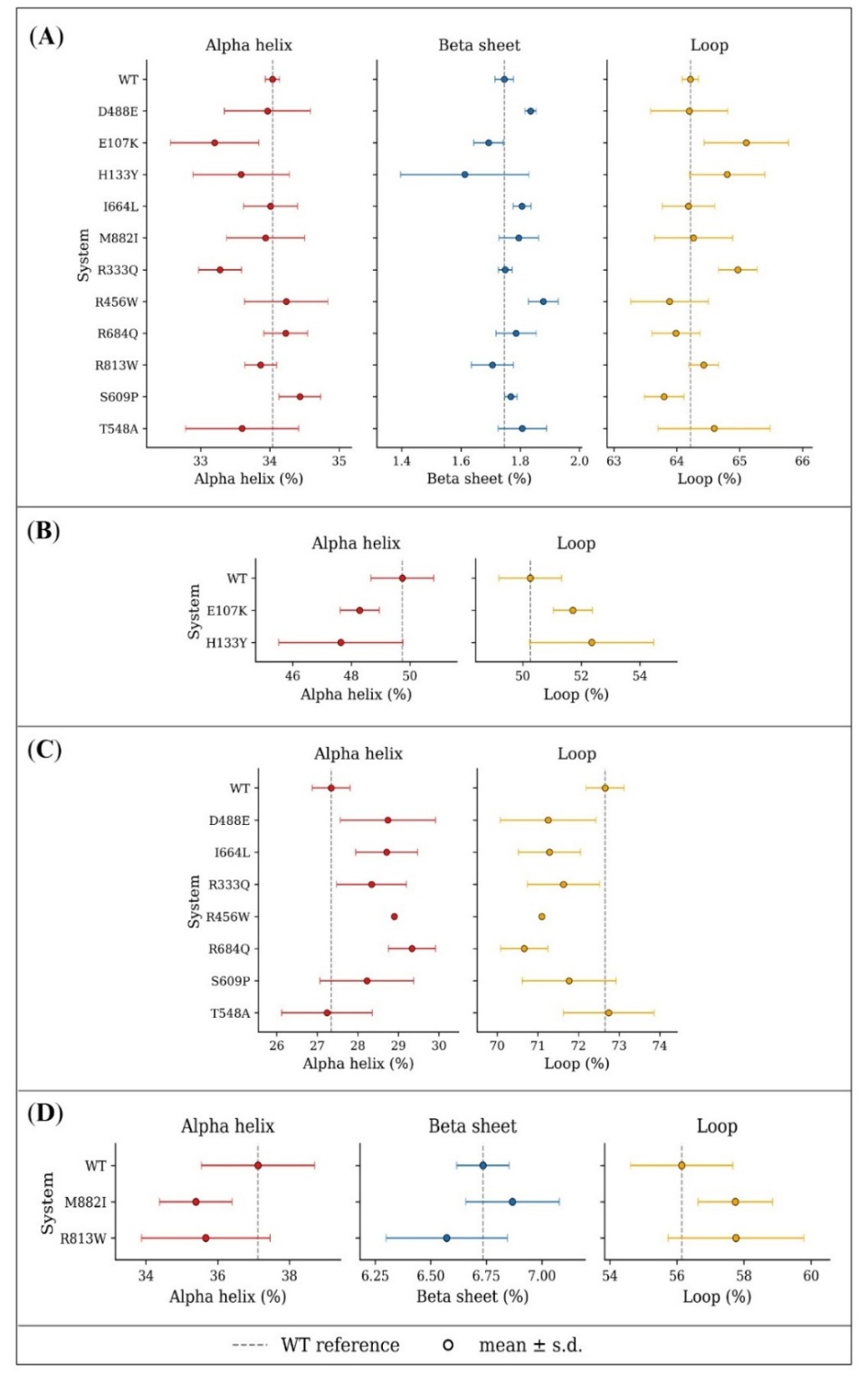

**Figure S44. SSC of the complete protein and the three predefined regions.** (A) Whole protein, (B) RUN domain, (C) central region and (D) RH domain. Helix, sheet and loop percentages are given as the mean ± sample SD of the three replica-level averages (ddof = 1), with dashed lines at the corresponding WT means. SSC is summarised at the replica level throughout, so no inferential testing was applied and no *q*-values are shown.

**Table S18.** Independent-replica summaries for the 88 predefined WT-versus-variant comparisons. Means, dispersions and effect sizes are computed across the three independent trajectories per system.

| **Region** | **Observable** | **Variant^a^** | **WT replica means^b^** | **Variant replica means^b^** | **WT mean ± SD^c^** | **Variant mean ± SD^c^** | **Δ^d^** | **\|Δ\| / WT replica-mean SD^e^** | **Hedges *g*^f^** |
| --- | --- | --- | --- | --- | --- | --- | --- | --- | --- |
| Whole protein | RMSD (Å) | E107K | 2.224; 2.734; 2.880 | 2.463; 2.185; 2.334 | 2.613 ± 0.345 | 2.327 ± 0.139 | -0.285 | 0.83 | -0.87 |
| Whole protein | RMSD (Å) | H133Y | 2.224; 2.734; 2.880 | 2.793; 2.462; 2.757 | 2.613 ± 0.345 | 2.671 ± 0.181 | 0.058 | 0.17 | 0.17 |
| Whole protein | RMSD (Å) | R333Q | 2.224; 2.734; 2.880 | 2.293; 2.216; 2.390 | 2.613 ± 0.345 | 2.299 ± 0.087 | -0.313 | 0.91 | -1.0 |
| Whole protein | RMSD (Å) | R456W | 2.224; 2.734; 2.880 | 2.302; 2.241; 2.349 | 2.613 ± 0.345 | 2.297 ± 0.054 | -0.315 | 0.91 | -1.02 |
| Whole protein | RMSD (Å) | D488E | 2.224; 2.734; 2.880 | 2.370; 2.448; 2.027 | 2.613 ± 0.345 | 2.282 ± 0.224 | -0.331 | 0.96 | -0.91 |
| Whole protein | RMSD (Å) | T548A | 2.224; 2.734; 2.880 | 2.123; 1.886; 2.286 | 2.613 ± 0.345 | 2.098 ± 0.201 | -0.514 | 1.49 | -1.46 |
| Whole protein | RMSD (Å) | S609P | 2.224; 2.734; 2.880 | 2.599; 2.190; 2.356 | 2.613 ± 0.345 | 2.382 ± 0.205 | -0.231 | 0.67 | -0.65 |
| Whole protein | RMSD (Å) | I664L | 2.224; 2.734; 2.880 | 1.891; 2.164; 2.368 | 2.613 ± 0.345 | 2.141 ± 0.239 | -0.472 | 1.37 | -1.27 |
| Whole protein | RMSD (Å) | R684Q | 2.224; 2.734; 2.880 | 2.438; 2.467; 2.460 | 2.613 ± 0.345 | 2.455 ± 0.015 | -0.158 | 0.46 | -0.52 |
| Whole protein | RMSD (Å) | R813W | 2.224; 2.734; 2.880 | 2.466; 2.770; 2.229 | 2.613 ± 0.345 | 2.488 ± 0.271 | -0.124 | 0.36 | -0.32 |
| Whole protein | RMSD (Å) | M882I | 2.224; 2.734; 2.880 | 2.306; 2.349; 2.366 | 2.613 ± 0.345 | 2.340 ± 0.031 | -0.272 | 0.79 | -0.89 |

**Table S18.** (continued)

| **Region** | **Observable** | **Variant^a^** | **WT replica means^b^** | **Variant replica means^b^** | **WT mean ± SD^c^** | **Variant mean ± SD^c^** | **Δ^d^** | **\|Δ\| / WT replica-mean SD^e^** | **Hedges *g*^f^** |
| --- | --- | --- | --- | --- | --- | --- | --- | --- | --- |
| Whole protein | Global-fit Cα RMSF (Å) | E107K | 1.097; 1.120; 1.023 | 1.015; 0.986; 0.943 | 1.080 ± 0.051 | 0.981 ± 0.036 | -0.099 | 1.94 | -1.79 |
| Whole protein | Global-fit Cα RMSF (Å) | H133Y | 1.097; 1.120; 1.023 | 1.011; 1.048; 1.003 | 1.080 ± 0.051 | 1.021 ± 0.024 | -0.059 | 1.16 | -1.19 |
| Whole protein | Global-fit Cα RMSF (Å) | R333Q | 1.097; 1.120; 1.023 | 0.973; 0.926; 1.025 | 1.080 ± 0.051 | 0.975 ± 0.049 | -0.105 | 2.06 | -1.68 |
| Whole protein | Global-fit Cα RMSF (Å) | R456W | 1.097; 1.120; 1.023 | 1.080; 1.019; 0.990 | 1.080 ± 0.051 | 1.030 ± 0.046 | -0.051 | 0.99 | -0.83 |
| Whole protein | Global-fit Cα RMSF (Å) | D488E | 1.097; 1.120; 1.023 | 1.112; 1.121; 0.938 | 1.080 ± 0.051 | 1.057 ± 0.103 | -0.023 | 0.45 | -0.23 |
| Whole protein | Global-fit Cα RMSF (Å) | T548A | 1.097; 1.120; 1.023 | 0.976; 0.949; 0.948 | 1.080 ± 0.051 | 0.958 ± 0.016 | -0.122 | 2.39 | -2.59 |
| Whole protein | Global-fit Cα RMSF (Å) | S609P | 1.097; 1.120; 1.023 | 0.907; 0.968; 1.124 | 1.080 ± 0.051 | 0.999 ± 0.112 | -0.081 | 1.58 | -0.74 |
| Whole protein | Global-fit Cα RMSF (Å) | I664L | 1.097; 1.120; 1.023 | 0.912; 1.042; 1.049 | 1.080 ± 0.051 | 1.001 ± 0.077 | -0.079 | 1.54 | -0.96 |
| Whole protein | Global-fit Cα RMSF (Å) | R684Q | 1.097; 1.120; 1.023 | 0.985; 0.967; 1.020 | 1.080 ± 0.051 | 0.991 ± 0.027 | -0.089 | 1.74 | -1.74 |
| Whole protein | Global-fit Cα RMSF (Å) | R813W | 1.097; 1.120; 1.023 | 1.092; 1.151; 0.911 | 1.080 ± 0.051 | 1.051 ± 0.125 | -0.029 | 0.57 | -0.24 |
| Whole protein | Global-fit Cα RMSF (Å) | M882I | 1.097; 1.120; 1.023 | 0.917; 1.062; 0.981 | 1.080 ± 0.051 | 0.986 ± 0.073 | -0.094 | 1.83 | -1.19 |

**Table S18.** (continued)

| **Region** | **Observable** | **Variant^a^** | **WT replica means^b^** | **Variant replica means^b^** | **WT mean ± SD^c^** | **Variant mean ± SD^c^** | **Δ^d^** | **\|Δ\| / WT replica-mean SD^e^** | **Hedges *g*^f^** |
| --- | --- | --- | --- | --- | --- | --- | --- | --- | --- |
| Whole protein | Rg (Å) | E107K | 31.933; 31.476; 32.072 | 33.202; 33.278; 32.039 | 31.827 ± 0.312 | 32.840 ± 0.695 | 1.012 | 3.25 | 1.5 |
| Whole protein | Rg (Å) | H133Y | 31.933; 31.476; 32.072 | 33.592; 31.262; 31.814 | 31.827 ± 0.312 | 32.223 ± 1.217 | 0.396 | 1.27 | 0.36 |
| Whole protein | Rg (Å) | R333Q | 31.933; 31.476; 32.072 | 33.332; 30.910; 32.366 | 31.827 ± 0.312 | 32.203 ± 1.219 | 0.376 | 1.2 | 0.34 |
| Whole protein | Rg (Å) | R456W | 31.933; 31.476; 32.072 | 32.685; 32.646; 33.668 | 31.827 ± 0.312 | 32.999 ± 0.579 | 1.172 | 3.76 | 2.02 |
| Whole protein | Rg (Å) | D488E | 31.933; 31.476; 32.072 | 33.291; 32.680; 31.110 | 31.827 ± 0.312 | 32.361 ± 1.125 | 0.533 | 1.71 | 0.52 |
| Whole protein | Rg (Å) | T548A | 31.933; 31.476; 32.072 | 31.610; 32.621; 33.151 | 31.827 ± 0.312 | 32.461 ± 0.783 | 0.634 | 2.03 | 0.85 |
| Whole protein | Rg (Å) | S609P | 31.933; 31.476; 32.072 | 31.819; 31.117; 33.282 | 31.827 ± 0.312 | 32.073 ± 1.104 | 0.246 | 0.79 | 0.24 |
| Whole protein | Rg (Å) | I664L | 31.933; 31.476; 32.072 | 31.841; 32.698; 31.974 | 31.827 ± 0.312 | 32.171 ± 0.461 | 0.344 | 1.1 | 0.7 |
| Whole protein | Rg (Å) | R684Q | 31.933; 31.476; 32.072 | 31.164; 32.333; 35.141 | 31.827 ± 0.312 | 32.879 ± 2.044 | 1.052 | 3.37 | 0.58 |
| Whole protein | Rg (Å) | R813W | 31.933; 31.476; 32.072 | 33.310; 32.351; 31.661 | 31.827 ± 0.312 | 32.441 ± 0.828 | 0.614 | 1.97 | 0.78 |
| Whole protein | Rg (Å) | M882I | 31.933; 31.476; 32.072 | 32.149; 32.163; 34.485 | 31.827 ± 0.312 | 32.933 ± 1.345 | 1.105 | 3.54 | 0.91 |

**Table S18.** (continued)

| **Region** | **Observable** | **Variant^a^** | **WT replica means^b^** | **Variant replica means^b^** | **WT mean ± SD^c^** | **Variant mean ± SD^c^** | **Δ^d^** | **\|Δ\| / WT replica-mean SD^e^** | **Hedges *g*^f^** |
| --- | --- | --- | --- | --- | --- | --- | --- | --- | --- |
| Whole protein | SASA (Å^2^) | E107K | 38758.6; 38812.7; 39276.1 | 38306.3; 38999.2; 38869.0 | 38949 ± 284 | 38725 ± 368 | -224.0 | 0.79 | -0.55 |
| Whole protein | SASA (Å^2^) | H133Y | 38758.6; 38812.7; 39276.1 | 38967.8; 38672.5; 38822.3 | 38949 ± 284 | 38821 ± 148 | -128.0 | 0.45 | -0.45 |
| Whole protein | SASA (Å^2^) | R333Q | 38758.6; 38812.7; 39276.1 | 38416.2; 38299.4; 38785.4 | 38949 ± 284 | 38500 ± 254 | -449.0 | 1.58 | -1.33 |
| Whole protein | SASA (Å^2^) | R456W | 38758.6; 38812.7; 39276.1 | 39142.5; 38789.8; 38478.4 | 38949 ± 284 | 38804 ± 332 | -146.0 | 0.51 | -0.38 |
| Whole protein | SASA (Å^2^) | D488E | 38758.6; 38812.7; 39276.1 | 39223.8; 39312.6; 38439.4 | 38949 ± 284 | 38992 ± 481 | 43.0 | 0.15 | 0.09 |
| Whole protein | SASA (Å^2^) | T548A | 38758.6; 38812.7; 39276.1 | 38518.9; 39048.4; 38865.9 | 38949 ± 284 | 38811 ± 269 | -138.0 | 0.49 | -0.4 |
| Whole protein | SASA (Å^2^) | S609P | 38758.6; 38812.7; 39276.1 | 39265.4; 38409.6; 39095.4 | 38949 ± 284 | 38923 ± 453 | -26.0 | 0.09 | -0.05 |
| Whole protein | SASA (Å^2^) | I664L | 38758.6; 38812.7; 39276.1 | 38572.7; 38954.7; 38396.8 | 38949 ± 284 | 38641 ± 285 | -308.0 | 1.08 | -0.86 |
| Whole protein | SASA (Å^2^) | R684Q | 38758.6; 38812.7; 39276.1 | 38313.9; 38268.5; 39303.0 | 38949 ± 284 | 38628 ± 585 | -321.0 | 1.13 | -0.56 |
| Whole protein | SASA (Å^2^) | R813W | 38758.6; 38812.7; 39276.1 | 40013.2; 39254.9; 38295.7 | 38949 ± 284 | 39188 ± 861 | 239.0 | 0.84 | 0.3 |
| Whole protein | SASA (Å^2^) | M882I | 38758.6; 38812.7; 39276.1 | 38750.8; 38985.2; 39585.8 | 38949 ± 284 | 39107 ± 431 | 158.0 | 0.56 | 0.35 |

**Table S18.** (continued)

| **Region** | **Observable** | **Variant^a^** | **WT replica means^b^** | **Variant replica means^b^** | **WT mean ± SD^c^** | **Variant mean ± SD^c^** | **Δ^d^** | **\|Δ\| / WT replica-mean SD^e^** | **Hedges *g*^f^** |
| --- | --- | --- | --- | --- | --- | --- | --- | --- | --- |
| RUN domain | RMSD (Å) | E107K | 1.211; 1.394; 1.745 | 1.712; 1.598; 1.610 | 1.450 ± 0.271 | 1.640 ± 0.063 | 0.19 | 0.7 | 0.77 |
| RUN domain | RMSD (Å) | H133Y | 1.211; 1.394; 1.745 | 1.378; 1.304; 1.754 | 1.450 ± 0.271 | 1.478 ± 0.241 | 0.029 | 0.11 | 0.09 |
| RUN domain | Global-fit Cα RMSF (Å) | E107K | 1.253; 1.132; 1.151 | 1.272; 1.105; 1.030 | 1.179 ± 0.065 | 1.135 ± 0.124 | -0.044 | 0.67 | -0.35 |
| RUN domain | Global-fit Cα RMSF (Å) | H133Y | 1.253; 1.132; 1.151 | 1.037; 1.289; 1.061 | 1.179 ± 0.065 | 1.129 ± 0.140 | -0.05 | 0.77 | -0.37 |
| RUN domain | Rg (Å) | E107K | 15.501; 15.454; 15.355 | 15.375; 15.500; 15.443 | 15.437 ± 0.075 | 15.440 ± 0.062 | 0.003 | 0.04 | 0.04 |
| RUN domain | Rg (Å) | H133Y | 15.501; 15.454; 15.355 | 15.401; 15.318; 15.364 | 15.437 ± 0.075 | 15.361 ± 0.041 | -0.076 | 1.01 | -1.0 |
| RUN domain | SASA (Å^2^) | E107K | 6255.0; 6180.3; 6227.0 | 6083.6; 6228.6; 6282.8 | 6221 ± 38 | 6198 ± 103 | -22.0 | 0.6 | -0.23 |
| RUN domain | SASA (Å^2^) | H133Y | 6255.0; 6180.3; 6227.0 | 6211.9; 6084.5; 6068.3 | 6221 ± 38 | 6122 ± 79 | -99.0 | 2.63 | -1.29 |
| Central region | RMSD (Å) | R333Q | 2.033; 2.439; 2.739 | 2.060; 1.952; 2.335 | 2.404 ± 0.355 | 2.116 ± 0.198 | -0.288 | 0.81 | -0.8 |
| Central region | RMSD (Å) | R456W | 2.033; 2.439; 2.739 | 2.119; 1.879; 2.028 | 2.404 ± 0.355 | 2.009 ± 0.121 | -0.395 | 1.11 | -1.19 |
| Central region | RMSD (Å) | D488E | 2.033; 2.439; 2.739 | 2.087; 2.191; 2.099 | 2.404 ± 0.355 | 2.126 ± 0.057 | -0.278 | 0.78 | -0.87 |

**Table S18.** (continued)

| **Region** | **Observable** | **Variant^a^** | **WT replica means^b^** | **Variant replica means^b^** | **WT mean ± SD^c^** | **Variant mean ± SD^c^** | **Δ^d^** | **\|Δ\| / WT replica-mean SD^e^** | **Hedges *g*^f^** |
| --- | --- | --- | --- | --- | --- | --- | --- | --- | --- |
| Central region | RMSD (Å) | T548A | 2.033; 2.439; 2.739 | 1.975; 1.732; 2.118 | 2.404 ± 0.355 | 1.941 ± 0.195 | -0.462 | 1.3 | -1.29 |
| Central region | RMSD (Å) | S609P | 2.033; 2.439; 2.739 | 2.446; 2.080; 2.325 | 2.404 ± 0.355 | 2.283 ± 0.186 | -0.12 | 0.34 | -0.34 |
| Central region | RMSD (Å) | I664L | 2.033; 2.439; 2.739 | 1.765; 1.918; 2.084 | 2.404 ± 0.355 | 1.922 ± 0.160 | -0.481 | 1.36 | -1.4 |
| Central region | RMSD (Å) | R684Q | 2.033; 2.439; 2.739 | 2.261; 2.352; 2.487 | 2.404 ± 0.355 | 2.367 ± 0.114 | -0.037 | 0.1 | -0.11 |
| Central region | Global-fit Cα RMSF (Å) | R333Q | 0.939; 1.021; 0.944 | 0.886; 0.811; 0.949 | 0.968 ± 0.046 | 0.882 ± 0.069 | -0.086 | 1.87 | -1.17 |
| Central region | Global-fit Cα RMSF (Å) | R456W | 0.939; 1.021; 0.944 | 1.043; 0.945; 0.897 | 0.968 ± 0.046 | 0.962 ± 0.074 | -0.006 | 0.14 | -0.08 |
| Central region | Global-fit Cα RMSF (Å) | D488E | 0.939; 1.021; 0.944 | 1.046; 0.977; 0.905 | 0.968 ± 0.046 | 0.976 ± 0.071 | 0.008 | 0.17 | 0.11 |
| Central region | Global-fit Cα RMSF (Å) | T548A | 0.939; 1.021; 0.944 | 0.936; 0.905; 0.913 | 0.968 ± 0.046 | 0.918 ± 0.016 | -0.05 | 1.09 | -1.16 |
| Central region | Global-fit Cα RMSF (Å) | S609P | 0.939; 1.021; 0.944 | 0.801; 0.951; 1.021 | 0.968 ± 0.046 | 0.924 ± 0.113 | -0.044 | 0.95 | -0.41 |
| Central region | Global-fit Cα RMSF (Å) | I664L | 0.939; 1.021; 0.944 | 0.836; 0.991; 0.947 | 0.968 ± 0.046 | 0.925 ± 0.080 | -0.044 | 0.95 | -0.53 |

**Table S18.** (continued)

| **Region** | **Observable** | **Variant^a^** | **WT replica means^b^** | **Variant replica means^b^** | **WT mean ± SD^c^** | **Variant mean ± SD^c^** | **Δ^d^** | **\|Δ\| / WT replica-mean SD^e^** | **Hedges *g*^f^** |
| --- | --- | --- | --- | --- | --- | --- | --- | --- | --- |
| Central region | Global-fit Cα RMSF (Å) | R684Q | 0.939; 1.021; 0.944 | 0.899; 0.835; 0.967 | 0.968 ± 0.046 | 0.901 ± 0.066 | -0.068 | 1.47 | -0.95 |
| Central region | Rg (Å) | R333Q | 22.919; 22.919; 23.054 | 22.940; 22.924; 22.963 | 22.964 ± 0.078 | 22.942 ± 0.020 | -0.022 | 0.28 | -0.31 |
| Central region | Rg (Å) | R456W | 22.919; 22.919; 23.054 | 23.002; 22.929; 22.974 | 22.964 ± 0.078 | 22.968 ± 0.036 | 0.004 | 0.05 | 0.05 |
| Central region | Rg (Å) | D488E | 22.919; 22.919; 23.054 | 22.887; 23.029; 22.856 | 22.964 ± 0.078 | 22.924 ± 0.092 | -0.04 | 0.52 | -0.38 |
| Central region | Rg (Å) | T548A | 22.919; 22.919; 23.054 | 22.928; 22.990; 23.066 | 22.964 ± 0.078 | 22.994 ± 0.069 | 0.03 | 0.39 | 0.33 |
| Central region | Rg (Å) | S609P | 22.919; 22.919; 23.054 | 23.072; 23.015; 22.838 | 22.964 ± 0.078 | 22.975 ± 0.122 | 0.011 | 0.14 | 0.09 |
| Central region | Rg (Å) | I664L | 22.919; 22.919; 23.054 | 22.943; 22.958; 22.929 | 22.964 ± 0.078 | 22.943 ± 0.015 | -0.021 | 0.27 | -0.3 |
| Central region | Rg (Å) | R684Q | 22.919; 22.919; 23.054 | 22.925; 22.885; 22.905 | 22.964 ± 0.078 | 22.905 ± 0.020 | -0.059 | 0.76 | -0.83 |
| Central region | SASA (Å^2^) | R333Q | 17837.3; 18076.4; 18035.5 | 17399.8; 17335.6; 17918.5 | 17983 ± 128 | 17551 ± 320 | -432.0 | 3.38 | -1.42 |
| Central region | SASA (Å^2^) | R456W | 17837.3; 18076.4; 18035.5 | 18092.5; 17937.6; 17808.8 | 17983 ± 128 | 17946 ± 142 | -37.0 | 0.29 | -0.22 |

**Table S18.** (continued)

| **Region** | **Observable** | **Variant^a^** | **WT replica means^b^** | **Variant replica means^b^** | **WT mean ± SD^c^** | **Variant mean ± SD^c^** | **Δ^d^** | **\|Δ\| / WT replica-mean SD^e^** | **Hedges *g*^f^** |
| --- | --- | --- | --- | --- | --- | --- | --- | --- | --- |
| Central region | SASA (Å^2^) | D488E | 17837.3; 18076.4; 18035.5 | 18075.4; 18130.5; 17660.6 | 17983 ± 128 | 17955 ± 257 | -28.0 | 0.22 | -0.11 |
| Central region | SASA (Å^2^) | T548A | 17837.3; 18076.4; 18035.5 | 17945.6; 18027.8; 18132.4 | 17983 ± 128 | 18035 ± 94 | 52.0 | 0.41 | 0.37 |
| Central region | SASA (Å^2^) | S609P | 17837.3; 18076.4; 18035.5 | 18169.2; 17768.8; 18169.0 | 17983 ± 128 | 18036 ± 231 | 53.0 | 0.41 | 0.23 |
| Central region | SASA (Å^2^) | I664L | 17837.3; 18076.4; 18035.5 | 17494.8; 17975.0; 17653.8 | 17983 ± 128 | 17708 ± 245 | -275.0 | 2.15 | -1.13 |
| Central region | SASA (Å^2^) | R684Q | 17837.3; 18076.4; 18035.5 | 17769.1; 17376.8; 17932.1 | 17983 ± 128 | 17693 ± 285 | -290.0 | 2.27 | -1.05 |
| RH domain | RMSD (Å) | R813W | 2.182; 2.665; 2.341 | 2.415; 2.346; 2.035 | 2.396 ± 0.246 | 2.265 ± 0.202 | -0.131 | 0.53 | -0.46 |
| RH domain | RMSD (Å) | M882I | 2.182; 2.665; 2.341 | 2.033; 2.415; 2.163 | 2.396 ± 0.246 | 2.203 ± 0.194 | -0.192 | 0.78 | -0.7 |

**Table S18.** (continued)

| **Region** | **Observable** | **Variant^a^** | **WT replica means^b^** | **Variant replica means^b^** | **WT mean ± SD^c^** | **Variant mean ± SD^c^** | **Δ^d^** | **\|Δ\| / WT replica-mean SD^e^** | **Hedges *g*^f^** |
| --- | --- | --- | --- | --- | --- | --- | --- | --- | --- |
| RH domain | Global-fit Cα RMSF (Å) | R813W | 1.381; 1.370; 1.111 | 1.305; 1.212; 0.988 | 1.287 ± 0.152 | 1.168 ± 0.163 | -0.119 | 0.78 | -0.6 |
| RH domain | Global-fit Cα RMSF (Å) | M882I | 1.381; 1.370; 1.111 | 1.059; 1.169; 0.990 | 1.287 ± 0.152 | 1.073 ± 0.090 | -0.214 | 1.4 | -1.37 |
| RH domain | Rg (Å) | R813W | 27.763; 27.638; 27.577 | 26.968; 26.651; 26.994 | 27.660 ± 0.095 | 26.871 ± 0.191 | -0.789 | 8.34 | -4.18 |
| RH domain | Rg (Å) | M882I | 27.763; 27.638; 27.577 | 27.658; 26.954; 28.367 | 27.660 ± 0.095 | 27.660 ± 0.706 | 0.0 | 0.0 | 0.0 |
| RH domain | SASA (Å²) | R813W | 12688.7; 12544.0; 13074.0 | 13058.2; 12831.2; 12567.4 | 12769 ± 274 | 12819 ± 246 | 50.0 | 0.18 | 0.15 |
| RH domain | SASA (Å²) | M882I | 12688.7; 12544.0; 13074.0 | 12939.3; 12789.3; 12950.9 | 12769 ± 274 | 12893 ± 90 | 124.0 | 0.45 | 0.49 |

^a^Regional rows list only the variants located within the interval concerned. ^b^Mean of each independently initiated trajectory over the final 100 ns. ^c^Mean ± sample SD across the three replica means (ddof =1); this differs from the pointwise between-replica scale in Tables S4 to S7. No trajectory-level *P* or *q* value was calculated. ^d^Variant mean minus WT mean, in Å for RMSD, RMSF and R_g_ and Å^2^ for SASA. ^e^Ratio of the absolute difference to the sample SD of the three WT replica means. It is a descriptive scale with no significance threshold and, at *n* = 3, is sensitive to the observed WT spread. ^f^Descriptive standardised effect size. RMSF refers throughout to the global-fit analysis.

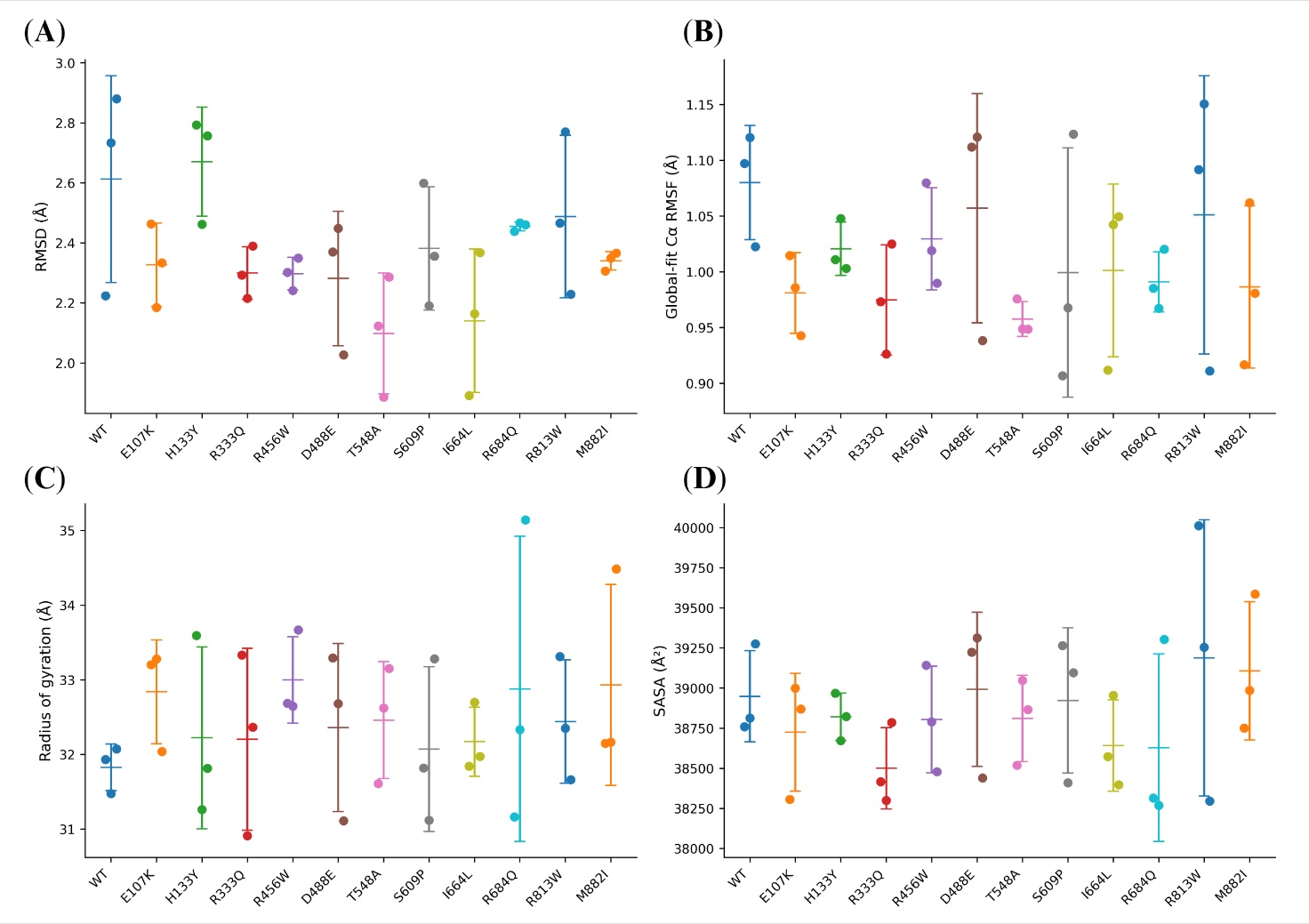

**Figure S45. Independent-replica summaries of whole-protein MD observables.** (A) Backbone RMSD, (B) global-fit Cα RMSF, (C) R_g_, and (D) SASA, each over the final 100 ns of the production trajectories. Each point is the mean of one independently initiated 200 ns trajectory (*n* = 3 per system); bars and whiskers give the mean ± sample SD (ddof = 1). RMSD used local fitting on the whole-protein selection; RMSF used complete-protein backbone fitting. No profile-level *q*-values are shown.

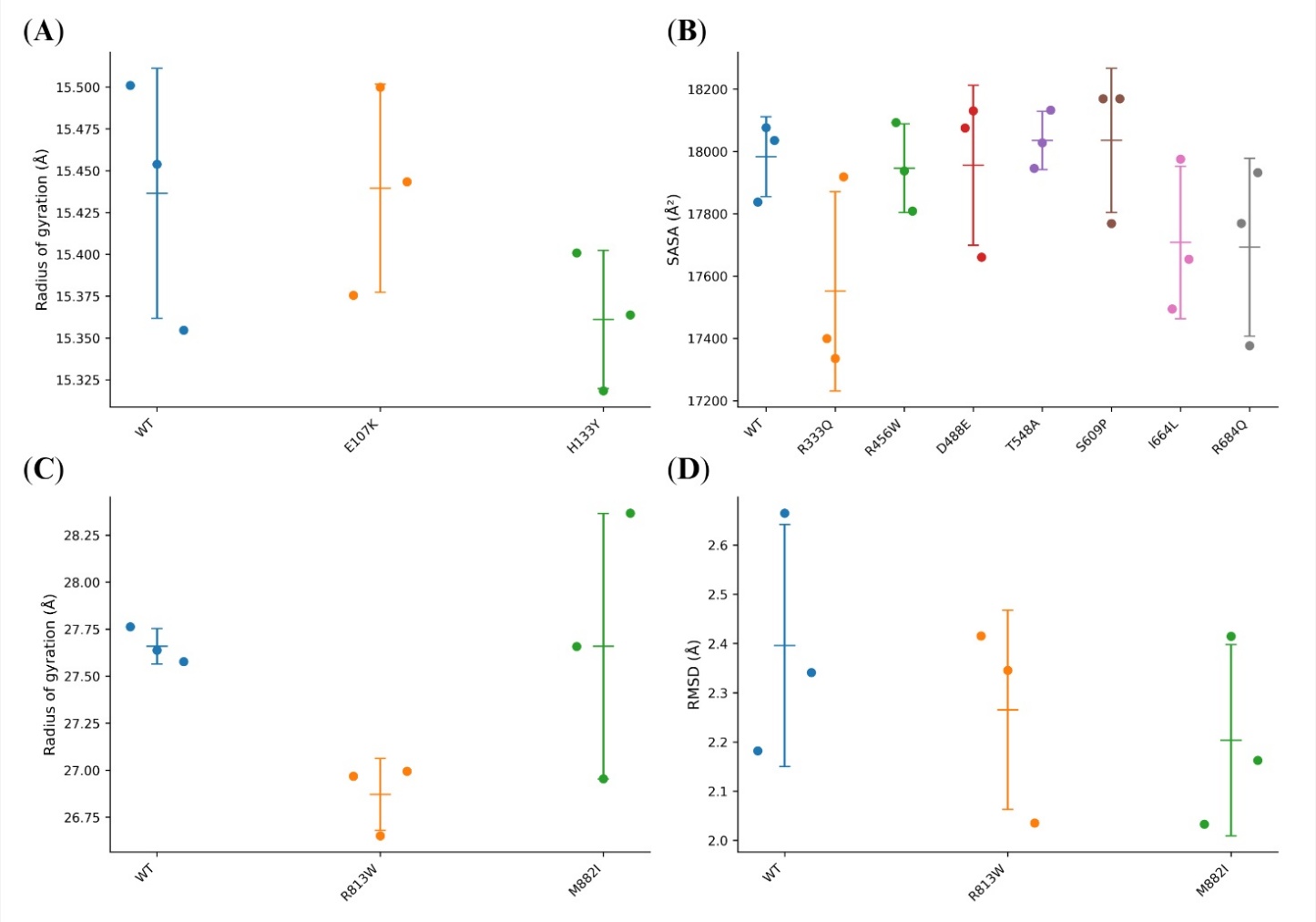

**Figure S46. Independent-replica summaries of selected regional signatures.** (A) RUN-domain R_g_ for WT, E107K and H133Y. (B) Central-region SASA for WT and the seven variants located in that interval. (C) RH-domain R_g_ and (D) RH-domain RMSD, both for WT, R813W and M882I. Each point is one independent trajectory mean; bars give the mean ± sample SD (ddof = 1). These panels show the principal region-specific descriptive responses; no profile-level significance annotation is applied.

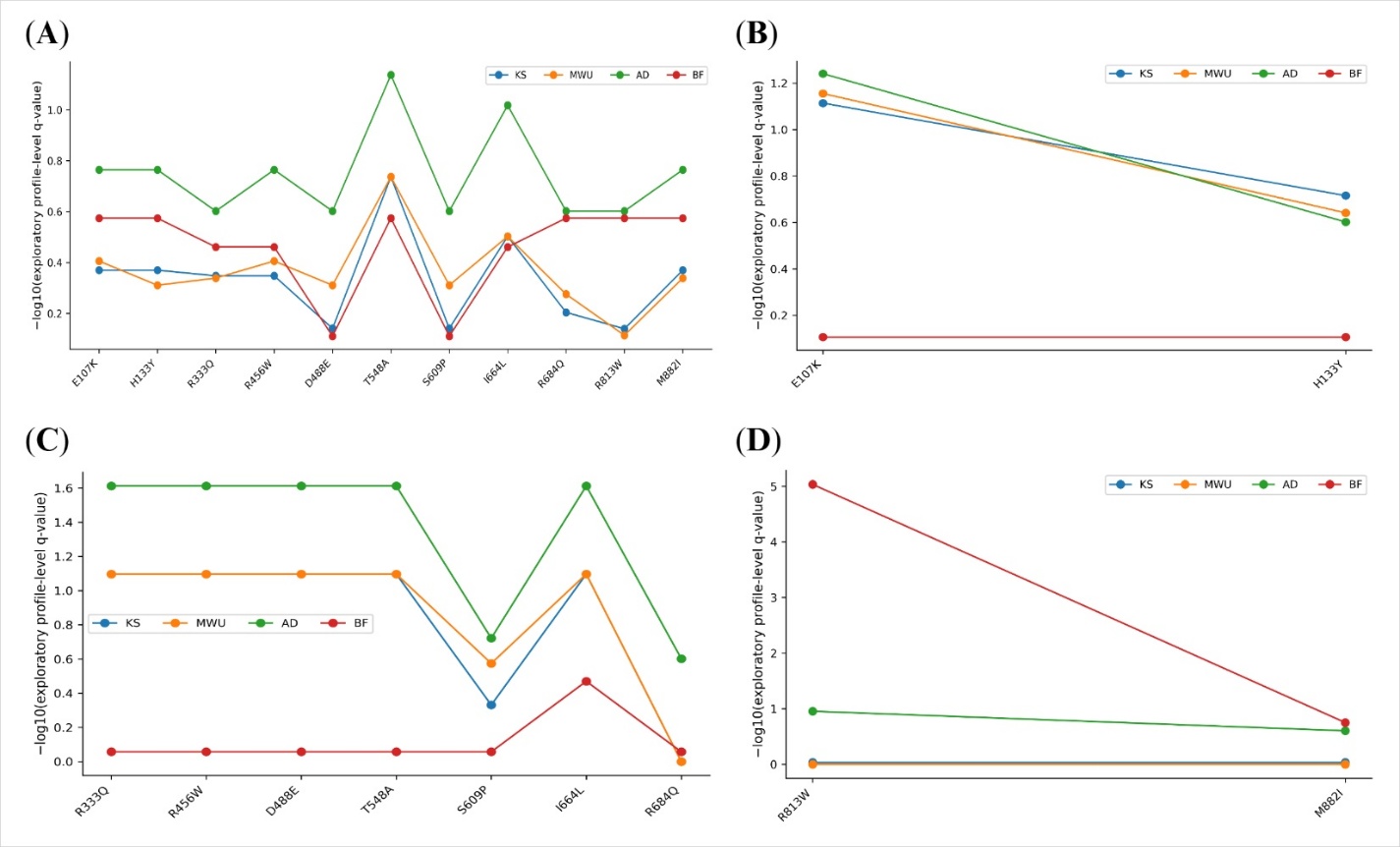

**Figure S47.** **Exploratory profile-level adjusted values for RMSD.** Panels give (A) the whole protein, (B) the RUN domain, (C) the central region and (D) the RH domain. Each panel plots the BH-adjusted *q*-values of the four tests (KS, MWU, AD and BF) as $-{log}_{10}q$ against variant, so higher points correspond to smaller adjusted *q*-values under the test concerned. The tests were applied to replica-averaged, autocorrelation-subsampled profiles and carry no trajectory-level inference. The corresponding replica-level values are given in Table S18.

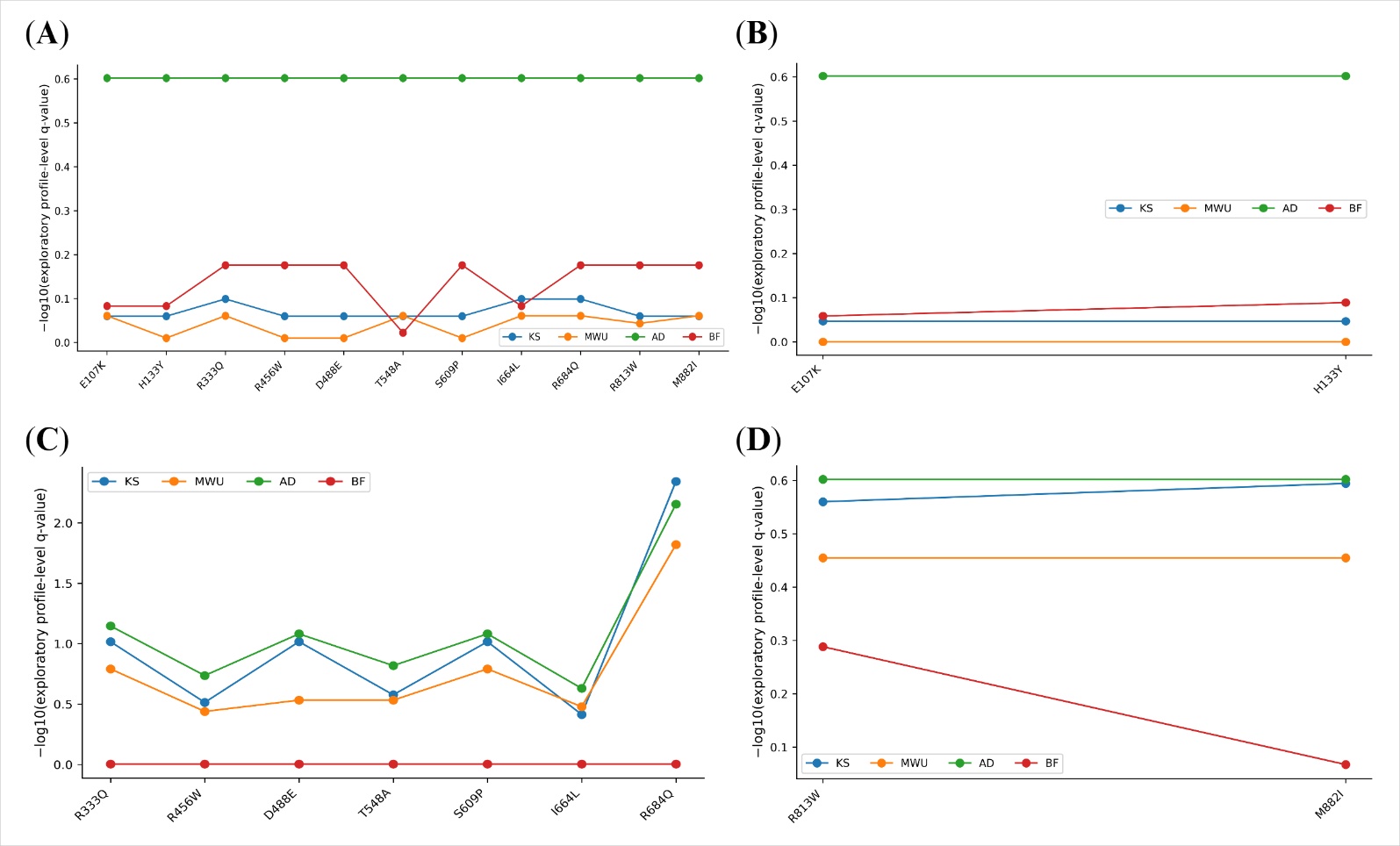

**Figure S48. Exploratory profile-level adjusted values for global-fit Cα RMSF.** These tests compare marginal RMSF distributions and do not preserve same-residue WT-variant pairing. The paired difference profiles are shown in Figures S43 to S46.

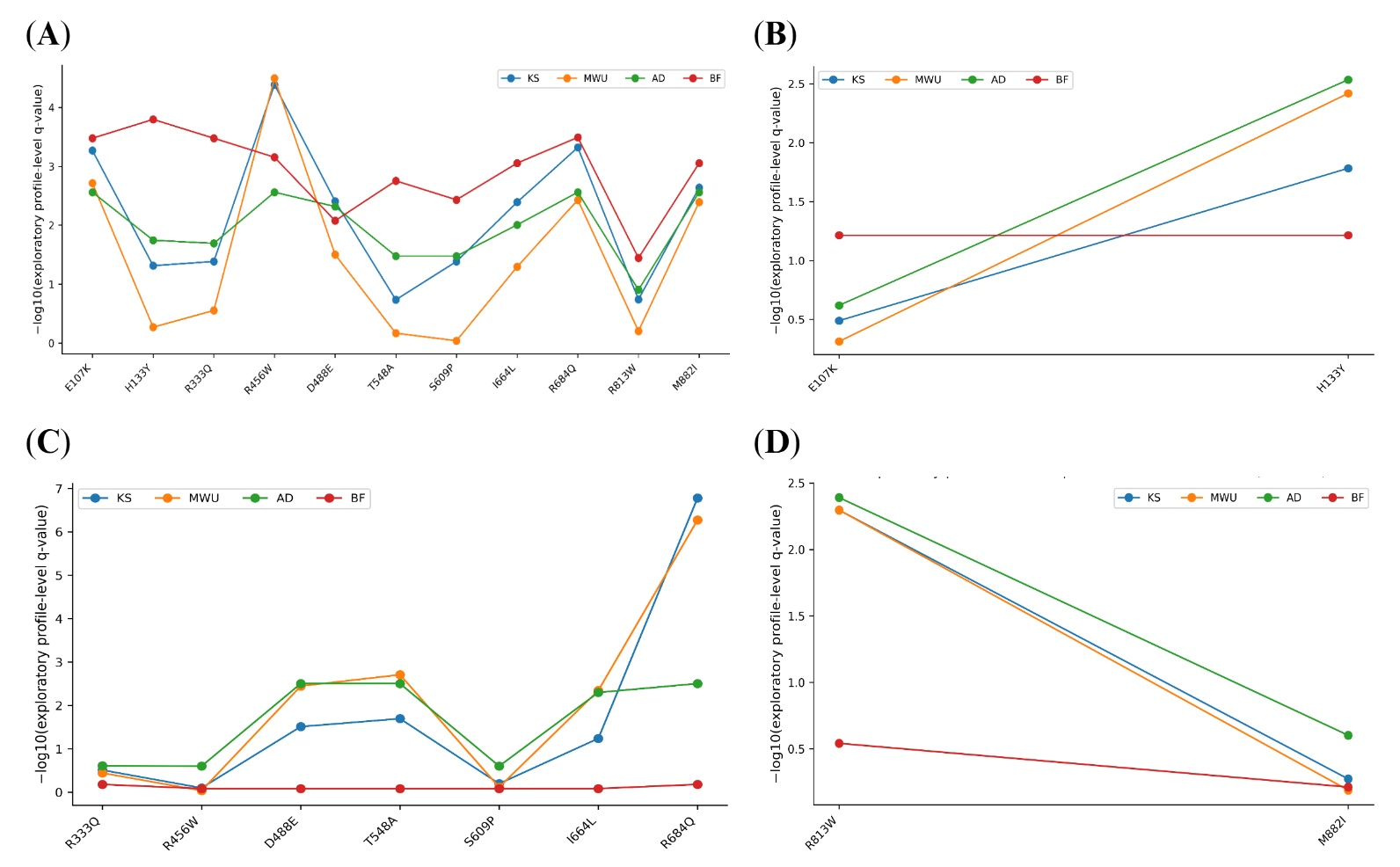

**Figure S49. Exploratory profile-level adjusted values for R_g_.** R_g_ gives the largest *q*-value excursions of the four observables, consistent with Tables S6 and S17.

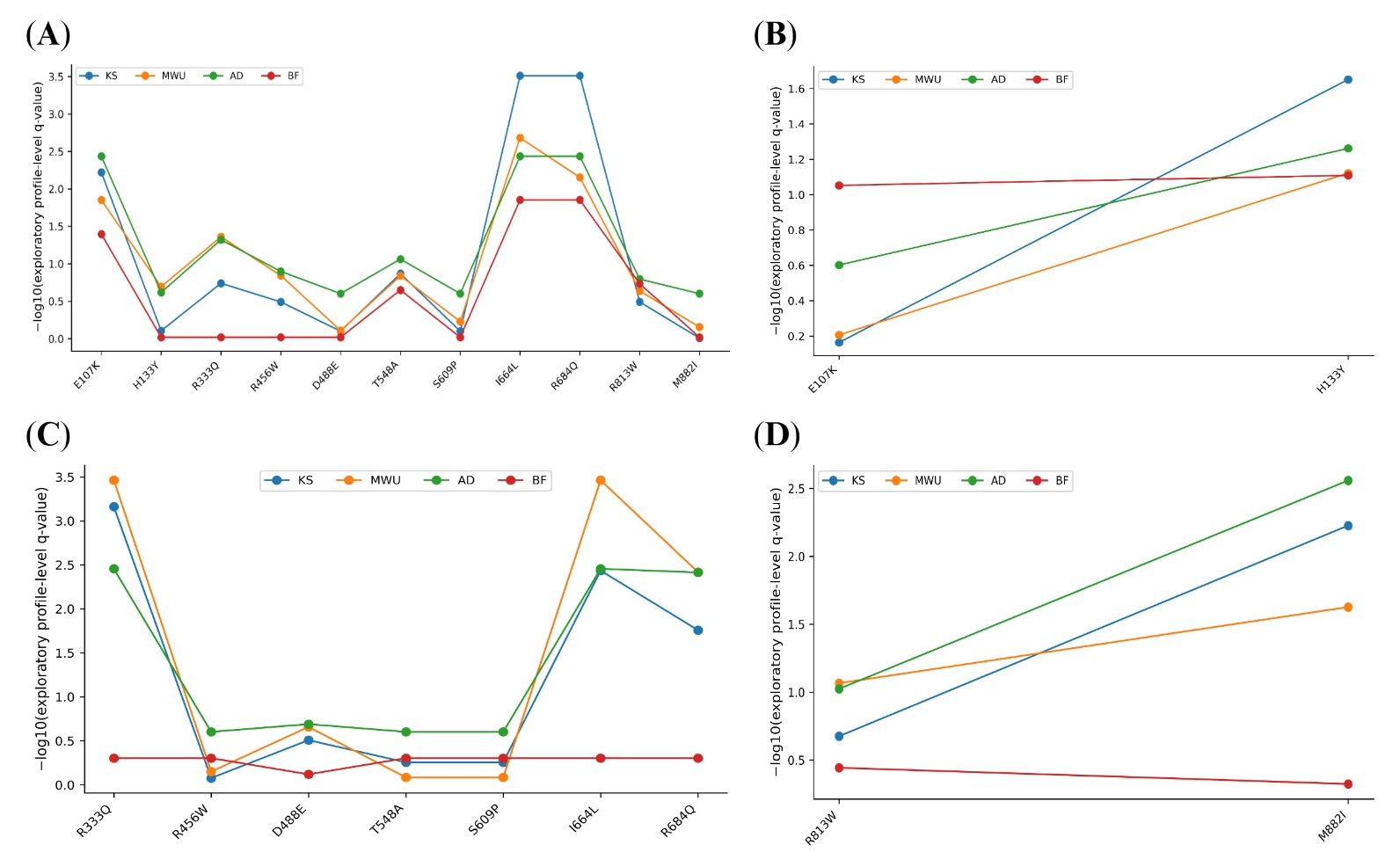

**Figure S50. Exploratory profile-level adjusted values for SASA.** As elsewhere in this series, the samples tested are averaged profiles, so between-trajectory variation is not represented.

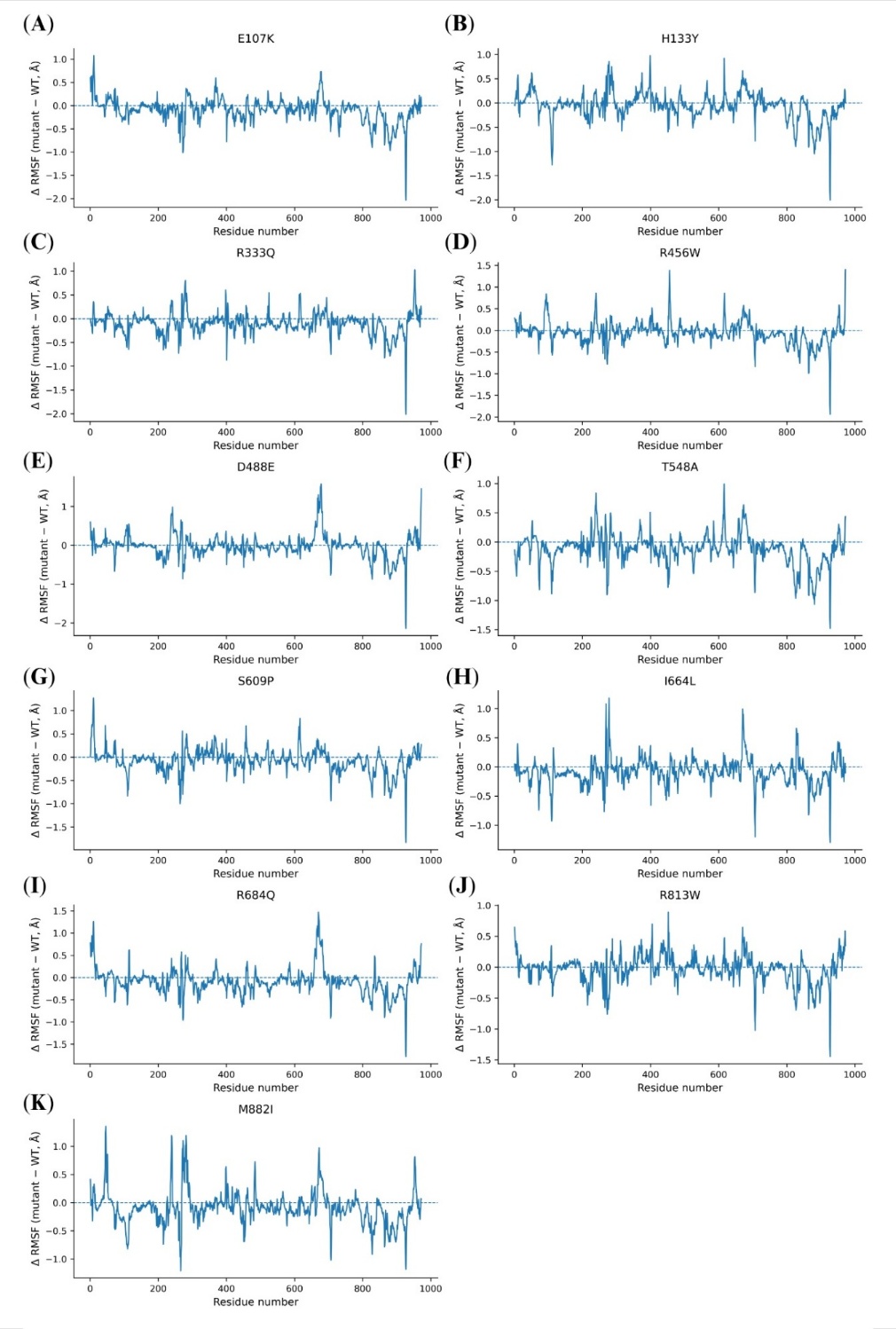

**Figure S51. Same-residue WT-variant Cα RMSF difference profiles for the whole protein.** One panel per variant plots ΔRMSF (variant minus WT) against residue number, with a dashed line at zero. Positive values mean the variant fluctuates more than WT at that residue. Pairing at each residue is preserved, but no residue-wise *P* value is assigned because neighbouring residues are sequence-correlated.

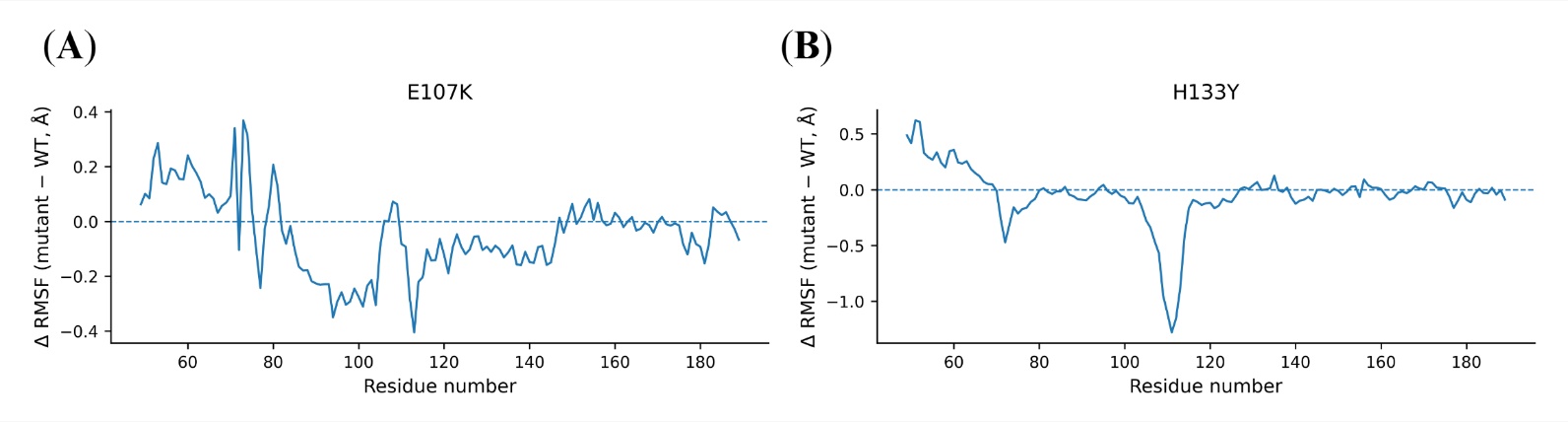

**Figure S52. Same-residue WT-variant Cα RMSF difference profiles for the RUN domain. o**ver residues 49 to 189, for (A) E107K and (B) H133Y.

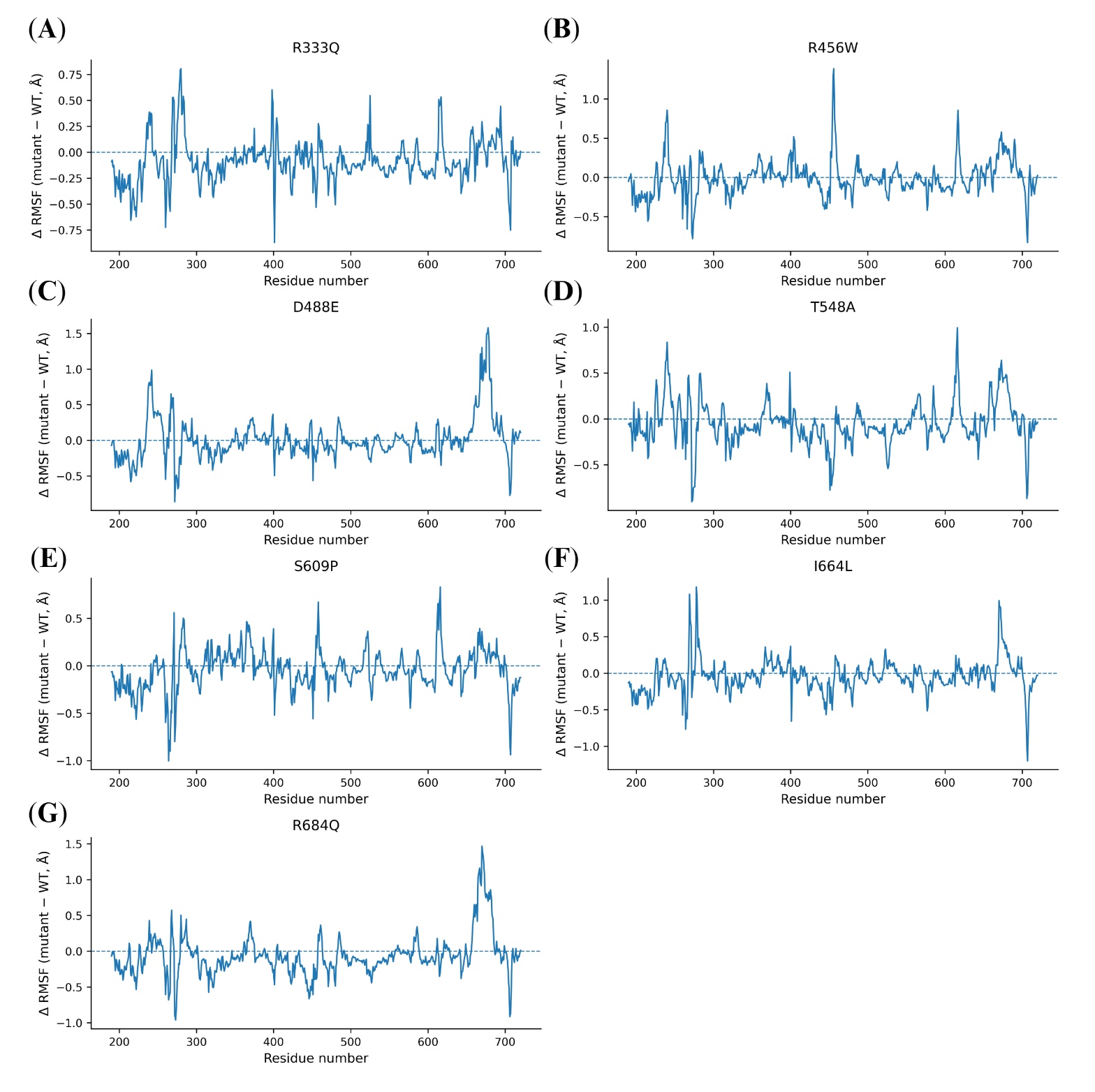

**Figure S53. Same-residue WT-variant Cα RMSF difference profiles for the central region.** Over residues 190 to 720, for the seven variants located in that interval.

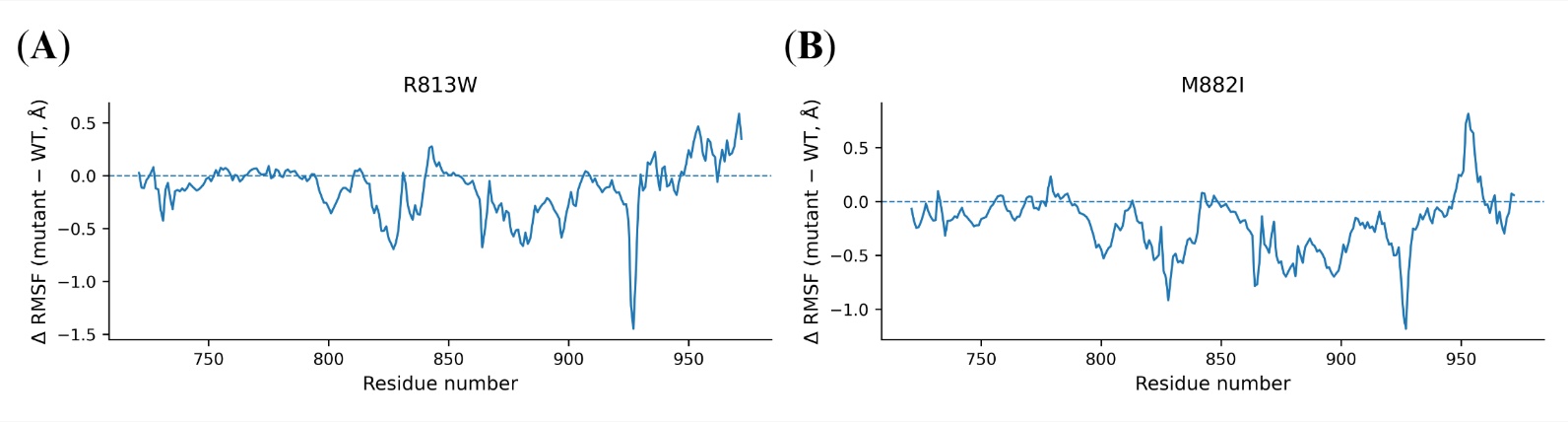

**Figure S54. Same-residue WT-variant Cα RMSF difference profiles for the RH domain.** Over residues 721 to 972, for (A) R813W and (B) M882I.
