## Supplementary material for "Loss of RUBCN causes autophagy overdrive in a neurodevelopmental disorder with age-dependent neurodegeneration": Table 1

| | | | |
|---|---|---|---|
| **Truncating variants** | Ala875Val*fs**146 | C-terminal truncation retains sufficient function for *C. elegans* viability | Abolish RUBCN–RAB7 interaction, causes cytosolic mislocalisation and loss of punctate structures; fail to suppress autophagic flux<br>**Consistent with LOF** |
| | Arg466* | severe LOF incompatible with homozygous viability in *C. elegans* | |
| | Glu519Arg*fs**11 | | |
| | Gln748His*fs**48 | | Predicted loss of the C-terminal/RAB7-binding region<br>**Likely loss of RUBCN–RAB7 interaction & impaired autophagy regulation** |
| **Missense variants** | Thr548Ala | Predicted local structural effect; formation of new residue contacts.<br>No major global conformational change detected. | |
| | Glu107Lys | No detectable structural effect in the modelling. | |
| | Ile664Leu | | |
| | Arg684Gln | Predicted local structural effect; loss of a charge pair/salt-bridge interaction.<br>No major global conformational change detected. | |
| | Arg456Trp | | |
| | Arg333Gln | | |
| | Ser609Pro | Predicted local structural effect with both loss and formation of residue contacts, consistent with the conformational constraints introduced by proline. | |
| | Asp488Glu | Predicted local structural effect; formation of new residue contacts.<br>No major global conformational change detected. | |
| | p.His133Tyr | | |
| | Arg813Trp | Strongest predicted structural effect that could alter RAB7A interaction.<br>Reorganises local packing within RH domain and increases exposure of an apolar surface. | |
